# Small-area estimation of district-level fertility in 36 countries in sub-Saharan Africa 2000-2025

**DOI:** 10.64898/2026.07.17.26358322

**Authors:** Oliver Stevens, Jeffrey W. Imai-Eaton

## Abstract

**Introduction:** District-level estimates of fertility are required by policymakers and programme planners in sub-Saharan Africa (SSA) to guide decision making at the local level. Existing approaches to produce fertility estimates at the district level use age-structures or total fertility trends over time from higher administrative levels which prevents representation of district-level heterogeneity in fertility estimates.

**Methods:** We extracted observed numbers of births and person-years stratified by five-year age group, single calendar year, and district from 194 nationally-representative household surveys. We constructed a spatiotemporal Bayesian hierarchical model that reconciles point and areal data and adjusts for non-sampling biases to estimate age-specific (ASFR) and total fertility rates (TFR) in 36 countries between 2000-2025.

**Results:** Fertility rates were calibrated to three million births from two million women over fifteen million person-years. TFR declined in the majority of countries 2000-2020 (median −15%, interquartile range (IQR) −3 to −24%). Substantial subnational heterogeneity in TFR levels was observed in most countries, exceeding that of differences between national fertility levels. ASFR heterogeneity was observed, though to a lesser degree than variation in total fertility. Teenage fertility was unchanged in several countries, with fertility declines driven by older age groups.

**Conclusion:** District-level fertility dynamics can differ greatly from national TFR trends and age-patterns of fertility. Use of national fertility rates will produce inaccurate estimates of births in the majority of SSA districts, leading to inappropriate resource management for key global public health objectives.

## Introduction

Estimates of age-specific fertility (ASFR) are core to understanding evolving demographic trends and population structures in sub-Saharan Africa (SSA), monitoring the need for and impacts of family planning services in tracking progress towards Sustainable Development Goal 3.7, and guiding myriad public health programmes including maternal and child services, prevention of mother-to-child HIV transmission, prophylactic malaria treatment for pregnant women, and infant vaccination programmes.

Low- and middle-income fertility declines have been observed in Latin America and South East Asia, where countries rapidly transitioned from high to low fertility [1], [2]. While SSA was expected to follow, dynamics have differed and the region was slow to enter fertility transition [3]–[5]. Some countries remain pre-transition, or have experienced fertility declines, followed by plateauing at a level above that of replacement level – “fertility stalls” [6], [7]. Consequently, estimates of SSA fertility are heterogeneous with lower fertility in Southern Africa compared to West Africa, and high fertility clusters in Central Africa and the Sahel [8].

In high-income settings, vital registration systems provide direct measures of fertility, but these are underdeveloped or absent in low- and middle-income countries. Instead, information on fertility is typically derived from nationally representative household surveys and household censuses [9], [10]. National-level estimates of total fertility rate (TFR) and ASFR, such as quinquennial estimates produced by the UN Population Division (UNPD) [1], [2], provide standardised and comparable data for global-level policy, but are often insufficient for public health programming which require higher resolution spatial and temporal data to set targets, allocate resources and commodities, and monitoring programmes. Accordingly, public health policy makers, implementers, and funders demand subnational population and demographic estimates furnished at the level of local decision making and health provisioning—often the second administrative or ‘district’ level.

Systematically produced subnational fertility data are scant. Twenty countries in SSA have developed first administrative level TFR estimates with support from the US Census Bureau [11]. These estimates were constructed allowing different fertility levels across subnational regions, but assumed the same relative age pattern and TFR trend in all regions mirroring the national trend [12]. This does not permit subnational deviation from the national trend or varying age patterns in fertility by area. Ševčíková, Raftery, and Gerland compared approaches for stochastic subnational TFR projections, but did not consider age-specific fertility patterns [13]. The WorldPop project produce estimates for the number of births by 1km x 1km pixel using survey-derived age-specific fertility rates at the first administrative level applied to pixel-level age-specific populations [8], [14]. Deriving district level births using fertility estimates from higher administrative levels may overstate homogeneity between neighbouring areas. At present, there are not estimates of district-level fertility for sub-Saharan Africa which analyse district-level birth data.

Several methodological challenges must be addressed to estimate fertility by granular age, spatial, and temporal strata. Nationally representative household surveys are typically powered to produce estimates at the first administrative level. The number of births observed in a given district and age group is usually very small and therefore direct estimates exhibit large sampling variability and associated uncertainty. Small-area model-based approaches, leveraging spatiotemporal correlation and smoothing, are well established for use in cases of data-sparse areal units, including in estimation of demographic and family planning indicators at the district level [15]–[18]. Secondly, some surveys only publish data located to the first administrative level, but not cluster geocoordinates with which to locate data to districts [19]. Finally, systematic reporting biases have been well documented in Demographic Health Survey birth histories, which affect direct fertility estimates [20]. Interviewers ask extended questions about births occurring in the five years preceding the survey, and abbreviated questions thereafter. Interviewers or respondents may reduce their workload by displacing births beyond the five-year threshold, or omitting recent births altogether, resulting in systematic biases in fertility estimates according to years before the survey [20].

We developed a hierarchical Bayesian model for spatially misaligned household survey data that adjusts for non-sampling bias to estimate TFR and ASFR at the district level for 36 countries in sub-Saharan Africa. We aimed to characterise spatial heterogeneity in TFR trends and age patterns of fertility over time, and to address the variation in subnational fertility relative to prevailing national trends.

## Data and methods

We analysed birth history data from nationally representative household surveys to model district-level ASFRs. Our analysis consisted of three steps. First, household survey clusters were assigned to the administrative level of interest using, preferably, geographic coordinates or survey administrative strata. Second, demographic methods were used to prepare birth and person-year datasets from survey data, stratified by age, space, calendar year, and time preceding the survey (TIPS). Finally, fertility rates were estimated within a spatiotemporal hierarchical Bayesian framework. This permits the exchange of information between adjacent age-space-time units, overcoming the large sampling variability exhibited by direct fertility estimates calculated from fine stratifications with small sample sizes.

### Data extraction and preparation

For each country, the list of health districts, hierarchy of administrative subdivisions, and district geographic boundaries were sourced from subnational geographic data consolidated by UNAIDS for subnational HIV estimates [21]. For each country, we identified the first administrative level at which household sampling were stratified and the second administrative level at which health system planning is administered and estimates are sought (Table S1). For ease of reference, henceforth the first administrative level referred to as “province”, and the chosen administrative level for modelled estimates in each country as “district”. No publicly available survey data was available for Botswana, Equatorial Guinea, or Eritrea.

Data on month of interview, female respondent month of birth, her children’s months of birth, and survey cluster geographic location were extracted from 184 nationally representative household surveys conducted in 36 countries since 1995 that included female birth histories (Table S2). These consisted of 108 Demographic Household Surveys (DHS), 28 Malaria Indicator Surveys (MIS), 4 AIDS Indicator Surveys (AIS), 37 Multiple Indicator Cluster Surveys (MICS) surveys, and 7 Population-based HIV Impact Assessment surveys (PHIA). DHS and some MICS contained full birth histories, enumerating the month of birth for all children ever born to the respondent. Remaining MICS, MIS, and AIS recorded births in the past five years. PHIA recorded births in the one year before the survey.

Most DHS, MIS, AIS, and PHIA surveys provided geographic coordinates for survey cluster centroids, offset by between 2km to 10km to ensure confidentiality. Clusters were assigned to administrative districts via spatial join, ensuring that clusters were also assigned to districts within the same province as the survey strata in which they were sampled. For MICS and earlier DHS surveys for which survey coordinates were not available, clusters were assigned at the lowest administrative level for which information was available, normally the first administrative level.

The survey-weighted number of births and person-years of exposure among female respondents aged 15-49 years were aggregated stratified by geographic area (district or province, as available), five-year age group, single calendar years, and the number of years preceding the survey for 15 years preceding surveys that included full birth histories (DHS and some MICS), five years preceding MIS, AIS, and remaining MICS, and one year for PHIA.

The population size of women of reproductive age by district and age group were extracted by overlaying district geographic boundaries on 100m gridded population rasters produced by the WorldPop project at five-year intervals [22], [23]. These were linearly interpolated to single year estimates. UN Population Division World Population Prospects (WPP) 2022 national populations were interpolated from quinquennial periods to single year [9]. Annual district populations were calibrated to WPP national totals by in each age group. These district populations were used to aggregate district-level ASFRs to province and national estimates.

Survey datasets were extracted with the R package *rdhs* [24] and space-age-time stratified number of births and person years were calculated with the package *demogsurv* [25].

### Statistical analysis

The model consisted of two components: first, a ‘process’ model that represented the true fertility rate *λ_ait_* for women age *a* ∈ (15-19, 20-24, …, 45-49), in district *i* ∈ 1,2, 3, … , *I*, in year *t* ∈ 1995: 2025. Second an ‘observation model’ related the true fertility rates to the ‘observed’ fertility rate reported in a particular survey *s* conducted in year *t*_[*s*]_, accounting for systematic reporting biases related to the number of years *u* preceding the survey (TIPS) to which the observation pertains, *u* = *t*_[*s*]_ − *t*,

#### Process model

The fertility rate *λ_ait_* was modelled as a log-linear function:

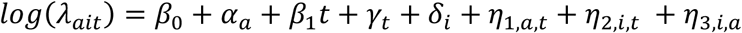

where *β*_0_ is the intercept, *α_a_* captures the typical age pattern of fertility, *β*_1_ is a linear trend, *γ_t_* is represents additional stochastic variation in the national fertility trend, and *δ_i_* represents district-level variation in fertility level. The terms *η*_1,*a*,*t*_, *η*_2,*i*,*t*_, and *η*_3,*i*,*a*_ are two-way interactions [26] permitting variation in fertility trends by age and time, district and time, and district and age, respectively.

The age pattern *α_a_* was modelled via a first order random walk with standard deviation *σ_α_*: *α_a_* ∼ *RW*1(*σ_α_*). Spatial variation was modelled by an intrinsic conditional autoregressive (ICAR) model with standard deviation 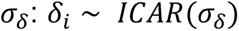. Each of these models are improper and therefore sum-to-zero constraints were specified for the parameters 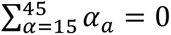 and 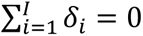

Temporal components were modelled as cubic B-splines with *K* = 9 knots equally spaced every five calendar years from *k* ∈ {1990, 1995, … , 2030}

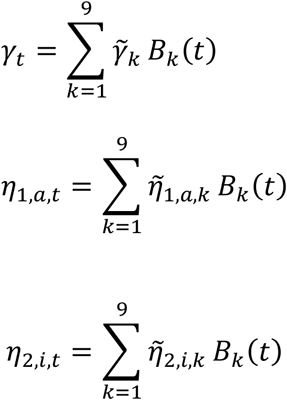

where *B_k_*(*t*) refers to the *k*th B-spline evaluated in year *t* and *γ̃_k_*, *η̃*_1,*a*,*k*_, and *η̃*_2,*i*,*k*_ are the corresponding spline coefficients. The spline coefficients were penalised using time series models to enforce smoothness. Several alternatives were considered for smoothing the coefficients *γ̃_t_* for the national-level time trend (Supplementary Text S1). The ARIMA(1,1,0) model was chosen as the best performing model (an AR1 process penalising the differences between spline coefficients) and used for the remainder of the results.

The interaction terms were modelled as separable via Kronecker products of AR1 models for age and temporal components and ICAR models for spatial components:

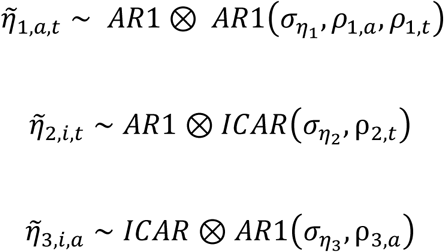

Interactions *η̃*_2,*i*,*t*_ and *η̃*_3,*i*,*a*_ involving improper ICAR priors required sum-to-zero identifiability constraints across districts within each age or year, respectively:

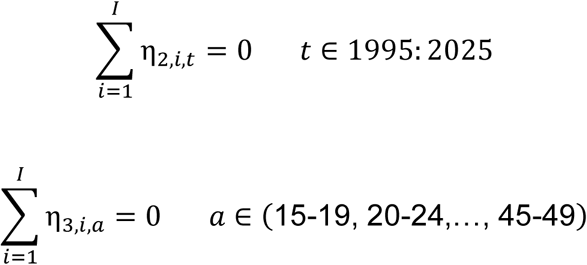

Fixed effects β_c,t_ capturing country-specific fertility shocks in country c in year t were added to the process model capturing the impact of famine in Malawi in years 2001-2002 and the fertility boom in Zimbabwe in 2010-2011. An urban/rural fixed effect was added to the process model in Ethiopia to constrain the estimates in urban areas which were not sampled by surveys.

#### Observation model

The observation model relates the true fertility rate to that which is observed in a household survey, accounting for systematic reporting and recall biases and additional overdispersion arising from unaccounted for non-sampling errors. In DHS, extended questions are asked about births occurring in the five years preceding the survey. Consequently, survey interviewers are incentivised to mis-record children beyond the five year threshold to ask only the abbreviated question set [20]. This birth displacement results in under-enumeration of births before the threshold, and an excess of births following it. Births many years before the survey are also more likely to be misreported due to recall bias, resulting in, for example, heaping of reported births ten years before the survey or years ending in 0 (e.g. 2000).

To adjust for reporting biases, we introduced a series of ‘time preceding survey’ (TIPS) coefficients accounting for the average bias in the reported log fertility rate *u* years before a survey. These coefficients are not distinguishable from true changes in fertility with time series data from a single survey, but systematic reporting biases are identifiable using overlapping data from multiple surveys reporting about fertility in the same year at different recall periods [20] (Supplementary Figure S1).

Several terms captured these biases in the observation model. We introduced fixed effects 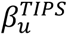 and *u* = 6 years to account for birth displacement and at *u* = 0 and *u* = 10 to adjust for heaping in the year of survey and ten years preceding the survey. For each 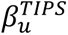, we incorporated an interaction, 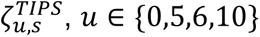 to allow the variation in the displacement effects for each survey, *s*. Secondly, we included time fixed effects 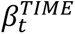 in years *t* ∈ {1999,2000,2001} to account for heaping of births reported in year 2000 and commensurate under-enumeration of births in the years 1999 and 2001. Finally, we included an observation-level random effect *ε_aitsu_* capturing overdispersion arising from other non-modelled error processes.

Taken together, the expected *reported* fertility rate in a survey *s* was function of the true fertility rate *λ_ait_* and the bias terms

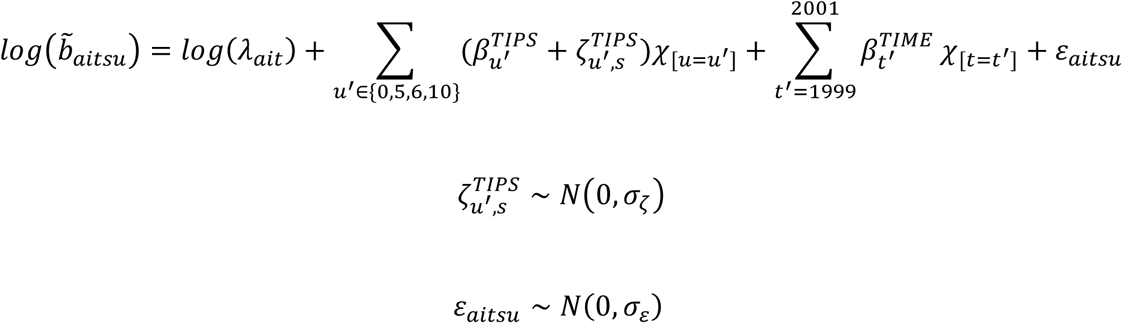

The fixed effect terms had informative prior distributions 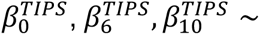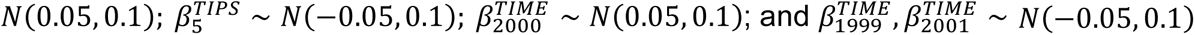

#### Likelihood

For surveys in which clusters were located to a specific district *i*, observations consisted of a (survey weighted) number of births *y_aitsu_* and person-years *E_aitsu_* in survey *s* among women aged *a* in year *t* occurring *u* years before the survey. The likelihood for these observations was

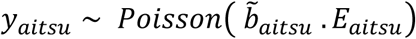

The model was implemented in C++ via the R package Template Model Builder (TMB) [28], and fit separately to survey datasets for each of the 36 countries. One thousand samples were drawn from the joint posterior distribution, from which the posterior mean, median, mode, standard deviation, and quantile-based 95% CI were calculated for each output indicator. Supplementary Text S1 provides further statistical details.

## Results

Data were extracted from 194 surveys, consisting of 17,763,982 person-years of observation and 3,301,400 births. Nearly all countries had multiple surveys (median 5, IQR 4-7), and median last year of survey was 2018.

### District-level variation in TFR

Figure 1 shows district-level TFR estimates in 2018, the median year of last survey across all countries. Total fertility was lowest in Southern Africa and highest in the Sahel region, Angola, and the Democratic Republic of the Congo (DRC). While total fertility differed between countries at the national level, in most countries, heterogeneity was greater at the district level within country than variation between countries across SSA regions. The largest variation was in in Ethiopia where TFR ranged across districts from 0.91 to 7.42 (Figure 1B). In most countries (29/36), the national TFR was below the median TFR across districts. This was because more populous districts tended to have lower TFRs (Figure 1B; point sizes), for example urban areas and capital cities.

**Figure 1:**
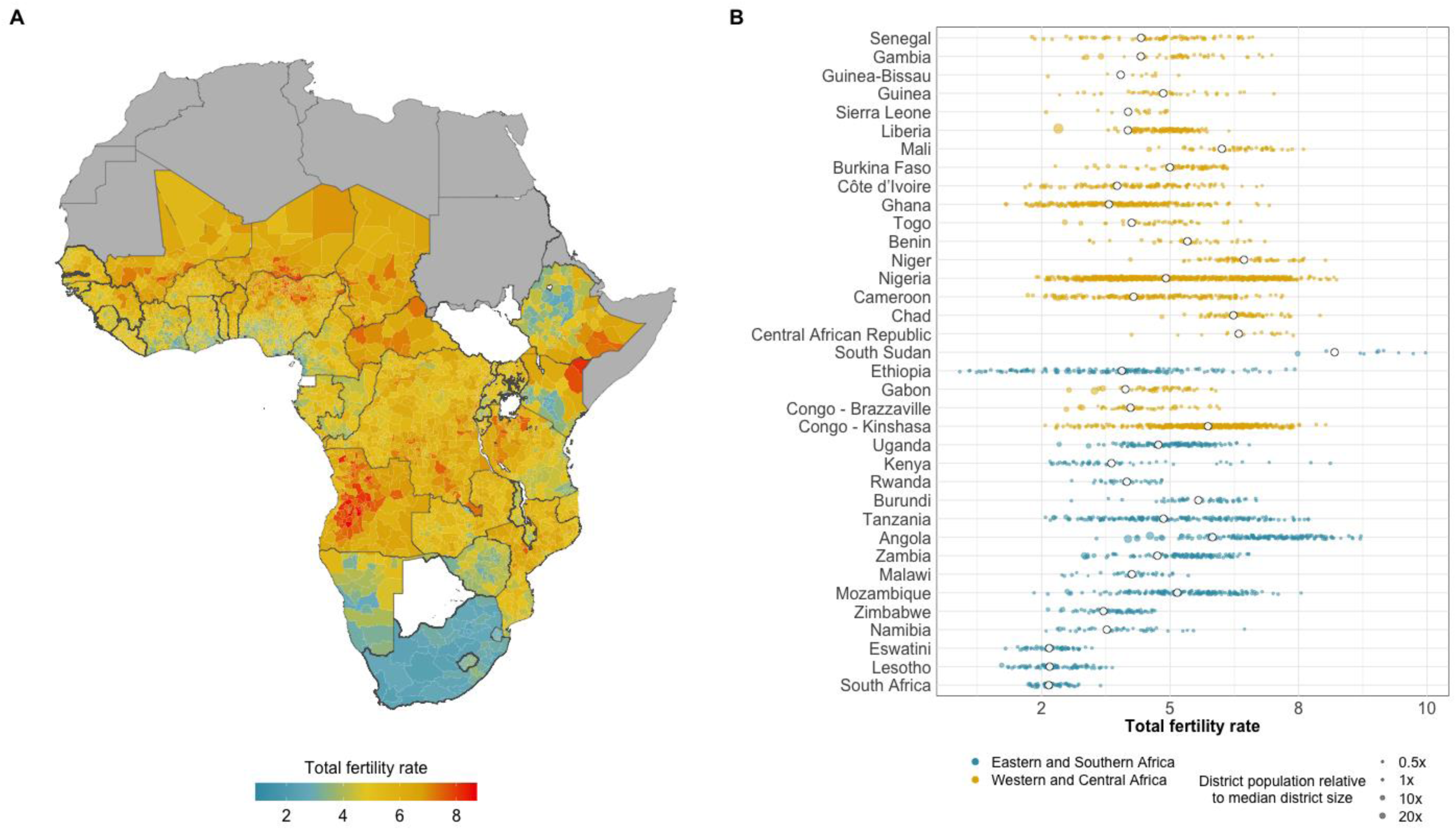
District-level estimates of total fertility rate in 2018. In (B) coloured points represent district estimates and open circles represent national estimate. Points are sized by district population relative to median district population size in each country. Countries are ordered North-West to South-East.

### Trends in TFR

Total fertility rate declined at the national level in the majority of countries during the period 2000-2020 (median change −22%, interquartile range [IQR] −32 to −16% across countries). Declines were similar in Eastern and Southern Africa (−20% IQR - 35 to −16%) and in Western and Central Africa (−22% IQR −29 to −16%). National fertility declines stalled in Mozambique (2011-2017), Namibia (2000-2017), Rwanda (2011-2018), and Zimbabwe (2003-2019) (Supplementary File 2). Within countries, district-level TFR trends largely followed national TFR trajectories (Figure 2A-C), with larger heterogeneity observed in some countries (Figure 2D-F). In 2018, 0.9% of districts (36/4,013) had total fertility beneath replacement level of 2.1 births per woman, of which 24 were in Ethiopia. Supplementary File 2 contains national and province-level TFR trends compared with estimates from household surveys.

**Figure 2:**
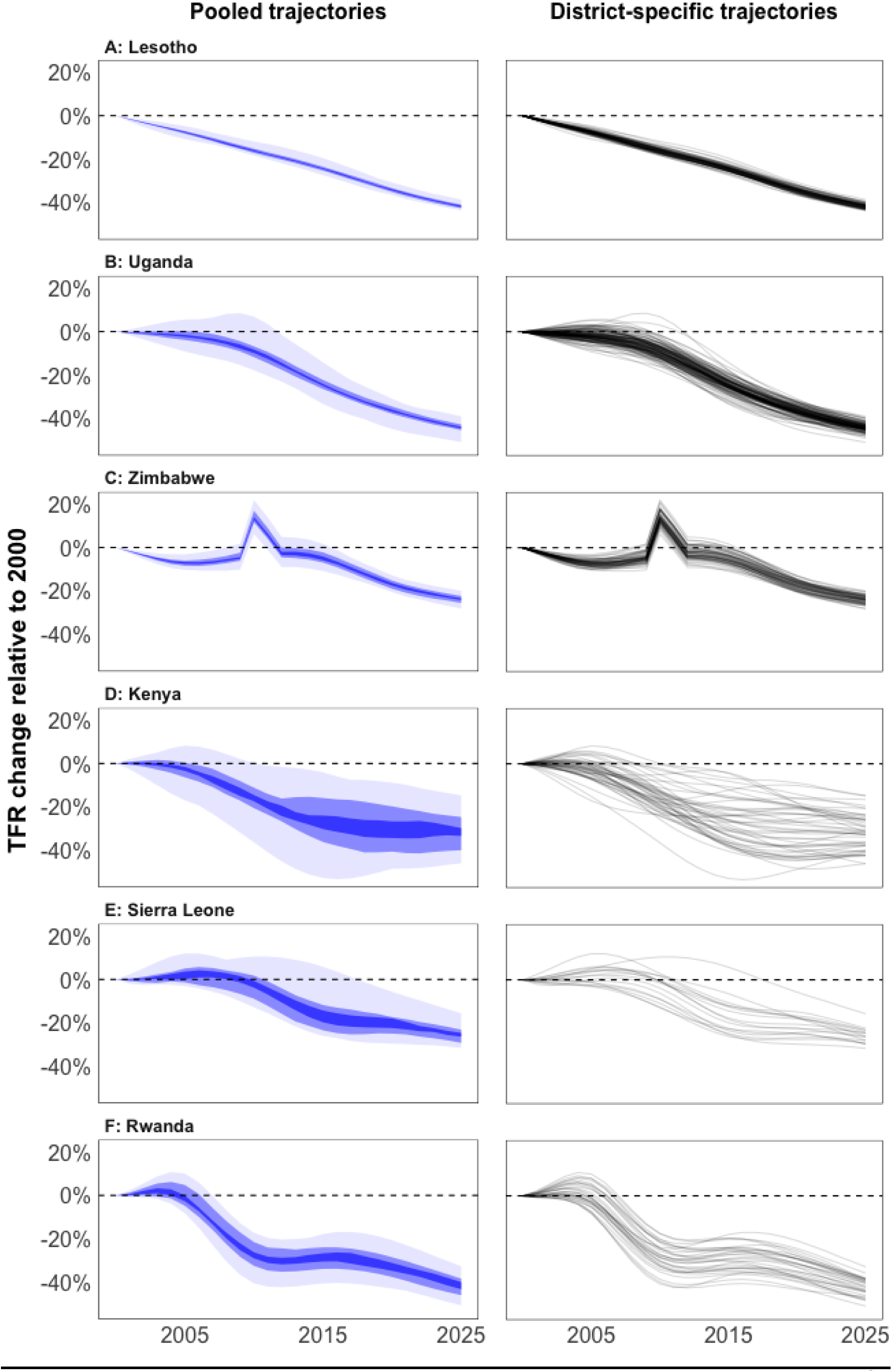
(A) District-level estimates of total fertility rate (TFR) from 1995-2000 in Tanzania, Senegal, and Ghana. National-level TFR shown in black and selected districts shown in colour. (B) Regional (Level 1) estimates of TFR from 1995-2000 in Ethiopia. National-level TFR shown in black and states in colour.

### Spatial variation and trends in age-specific fertility rates (ASFR)

Age patterns of fertility varied spatially in most countries, though to a lesser degree than spatial variation in total fertility. The ratio between ASFRs in the peak fertility age groups (ASFR among age 25-29 years relative age 20-24 years) illustrated the variation in district-level age patterns of fertility (Figure 3A). Fertility peaked among age 20-24 in the majority of Eastern and Southern Africa, while peak fertility was among the 25-29 age group in Western and Central Africa. District-level fertility ratios differed by up to 20% in several countries, with the most district-level variation in Nigeria where fertility in 25-29 year olds varied from −31 to +100% of that in 20-24 year olds. In 2016, the lowest mean age of child bearing was in Lesotho (27.7 years) and the highest was in South Sudan (30.8 years). Throughout SSA, fertility was older in provinces containing large urban areas than surrounding provinces. Senegal, Nigeria, and The Gambia had an older mean age of fertility in western than in eastern provinces, and central Burundi had an older age pattern than border provinces (Figure 3B).

**Figure 3:**
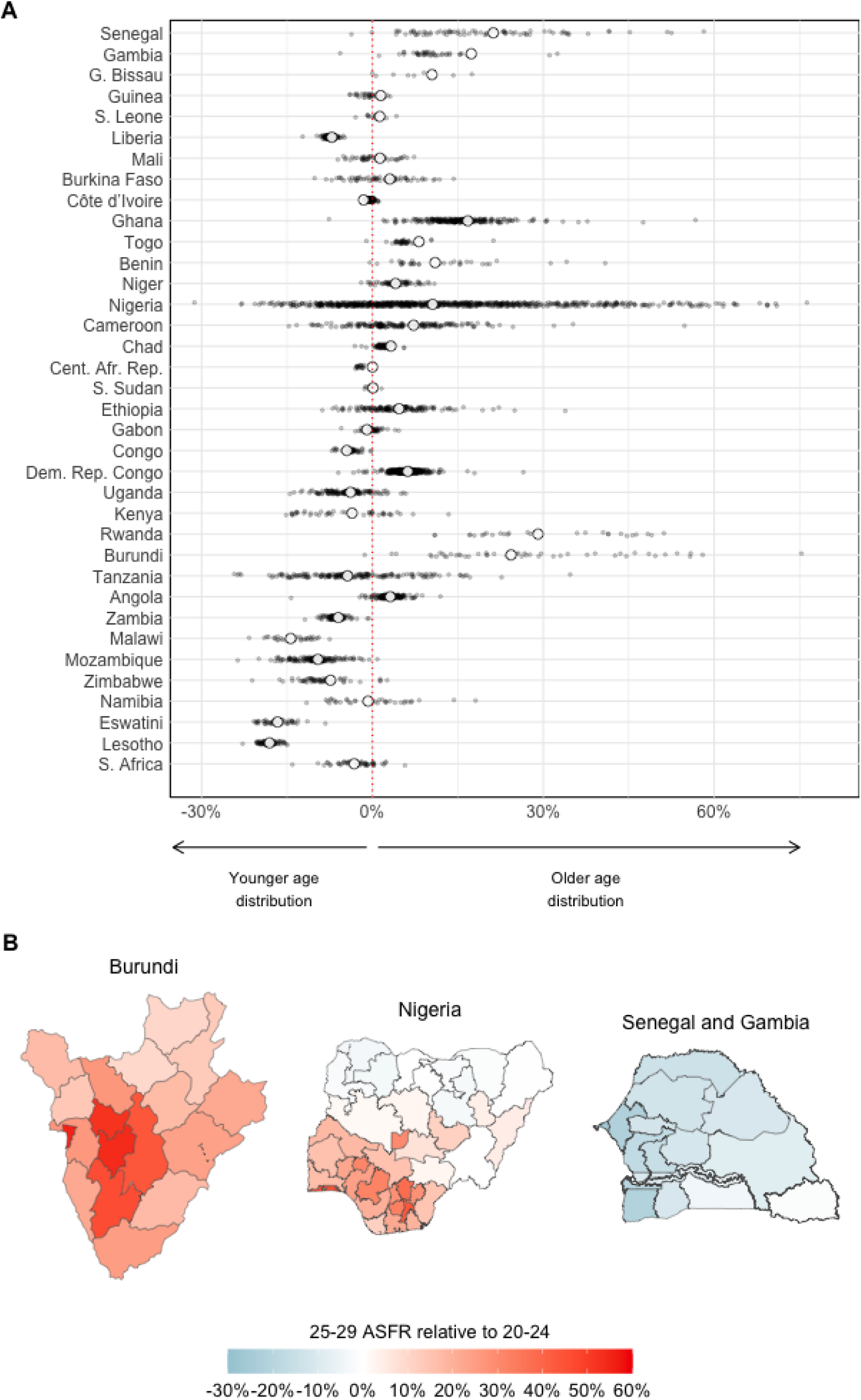
(A) ASFR in ages 25-29 expressed relative to ASFR in ages 20-24 at the district-level (coloured points) and national level (white circle). Countries are ordered North-West to South-East. (B) Spatial heterogeneity in ASFR ratio between 25-29 and 20-24 in Burundi, Nigeria, and Senegal and Gambia

Fertility declines were not equal in all age groups (Figure 4A). Across all countries, median fertility declined most among women aged 40-49 years in both ESA and WCA (−42% and −33% respectively), while fertility in the highest fertility age groups (20-24 and 25-29) declined 18% in both ESA and WCA. Subnational variation in ASFR trends was common throughout sub-Saharan Africa. This encompassed different fertility trends across age groups (Figure 4B) and varying trends within a single age group in different spatial units (Figure 4C).

**Figure 4:**
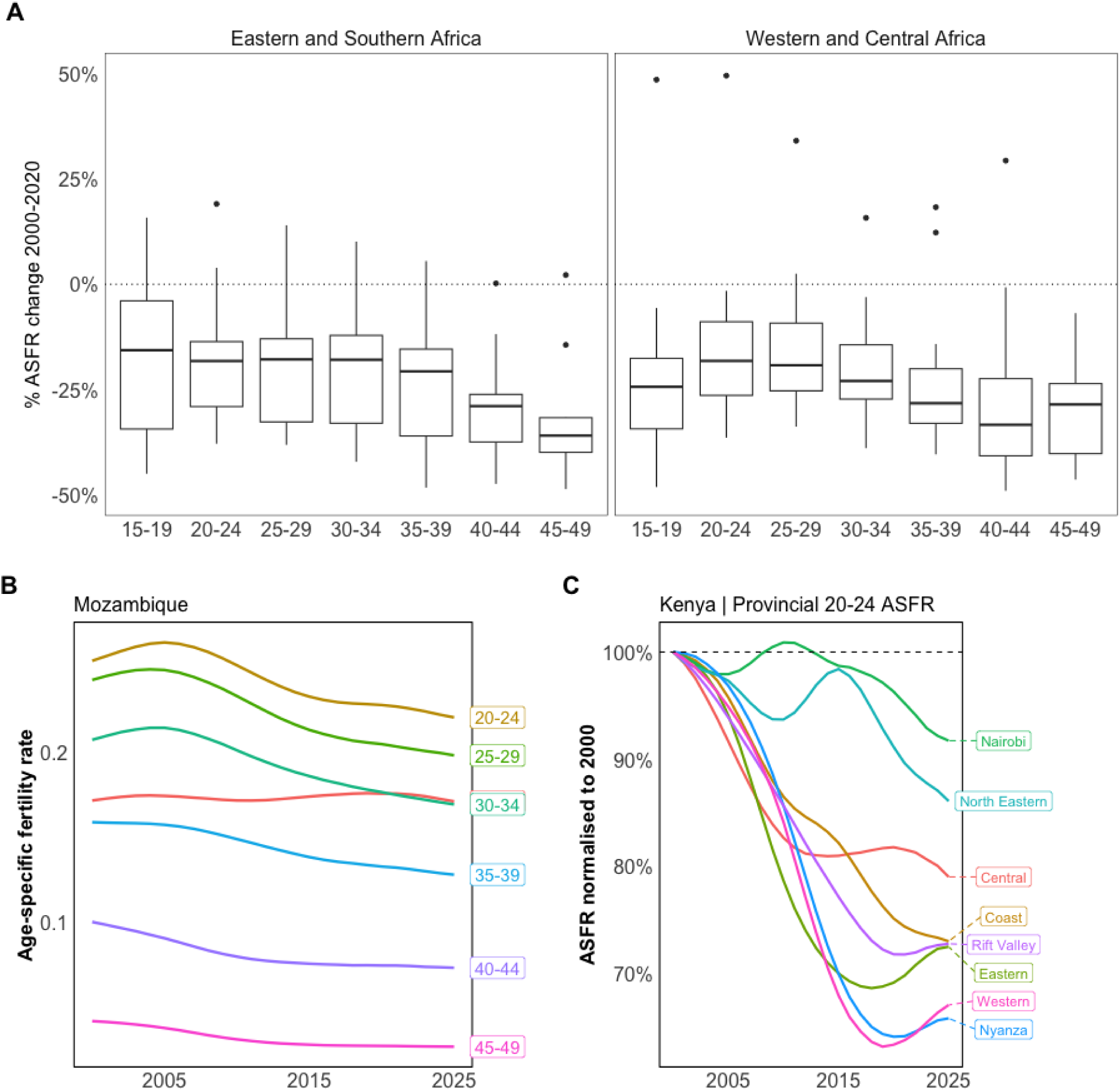
(A) Percentage change in national ASFR from 2000 to 2020 in Eastern and Southern Africa and Western and Central Africa. Box plots indicate range across countries in each region. (B) National level ASFR trends in Mozambique from 2000 to 2025. Shaded areas indicate 95% credible intervals. The figure illustrates relatively flat adolescent (15-19 years) fertility compared to larger fertility reductions among older age groups. (C) Relative change in ASFR among age 20-24 years from 2000 to 2025 in Kenya former provinces. Values are normalised to ASFR in year 2000 for each province.

**Figure 5:**
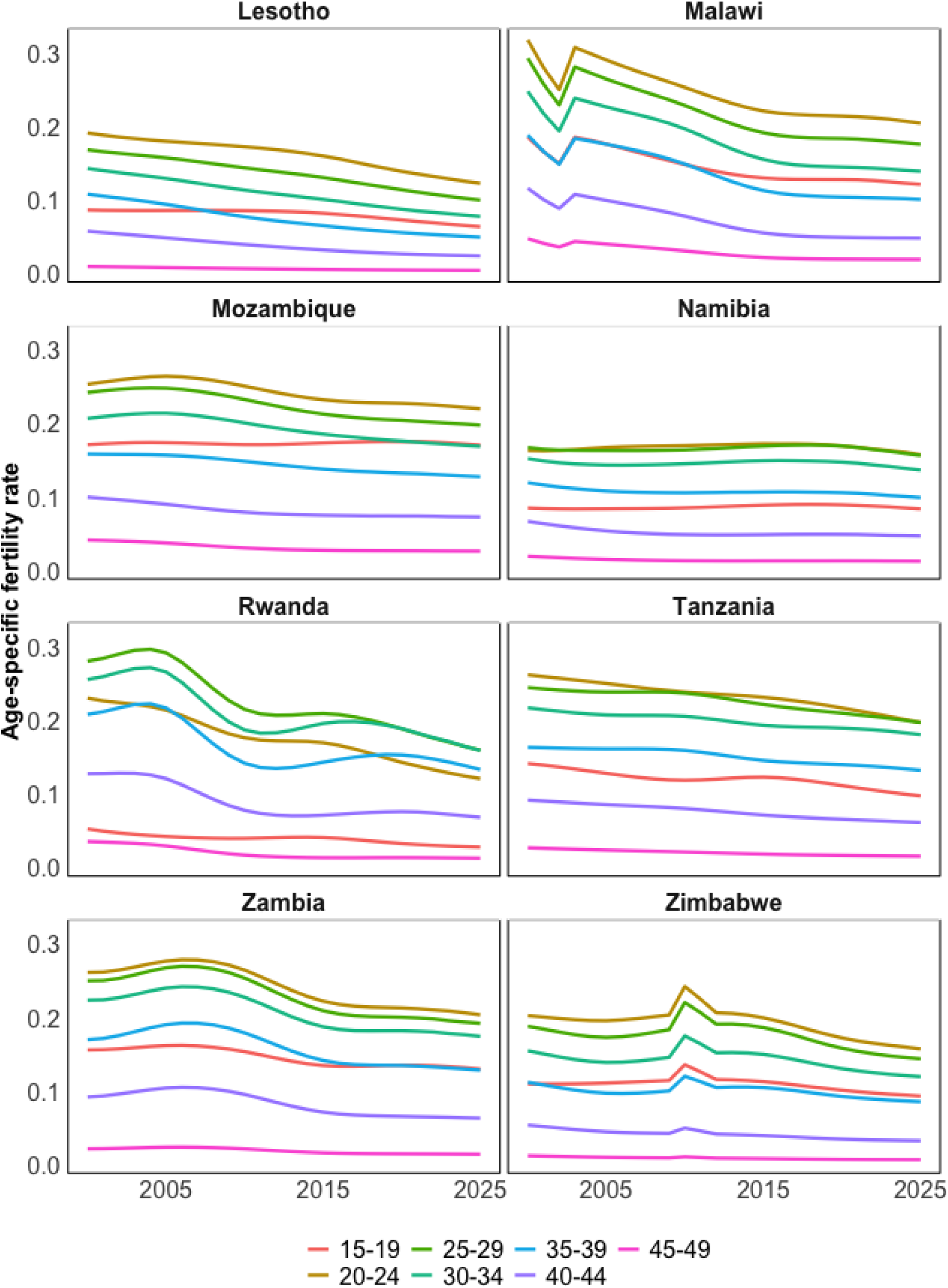
Unchanged or minimally decreasing fertility amongst 15-19 year olds in Eastern and Southern Africa.

Fertility trends in 15-19 year olds from 2000-2020 remained unchanged or decreased minimally in several countries. This was especially the case in Eastern and Southern Africa, where 15-19 year olds had the smallest relative fertility decline across all age groups in Rwanda, Tanzania, Mozambique, Zambia, Malawi, Zimbabwe, and Lesotho (Figures 4A & 5).

### Observation model estimates and reporting biases

Birth displacement and data heaping were substantial in most countries (Figure S1). Across all countries, year-of-survey fertility (TIPS = 0) was 3% higher (median across countries; IQR 0-8%); displaced fertility in DHS surveys (TIPS = 6) was 5% higher (IQR 0-9%); and ten-year recall heaping (TIPS = 10) was 5% higher (IQR −5 to +10%) than estimated true fertility.

## Discussion

National-level fertility in sub-Saharan Africa has received considerable attention as the remaining high-fertility region of the world with appreciable uncertainty and debate about the prospects for further fertility transition, with implications for global population, economic development, health, and wellbeing. We have added to this discourse by characterising fertility levels, age patterns, and changes at subnational level across sub-Saharan Africa by developing a spatiotemporal model that reconciles sparse areal and point survey data. Our resulting fertility estimates stratified by year, district, and age group also have probabilistic uncertainty intervals, providing estimates and ranges appropriate for an array of public health and demographic programmatic planning and decision making. Estimates of total fertility and age-specific fertility at the national level were consistent with UN World Population Prospects estimates in the majority of countries.

We found that subnational heterogeneity in total fertility rate is commonplace, including in the most populous countries in sub-Saharan Africa: Nigeria, Ethiopia, the DRC, and Tanzania. In smaller countries where absolute total fertility differences between districts are lower, the national TFR was often much lower than the median of district-level estimates, drawn by urban centres with large populations and low fertility. Further, applying national-level age patterns and trends in fertility rate to all areas constrains and misrepresents local dynamics. In these countries, district-level birth estimates calculated using the national ASFRs will lead to substantial misestimation and poor resource management.

Total fertility was lower in countries in Eastern and Southern Africa that have higher levels of modern contraceptive use (mCP) and TFR higher in Western and Central Africa where mCP use is lower [30]. Small-area analyses have highlighted district-level disparities mCP use within Ethiopia, Nigeria, Uganda, and national-level disparities between DRC and neighbouring countries in ESA where higher mCP coverage is mirrored by lower TFR estimates in our analysis [17], [18], [31]. We estimated high teenage fertility relative to total fertility levels in several countries, echoing a recent subnational analysis of fertility rates in sub-Saharan Africa [8]. Analysis of household survey data finds that teenage women in SSA have low mCP use and high unmet need [32]. Given the negative health and social outcomes associated with teenage pregnancy for both mother and child, these trends in teenage fertility and contraceptive use remain concerning. High-resolution estimates of mCP, unmet contraceptive need, and ASFR can be used to focus family planning programmes and facilitate progress towards Family Planning 2030 and SDG 3.7. Future modelling approaches should consider jointly modelling these indicators.

Fertility stalls, when countries that have initiated the fertility transition plateau at a TFR above replacement level, have been widely described in sub-Saharan Africa [6], [7], [33]. Survey- and census-based evidence for fertility stalls have been synthesised by Schoumaker [6], who identified strong evidence for stalls in Congo, Kenya, Namibia, Zambia, and Zimbabwe. Recent survey data in Zimbabwe (2016 PHIA, 2019 MICS) and Namibia (2017 PHIA) support continued stalled fertility in both countries. This analysis does not support a fertility stall in Kenya, where we estimated consistently declining fertility since 2000. Three recent surveys in Rwanda (2016 PHIA, 2017 MIS, 2019 DHS) indicate that following a large fertility decline from 2005-2010, and a stall from 2010-2017, the fertility transition may have re-initiated. In Mozambique, constant national TFR as also reported by Schoumaker [6], masked diverging subnational trends. Fertility declined sharply in southern provinces, but was steady or increased in populous northern provinces. We suggest that subnational estimates should be included in the characterisation of fertility stalls to discriminate between ‘true’ stalls—where all subnational units experience unchanging fertility—versus cases where apparently stalled fertility masks more nuanced subnational dynamics.

Additionally to modelling subnational ASFR dynamics, systematically accounting for birth displacement and heaping across all survey types was a major methodological innovation in our analysis. The household survey data we analysed are ubiquitously used in other demographic assessments [8]–[10], [34], but instead of explicitly accounting for known systematic deficiencies in the data, common approaches are to aggregate data into, for example, three-year intervals that conceal the heaping effects apparent in annualised birth history estimates. An unexpected finding during our model development was 3% higher reported fertility in the year-of-survey. This requires further investigation as it is contrary to existing literature which finds evidence for omission of recent births in surveys [20] and under-enumeration of young children in censuses [35]. Direct survey fertility estimates use the three years preceding survey [34] which may lead to overestimation of fertility or inappropriately diagnosing a fertility stall due to year-of-survey effects [36].

Presently, the estimates in several WCA countries are inconsistent with other estimates of national TFR trends and exhibit implausible variability over time. Our model requires further development for these countries. We believe that the challenges estimating subnational ASFRs for WCA countries may be related to the nature, amount, and characteristics of data available in this region. First, age heaping is more severe in surveys and censuses in WCA region, which may implicate other data quality and consistency challenges [20]. Second, ESA countries have high density of DHS surveys with full birth histories, while MICS surveys are most common in WCA countries. This poses several challenges: (1) MICS surveys do not provide district-level resolution data; (2) birth histories in most WCA MICS are only for the five years preceding survey, rather than the fifteen years in ESA MICS; (3) preliminary evidence suggests that there may be data quality issues with five-year birth histories in some MICS surveys; (4) MICS surveys are smaller than DHS surveys, resulting in fewer person-years in each stratum; and (5) identifying the TIPS fixed and random effects is challenging with MICS surveys. In several countries, the modelled estimates insufficiently capture spatially heterogeneous fertility changes apparent in crude fertility estimates, particularly failing to capture downward trends in recent fertility in some major urban areas (e.g. Nairobi, Maputo, several provinces in Ethiopia).

This analysis has several limitations and future avenues for development. First, the model uses district-level population denominators to aggregate district-level births and calculate fertility rates at higher administrative levels. Uncertainty in these population estimates is not reflected in the final fertility estimate uncertainty, which may be sizable at fine spatial resolution. Second, several sources of birth and fertility data exist that are not included in model calibration, including summary birth histories from other survey-based sources and census data. These may improve fertility estimates, particularly in countries with infrequent surveys. Third, estimates are not adjusted for survey non-response, nor for women who died prior to the survey who may have had higher parity than surviving women.

## Data Availability

All data produced in the present study are available upon reasonable request to the authors

## References

[1] L. Alkema, A. E. Raftery, P. Gerland, S. J. Clark, and F. Pelletier, “Estimating trends in the total fertility rate with uncertainty using imperfect data: Examples from West Africa,” Demogr. Res., vol. 26, pp. 331–362, 2012, doi: 10.4054/DemRes.2012.26.15.

[2] L. Alkema et al., “Probabilistic Projections of the Total Fertility Rate for All Countries,” Demography, vol. 48, no. 3, pp. 815–839, Aug. 2011, doi: 10.1007/s13524-011-0040-5.

[3] P. Gerland, A. Biddlecom, and V. Kantorová, “Patterns of Fertility Decline and the Impact of Alternative Scenarios of Future Fertility Change in sub-Saharan Africa,” Popul. Dev. Rev., vol. 43, pp. 21–38, May 2017, doi: 10.1111/padr.12011.

[4] A. Dasgupta, M. Wheldon, V. Kantorová, and P. Ueffing, “Contraceptive use and fertility transitions: The distinctive experience of sub-Saharan Africa,” Demogr. Res., vol. 46, pp. 97–130, 2022, doi: 10.4054/DEMRES.2022.46.4.

[5] J. Bongaarts, “Trends in fertility and fertility preferences in sub-Saharan Africa: the roles of education and family planning programs,” Genus, vol. 76, no. 1, p. 32, Dec. 2020, doi: 10.1186/s41118-020-00098-z.

[6] B. Schoumaker, “Stalls in Fertility Transitions in sub-Saharan Africa: Revisiting the Evidence,” Stud. Fam. Plann., vol. 50, no. 3, pp. 257–278, Sep. 2019, doi: 10.1111/sifp.12098.

[7] D. A. Sánchez-Páez and B. Schoumaker, “Fertility Transition in Africa: What do we know and what have we learned about Fertility Stalls? Introduction: The (particular) demographic transition.”

[8] C. Pezzulo et al., “Geographical distribution of fertility rates in 70 low-income, lower-middle-income, and upper-middle-income countries, 2010–16: a subnational analysis of cross-sectional surveys,” Lancet Glob. Heal., vol. 9, no. 6, pp. e802–e812, Jun. 2021, doi: 10.1016/S2214-109X(21)00082-6.

[9] UN Population Division, “World Population Prospects -Population Division -United Nations.” Accessed: Apr. 03, 2020. [Online]. Available: https://population.un.org/wpp/.

[10] C. J. L. Murray, C. S. K. H. Callender, and X. R. Kulikoff, “Population and fertility by age and sex for 195 countries and territories, 1950–2017: a systematic analysis for the Global Burden of Disease Study 2017,” Lancet, vol. 392, no. 10159, pp. 1995–2051, Nov. 2018, doi: 10.1016/S0140-6736(18)32278-5.

[11] US Census Bureau, “Subnational Projections Toolkit,” 2016. [Online]. Available: http://www.census.gov/population/international/software/sptoolkit/.

[12] UNAIDS Reference Group on Estimates Modelling and Projections, “Methods and assumption for subnational demographic inputs to Spectrum in sub-Saharan Africa,” 2019.

[13] H. Ševciková, A. E. Raftery, and P. Gerland, “Probabilistic projection of subnational total fertility rates,” Demogr. Res., vol. 38, no. 1, pp. 1843–1884, 2018, doi: 10.4054/DemRes.2018.38.60.

[14] A. J. Tatem, J. Campbell, M. Guerra-Arias, L. de Bernis, A. Moran, and Z. Matthews, “Mapping for maternal and newborn health: The distributions of women of childbearing age, pregnancies and births,” Int. J. Health Geogr., vol. 13, no. 1, p. 2, Jan. 2014, doi: 10.1186/1476-072X-13-2.

[15] Z. Li et al., “Changes in the spatial distribution of the under-five mortality rate: Small-area analysis of 122 DHS surveys in 262 subregions of 35 countries in Africa.,” PLoS One, vol. 14, no. 1, p. e0210645, 2019, doi: 10.1371/journal.pone.0210645.

[16] J. Wakefield, G. A. Fuglstad, A. Riebler, J. Godwin, K. Wilson, and S. J. Clark, “Estimating under-five mortality in space and time in a developing world context,” Stat. Methods Med. Res., vol. 28, no. 9, pp. 2614–2634, Sep. 2019, doi: 10.1177/0962280218767988.

[17] Q. Li, T. A. Louis, L. Liu, C. Wang, and A. O. Tsui, “Subnational estimation of modern contraceptive prevalence in five sub-Saharan African countries: a Bayesian hierarchical approach,” BMC Public Health, vol. 19, no. 1, p. 216, Dec. 2019, doi: 10.1186/s12889-019-6545-3.

[18] L. D. Mercer, F. Lu, and J. L. Proctor, “Sub-national levels and trends in contraceptive prevalence, unmet need, and demand for family planning in Nigeria with survey uncertainty,” BMC Public Health, vol. 19, no. 1, pp. 1–9, Dec. 2019, doi: 10.1186/s12889-019-8043-z.

[19] N. Marquez and J. Wakefield, “Harmonizing Child Mortality Data at Disparate Geographic Levels,” 2016.

[20] B. Schoumaker, “Quality and consistency of DHS fertility estimates,” 2014.

[21] J. W. Eaton et al., “Naomi: a new modelling tool for estimating HIV epidemic indicators at the district level in sub-Saharan Africa,” J. Int. AIDS Soc., 2021, doi: 10.1002/jia2.25788.

[22] C. Linard, M. Gilbert, R. W. Snow, A. M. Noor, and A. J. Tatem, “Population Distribution, Settlement Patterns and Accessibility across Africa in 2010,” PLoS One, vol. 7, no. 2, p. e31743, Feb. 2012, doi: 10.1371/journal.pone.0031743.

[23] “WorldPop.” https://www.worldpop.org/ (accessed Mar. 28, 2022).

[24] O. J. Watson, R. FitzJohn, and J. W. Eaton, “rdhs: an R package to interact with The Demographic and Health Surveys (DHS) Program datasets,” Wellcome Open Res., vol. 4, p. 103, Jun. 2019, doi: 10.12688/wellcomeopenres.15311.1.

[25] J. W. Eaton, “mrc-ide/demogsurv: Analysis of demographic indicators from Demographic and Health Surveys (DHS) and other household surveys.” Accessed: Mar. 31, 2020. [Online]. Available: https://github.com/mrc-ide/demogsurv.

[26] L. Knorr-Held, “Bayesian modelling of inseparable space-time variation in disease risk,” Stat. Med., vol. 19, no. 17-18, pp. 2555–2567, 2000, doi: doi:10.1002/1097-0258(20000915/30)19:17/18<2555::AID-SIM587>3.0.CO;2-#.

[27] “Autoregressive model of order 1 (AR1) Parametrization.”

[28] K. Kristensen, A. Nielsen, C. W. Berg, H. Skaug, and B. M. Bell, “TMB: Automatic differentiation and laplace approximation,” J. Stat. Softw., vol. 70, no. 1, pp. 1–21, Apr. 2016, doi: 10.18637/jss.v070.i05.

[29] L. Alkema and J. R. New, “Global estimation of child mortality using a Bayesian B-spline bias-reduction model,” Ann. Appl. Stat., vol. 8, no. 4, pp. 2122–2149, 2014, doi: 10.1214/14-AOAS768.

[30] N. Cahill et al., “Modern contraceptive use, unmet need, and demand satisfied among women of reproductive age who are married or in a union in the focus countries of the Family Planning 2020 initiative: a systematic analysis using the Family Planning Estimation Tool,” Lancet, vol. 391, no. 10123, pp. 870– 882, Mar. 2018, doi: 10.1016/S0140-6736(17)33104-5.

[31] J. L. Proctor and L. D. Mercer, “Estimating the levels and trends of family planning indicators in 436 sub-national areas across 26 countries in sub-Saharan Africa,” medRxiv, p. 2021.03.03.21252829, Mar. 2021, doi: 10.1101/2021.03.03.21252829.

[32] V. Kantorová, M. C. Wheldon, A. N. Z. Dasgupta, P. Ueffing, and H. C. Castanheira, “Contraceptive use and needs among adolescent women aged 15–19: Regional and global estimates and projections from 1990 to 2030 from a Bayesian hierarchical modelling study,” PLoS One, vol. 16, no. 3, p. e0247479, Mar. 2021, doi: 10.1371/journal.pone.0247479.

[33] E. Kebede, A. Goujon, and W. Lutz, “Stalls in Africa’s fertility decline partly result from disruptions in female education,” Proc. Natl. Acad. Sci. U. S. A., vol. 116, no. 8, pp. 2891–2896, Feb. 2019, doi: 10.1073/pnas.1717288116.

[34] Demographic Health Surveys, “Demographic and Health Surveys Methodology,” 2006.

[35] F. Pelletier, S. Hertog, B. Hovy, K. Schmid, F. Swiaczny, and J. Wilmoth Guangyu Zhang, “Census counts, undercounts and population estimates: The importance of data quality evaluation,” 2020. Accessed: Mar. 29, 2022. [Online]. Available: www.un.org/ga/search/view_doc.asp?symbol=A/RES/70/1&Lang=E.

[36] B. Schoumaker, “Stalls in Fertility Transitions in Sub-Saharan Africa: Real or Spurious?.” Accessed: Mar. 29, 2022. [Online]. Available: https://www.researchgate.net/publication/265084607.

