## Supplementary File 1 for "Small-area estimation of district-level fertility in 36 countries in sub-Saharan Africa 2000-2025"

### Supplementary Text S1: Statistical model details

#### Aggregation model

For surveys in which clusters were only located to a province $J$ (the survey strata), observations consisted of the number of births $y_{aJtsu}$ and person years $E_{aJtsu}$ observed in each province in the survey. The likelihood was constructed by a weighted aggregation of the district-level fertility rates for all districts $\left\{ i \right\}\in J$ to a predicted ASFR for the province and adding the observation model components:

$$\tilde{b}_{aJtsu}=\log\left( \frac{\sum_{i\in J} \lambda_{ait}P_{ait}}{\sum_{i\in J} P_{ait}} \right)+\sum_{u^{'}\in\{0,5,6,10\}} (\beta_{u^{'}}^{TIPS}+\zeta_{u^{'}, s}^{TIPS})\chi_{[u=u^{'}]}+\omega_{u}^{TIPS}+\sum_{t^{'}=1999}^{2001} \beta_{t^{'}}^{TIME}\chi_{[t=t^{'}]}+\varepsilon_{aitsu}$$

where $P_{ait}$ is the population size of women aged $a$ in district $i$in year $t$. Then the likelihood for the province-level survey data is analogously

$$y_{aJtsu}\sim Poisson\left( \tilde{b}_{aJtsu} . E_{aJtsu} \right).$$

#### Priors and hyperpriors

Diffuse priors were specified on the model intercept and linear time trend: $\beta_{0}, \beta_{1}\sim N(0, 5)$. All random effects have variances parameters which are estimated from the data with gamma distributed hyperpriors: $\Gamma\left( 1, 2\times{10}^{5} \right).$Lag logit-transformed autocorrelation parameters were given normal hyperpriors: $N(0, 2.5)$ [27].

#### Computation and calculation

The model was implemented in C++ via the R package Template Model Builder (TMB), which provides analytical gradients for the posterior density via automatic differentiation and Laplace approximation of the marginal posterior [28]. The posterior distribution was approximated with an empirical Bayes strategy. The distribution of latent model parameters (fixed and random effect terms in the process and observation model) were approximated via Laplace approximation conditional on maximum a posteriori estimates for model hyper-parameters. One thousand samples were drawn from the joint posterior distribution, from which the posterior mean, median, mode, standard deviation, and quantile-based 95% CI were calculated for each output indicator.

### Supplementary Text S1: Model selection

Several alternatives were considered for smoothing the coefficients γ ̃_t for the national-level time trend:

1. First order random walk penalised B-splines (RW1)
2. RW1 penalised B-splines with linear time trend (RW1 + trend)
3. Second order random walk penalised B-splines (RW2)
4. Second order random walk penalised B-splines with linear time trend (RW2 + trend)
5. First order autoregressive penalised B-splines (AR1)
6. AR1 penalised B-splines with linear time trend (AR1 + trend)
7. AR1 penalising the differences between spline coefficients (ARIMA(1,1,0))
8. ARIMA(1,1,0) with linear time trend (ARIMA(1,1,0) + trend)

In the variants of models without a ‘trend’ term the value $\beta_{1}$ (linear time trend) was fixed to zero and all national-level temporal variation was captured via the temporal random effects terms $\gamma_{t}$. The models with RW1 or RW2 term had a single hyper-parameter $\sigma_{\gamma}$. Models with AR1 or ARIMA(1,1,0) were defined by an additional correlation parameter $\rho_{\gamma}.$

The seven model specifications for the temporal component $\gamma_{t}$ were assessed by cross-validation. The cross-validation scheme was designed with particular interest in performance characteristics for short-term projections from the most recent household survey to present (a period of two to five years in most countries). The five most recent years of data were excluded from each country. For a country with data up to 2020, this excluded both fertility observations 2015-2020, and retrospective data before 2015 collected in surveys since 2015. The coverage of 95% posterior predictive intervals, interval score (lower values desired) [29], continuous rank probability score (CRPS, lower values desired), and expected log posterior density (ELPD, higher values desired) were calculated based on 1000 posterior predictive samples from models calibrated excluding the held-out data. These were compared to survey-observed total fertility rates aggregated to the province level.

| **Model** | **Coverage** (95% target) | **Interval Score** (lower better) | **CRPS**  (lower better) | **ELPD**  (higher better) |
| --- | --- | --- | --- | --- |
| AR1 | 0.85 | 1127 | 68458 | 5495 |
| AR1 + trend | **0.87** | **1022** | **67907** | **5503** |
| ARIMA(1,1,0) | 0.86 | 1142 | 68564 | 5493 |
| ARIMA(1,1,0) + trend | **0.88** | **1025** | **68204** | **5502** |
| RW1 | 0.87 | 1214 | 71740 | 5504 |
| RW1 + trend | 0.87 | 1295 | 71674 | 5507 |
| RW2 | 0.87 | 1309 | 71866 | 5506 |
| RW2 + trend | 0.87 | 1280 | 71450 | 5506 |

The median coverage of 95% posterior predictive intervals for withheld province-level TFR observations was around 87% for all models. In countries with smooth fertility trends, random walk models had high interval coverage, but uncertainty ranges for short-term (3-5 year) projections became very large with biologically impossible upper bounds (for example TFR above 12). These unreasonably large projection ranges were penalised by the interval score and CRPS. On these scores, the AR1 and ARIMA(1,1,0) based models performed better than the random-walk based models. For the AR1 and ARIMA(1,1,0) models, adding a linear time trend improved interval score, CRPS, and ELPD. In several countries with declining fertility, the AR1 + trend model flattened too quickly and did not continue the declining trend, as shown in Kenya below. As the AR1 and ARIMA(1,1,0) models with a linear time trend performed similarly across all criteria, and the ARIMA(1,1,0) + trend captured declining fertility better, it was chosen as the best performing model.

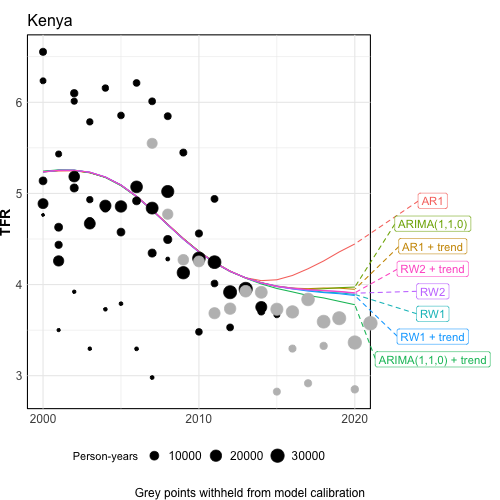

### Supplementary Table 1: Administrative levels

| **Country** | **Province (n)** | **District (n)** |
| --- | --- | --- |
| Angola | Province (18) | Municipality (164) |
| Burundi | Region (18) | District Sanitaire (49) |
| Benin | Department (12) | District (34) |
| Burkina Faso | Region (13) | District Sanitaire (70) |
| Cent. Afr. Rep. | Region sanitaires (7) | Prefecture (17) |
| Cote d’Ivoire | Region (33) | District Sanitaire (113) |
| Cameroon | Region (12) | District Sanitaire (190) |
| Dem. Rep. Congo | Territoire (164) | District Sanitaire (519) |
| Congo | Combined Region (10) | District (52) |
| Ethiopia | Region (12) | Zone (160) |
| Gabon | Province (9) | Department (51) |
| Ghana | Region (16) | District (260) |
| Guinea | Region (8) | District (38) |
| Gambia | Aggregated LGA (5) | District (42) |
| G. Bissau | Region (9) | Region (9) |
| Kenya | Former Province (8) | County (47) |
| Liberia | County (15) | Health District (136) |
| Lesotho | District (10) | Community Council (78) |
| Mali | Region (9) | Cercles (50) |
| Mozambique | Merged provinces (10) | Distrito (161) |
| Malawi | Region (3) | Health District + Cities (33) |
| Namibia | Region (14) | District (38) |
| Niger | Region (8) | Department (67) |
| Nigeria | State (37) | LGA (774) |
| Rwanda | Province (5) | District (30) |
| Senegal | Region (14) | District (79) |
| S. Leone | Province (5) | District (16) |
| S. Sudan | State (10) | State (10) |
| Eswatini | Region (4) | Tinkundla (55) |
| Chad | Region (23) | Department (70) |
| Togo | Region (6) | Prefecture (39) |
| Tanzania | Zone (7) | District (195) |
| Uganda | AIS region (10) | District (136) |
| S. Africa | Province (9) | District (52) |
| Zambia | Province (10) | District (116) |
| Zimbabwe | Province (10) | District (63) |

### Supplementary Table 2: Model calibration data

| **Country** | **Survey Year** | **Survey Type** | **Geographic resolution** | **TIPS** | **Number of births** | **Person-years** |
| --- | --- | --- | --- | --- | --- | --- |
| Angola | 2006 | MIS | Point | 5 | 2705 | 14820 |
| Angola | 2011 | MIS | Point | 5 | 18052 | 82175 |
| Angola | 2015 | DHS | Point | 15 | 30981 | 141705 |
| Burundi | 2005 | MICS | Areal | 5 | 6669 | 39787 |
| Burundi | 2010 | DHS | Point | 15 | 19213 | 92099 |
| Burundi | 2012 | MIS | Point | 5 | 5726 | 29643 |
| Burundi | 2016 | DHS | Point | 15 | 35766 | 176052 |
| Benin | 1996 | DHS | Point | 15 | 1716 | 8463 |
| Benin | 2001 | DHS | Point | 15 | 7145 | 36236 |
| Benin | 2006 | DHS | Point | 15 | 34733 | 164067 |
| Benin | 2012 | DHS | Point | 15 | 34630 | 180534 |
| Benin | 2014 | MICS | Areal | 5 | 12134 | 78860 |
| Benin | 2017 | DHS | Point | 15 | 33683 | 165056 |
| Burkina Faso | 1999 | DHS | Point | 15 | 5078 | 23739 |
| Burkina Faso | 2003 | DHS | Point | 15 | 19094 | 87678 |
| Burkina Faso | 2006 | MICS | Areal | 5 | 5012 | 34666 |
| Burkina Faso | 2010 | DHS | Point | 15 | 41349 | 181936 |
| Burkina Faso | 2014 | MIS | Point | 5 | 8490 | 47110 |
| Burkina Faso | 2017 | MIS | Point | 5 | 6815 | 39760 |
| Burkina Faso | 2021 | DHS | Point | 15 | 34445 | 184899 |
| Cent. Afr. Rep. | 2006 | MICS | Areal | 5 | 7930 | 53221 |
| Cent. Afr. Rep. | 2010 | MICS | Areal | 5 | 9437 | 57927 |
| Cent. Afr. Rep. | 2018 | MICS | Areal | 5 | 7738 | 40740 |
| Cote d’Ivoire | 1998 | DHS | Point | 15 | 1811 | 10490 |
| Cote d’Ivoire | 2005 | AIS | Areal | 5 | 7292 | 41515 |
| Cote d’Ivoire | 2006 | MICS | Areal | 5 | 7893 | 64363 |
| Cote d’Ivoire | 2012 | DHS | Point | 15 | 19359 | 105278 |
| Cote d’Ivoire | 2017 | PHIA | Point | 1 | 1111 | 8256 |
| Cameroon | 1998 | DHS | Areal | 15 | 2721 | 16730 |
| Cameroon | 2000 | MICS | Areal | 5 | 2260 | 18176 |
| Cameroon | 2004 | DHS | Point | 15 | 14290 | 76511 |
| Cameroon | 2006 | MICS | Areal | 5 | 5709 | 42246 |
| Cameroon | 2011 | DHS | Point | 15 | 29686 | 154840 |
| Cameroon | 2014 | MICS | Areal | 15 | 19263 | 101438 |
| Cameroon | 2017 | PHIA | Point | 1 | 1701 | 12326 |
| Cameroon | 2018 | DHS | Point | 15 | 25564 | 146274 |
| Dem. Rep. Congo | 2007 | DHS | Point | 15 | 19357 | 91276 |
| Dem. Rep. Congo | 2013 | DHS | Point | 15 | 43777 | 192882 |
| Dem. Rep. Congo | 2017 | MICS | Areal | 15 | 43745 | 214905 |
| Congo | 2005 | DHS | Point | 15 | 9392 | 57202 |
| Congo | 2011 | DHS | Point | 15 | 19431 | 113980 |
| Congo | 2014 | MICS | Areal | 5 | 7085 | 51085 |
| Ethiopia | 2005 | DHS | Point | 15 | 23782 | 111753 |
| Ethiopia | 2011 | DHS | Point | 15 | 33638 | 164043 |
| Ethiopia | 2016 | DHS | Point | 15 | 32288 | 162093 |
| Ethiopia | 2019 | DHS | Point | 15 | 16039 | 88120 |
| Gabon | 2000 | DHS | Point | 15 | 4545 | 30147 |
| Gabon | 2012 | DHS | Point | 15 | 12705 | 87634 |
| Gabon | 2019 | DHS | Point | 15 | 14103 | 112295 |
| Ghana | 1998 | DHS | Point | 15 | 2580 | 17688 |
| Ghana | 2003 | DHS | Point | 15 | 6241 | 40745 |
| Ghana | 2008 | DHS | Point | 15 | 7352 | 49102 |
| Ghana | 2014 | DHS | Point | 15 | 15367 | 103938 |
| Ghana | 2016 | MIS | Point | 5 | 3613 | 26940 |
| Ghana | 2019 | MIS | Point | 5 | 3332 | 27076 |
| Guinea | 1999 | DHS | Point | 15 | 5502 | 27379 |
| Guinea | 2005 | DHS | Point | 15 | 14133 | 64361 |
| Guinea | 2012 | DHS | Point | 15 | 19197 | 93935 |
| Guinea | 2018 | DHS | Point | 15 | 20521 | 109906 |
| Guinea | 2021 | MIS | Point | 5 | 4489 | 29042 |
| Gambia | 2005 | MICS | Areal | 5 | 5815 | 43765 |
| Gambia | 2010 | MICS | Areal | 5 | 10438 | 69311 |
| Gambia | 2013 | DHS | Point | 15 | 19110 | 102107 |
| Gambia | 2018 | MICS | Areal | 5 | 8446 | 62978 |
| Gambia | 2019 | DHS | Point | 15 | 20752 | 120623 |
| G. Bissau | 2014 | MICS | Areal | 15 | 18646 | 104394 |
| G. Bissau | 2018 | MICS | Areal | 15 | 18241 | 112666 |
| Kenya | 1998 | DHS | Areal | 15 | 3803 | 24267 |
| Kenya | 2003 | DHS | Point | 15 | 9769 | 56248 |
| Kenya | 2008 | DHS | Point | 15 | 14887 | 83530 |
| Kenya | 2009 | MICS | Areal | 15 | 1086 | 7819 |
| Kenya | 2011 | MICS | Areal | 15 | 12872 | 61112 |
| Kenya | 2014 | DHS | Point | 15 | 54897 | 333434 |
| Kenya | 2015 | MIS | Point | 5 | 3612 | 27161 |
| Kenya | 2018 | PHIA | Point | 1 | 1653 | 14296 |
| Kenya | 2020 | MIS | Point | 5 | 3887 | 35317 |
| Kenya | 2022 | DHS | Point | 15 | 46992 | 335722 |
| Liberia | 2007 | DHS | Point | 15 | 12608 | 66646 |
| Liberia | 2009 | MIS | Point | 5 | 9486 | 45047 |
| Liberia | 2011 | MIS | Point | 5 | 3539 | 23369 |
| Liberia | 2013 | DHS | Point | 15 | 18165 | 95652 |
| Liberia | 2016 | MIS | Point | 5 | 3163 | 21885 |
| Liberia | 2019 | DHS | Point | 15 | 14605 | 84937 |
| Lesotho | 2004 | DHS | Point | 15 | 6714 | 53973 |
| Lesotho | 2009 | DHS | Point | 15 | 9328 | 76608 |
| Lesotho | 2014 | DHS | Point | 15 | 8091 | 67760 |
| Lesotho | 2017 | PHIA | Point | 1 | 594 | 6369 |
| Lesotho | 2018 | MICS | Areal | 15 | 7274 | 69410 |
| Mali | 1996 | DHS | Point | 15 | 2536 | 10644 |
| Mali | 2001 | DHS | Point | 15 | 16575 | 70225 |
| Mali | 2006 | DHS | Point | 15 | 31666 | 127136 |
| Mali | 2009 | MICS | Areal | 5 | 20591 | 115147 |
| Mali | 2012 | DHS | Point | 15 | 26667 | 112020 |
| Mali | 2015 | MICS | Areal | 5 | 15289 | 91891 |
| Mali | 2015 | MIS | Point | 5 | 8966 | 45407 |
| Mali | 2018 | DHS | Point | 15 | 26878 | 111399 |
| Mali | 2021 | MIS | Point | 5 | 10534 | 55170 |
| Mozambique | 1997 | DHS | Areal | 15 | 3540 | 19695 |
| Mozambique | 2003 | DHS | Areal | 15 | 18359 | 90596 |
| Mozambique | 2008 | MICS | Areal | 15 | 30563 | 143394 |
| Mozambique | 2011 | DHS | Point | 15 | 28893 | 142326 |
| Mozambique | 2015 | AIS | Point | 5 | 6530 | 41448 |
| Mozambique | 2018 | MIS | Point | 5 | 5297 | 29763 |
| Malawi | 2000 | DHS | Point | 15 | 13457 | 66016 |
| Malawi | 2004 | DHS | Point | 15 | 19211 | 90868 |
| Malawi | 2006 | MICS | Areal | 15 | 51048 | 230219 |
| Malawi | 2010 | DHS | Point | 15 | 51532 | 235083 |
| Malawi | 2012 | MIS | Point | 5 | 2813 | 16158 |
| Malawi | 2013 | MICS | Areal | 15 | 52331 | 250484 |
| Malawi | 2014 | MIS | Point | 5 | 2547 | 16558 |
| Malawi | 2015 | DHS | Point | 15 | 48744 | 251652 |
| Malawi | 2017 | MIS | Point | 5 | 2838 | 18268 |
| Malawi | 2017 | PHIA | Point | 1 | 1332 | 9819 |
| Malawi | 2019 | MICS | Areal | 15 | 39815 | 220797 |
| Namibia | 2000 | DHS | Point | 15 | 4590 | 34325 |
| Namibia | 2006 | DHS | Point | 15 | 11005 | 86466 |
| Namibia | 2013 | DHS | Point | 15 | 13226 | 105146 |
| Namibia | 2017 | PHIA | Point | 1 | 1093 | 8633 |
| Niger | 1998 | DHS | Point | 15 | 5545 | 23503 |
| Niger | 2006 | DHS | Areal | 15 | 22289 | 80549 |
| Niger | 2012 | DHS | Areal | 15 | 35717 | 121311 |
| Niger | 2021 | MIS | Point | 5 | 5804 | 28421 |
| Nigeria | 2003 | DHS | Point | 15 | 10242 | 51446 |
| Nigeria | 2007 | MICS | Areal | 5 | 15183 | 117630 |
| Nigeria | 2008 | DHS | Point | 15 | 68082 | 331060 |
| Nigeria | 2010 | MIS | Point | 5 | 14668 | 68433 |
| Nigeria | 2011 | MICS | Areal | 5 | 22947 | 145231 |
| Nigeria | 2013 | DHS | Point | 15 | 84422 | 410079 |
| Nigeria | 2015 | MIS | Point | 5 | 7554 | 41964 |
| Nigeria | 2016 | MICS | Areal | 15 | 78102 | 364737 |
| Nigeria | 2018 | DHS | Point | 15 | 90210 | 445787 |
| Nigeria | 2021 | MIS | Point | 5 | 11966 | 74623 |
| Rwanda | 2000 | DHS | Areal | 15 | 9251 | 50882 |
| Rwanda | 2005 | DHS | Point | 15 | 17219 | 89402 |
| Rwanda | 2008 | DHS | Point | 15 | 13136 | 71016 |
| Rwanda | 2010 | DHS | Point | 15 | 24515 | 139874 |
| Rwanda | 2013 | MIS | Areal | 5 | 3871 | 27500 |
| Rwanda | 2015 | DHS | Point | 15 | 22801 | 142845 |
| Rwanda | 2017 | MIS | Areal | 5 | 3419 | 26165 |
| Rwanda | 2019 | DHS | Point | 15 | 21914 | 150695 |
| Rwanda | 2019 | PHIA | Point | 1 | 1635 | 13372 |
| Senegal | 1997 | DHS | Point | 15 | 3301 | 18127 |
| Senegal | 2005 | DHS | Point | 15 | 20565 | 113347 |
| Senegal | 2006 | MIS | Areal | 5 | 4477 | 29106 |
| Senegal | 2008 | MIS | Point | 5 | 33321 | 185519 |
| Senegal | 2010 | DHS | Point | 15 | 28411 | 158236 |
| Senegal | 2012 | DHS | Point | 15 | 15412 | 84533 |
| Senegal | 2014 | DHS | Point | 15 | 15376 | 87576 |
| Senegal | 2015 | DHS | Point | 15 | 16271 | 90515 |
| Senegal | 2016 | DHS | Point | 15 | 15926 | 91124 |
| Senegal | 2017 | DHS | Areal | 15 | 30177 | 173970 |
| Senegal | 2019 | DHS | Point | 15 | 15216 | 89104 |
| S. Leone | 2008 | DHS | Point | 15 | 15006 | 75471 |
| S. Leone | 2010 | MICS | Areal | 5 | 7395 | 58981 |
| S. Leone | 2013 | DHS | Point | 15 | 34078 | 170920 |
| S. Leone | 2017 | MICS | Areal | 5 | 9876 | 86677 |
| S. Leone | 2019 | DHS | Point | 15 | 27452 | 161936 |
| S. Sudan | 2010 | MICS | Areal | 15 | 24325 | 101635 |
| Eswatini | 2000 | MICS | Areal | 5 | 2410 | 21199 |
| Eswatini | 2006 | DHS | Point | 15 | 6154 | 42014 |
| Eswatini | 2010 | MICS | Areal | 15 | 6663 | 47277 |
| Eswatini | 2014 | MICS | Areal | 15 | 6493 | 49639 |
| Eswatini | 2017 | PHIA | Point | 1 | 453 | 4957 |
| Chad | 2004 | DHS | Areal | 15 | 11610 | 46355 |
| Chad | 2010 | MICS | Areal | 5 | 15380 | 74669 |
| Chad | 2014 | DHS | Point | 15 | 50665 | 179615 |
| Chad | 2019 | MICS | Areal | 5 | 17357 | 95120 |
| Togo | 1998 | DHS | Point | 15 | 4346 | 26265 |
| Togo | 2006 | MICS | Areal | 5 | 1978 | 19381 |
| Togo | 2010 | MICS | Areal | 5 | 4086 | 33059 |
| Togo | 2013 | DHS | Point | 15 | 17999 | 102548 |
| Togo | 2017 | MICS | Areal | 5 | 4495 | 36518 |
| Togo | 2017 | MIS | Point | 5 | 3724 | 24532 |
| Tanzania | 1996 | DHS | Areal | 15 | 2623 | 13497 |
| Tanzania | 1999 | DHS | Point | 15 | 3190 | 17093 |
| Tanzania | 2004 | DHS | Areal | 15 | 15940 | 80356 |
| Tanzania | 2007 | AIS | Point | 5 | 17378 | 88266 |
| Tanzania | 2010 | DHS | Point | 15 | 20960 | 105487 |
| Tanzania | 2012 | AIS | Point | 5 | 10745 | 66438 |
| Tanzania | 2015 | DHS | Point | 15 | 25611 | 137524 |
| Tanzania | 2017 | MIS | Point | 5 | 8401 | 51976 |
| Tanzania | 2017 | PHIA | Point | 1 | 2256 | 14755 |
| Tanzania | 2022 | DHS | Point | 15 | 27235 | 156974 |
| Uganda | 2000 | DHS | Point | 15 | 8829 | 37246 |
| Uganda | 2006 | DHS | Point | 15 | 18041 | 73035 |
| Uganda | 2009 | MIS | Point | 5 | 10152 | 41414 |
| Uganda | 2014 | MIS | Point | 5 | 5901 | 30927 |
| Uganda | 2016 | DHS | Point | 15 | 40141 | 186731 |
| Uganda | 2017 | PHIA | Point | 1 | 2288 | 14103 |
| Uganda | 2018 | MIS | Point | 5 | 7922 | 45509 |
| S. Africa | 1998 | DHS | Areal | 15 | 3383 | 36160 |
| S. Africa | 2016 | DHS | Point | 15 | 9527 | 95652 |
| Zambia | 1996 | DHS | Point | 15 | 2713 | 13728 |
| Zambia | 2002 | DHS | Point | 15 | 9311 | 45609 |
| Zambia | 2007 | DHS | Point | 15 | 14011 | 65479 |
| Zambia | 2013 | DHS | Point | 15 | 35537 | 169200 |
| Zambia | 2016 | PHIA | Point | 1 | 1584 | 10704 |
| Zambia | 2018 | DHS | Point | 15 | 26730 | 140473 |
| Zimbabwe | 1999 | DHS | Point | 15 | 3447 | 24920 |
| Zimbabwe | 2005 | DHS | Point | 15 | 10435 | 71863 |
| Zimbabwe | 2009 | MICS | Areal | 15 | 16269 | 110993 |
| Zimbabwe | 2010 | DHS | Point | 15 | 13665 | 93932 |
| Zimbabwe | 2014 | MICS | Areal | 15 | 23493 | 152131 |
| Zimbabwe | 2015 | DHS | Point | 15 | 15923 | 104486 |
| Zimbabwe | 2016 | PHIA | Point | 1 | 1237 | 10632 |
| Zimbabwe | 2019 | MICS | Areal | 15 | 16216 | 108156 |

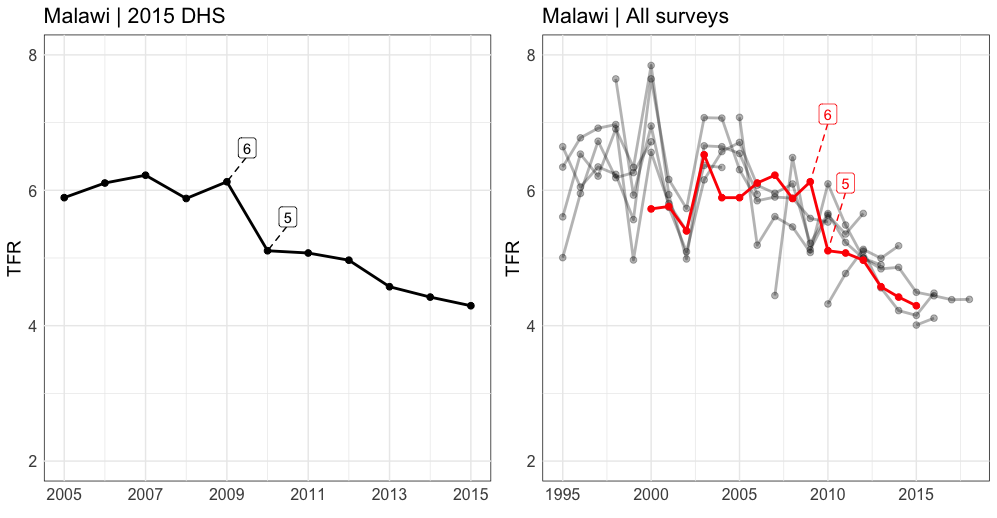

**Supplementary Figure S1: Identification of TIPS coefficients and birth displacement bias**

The Malawi 2015 DHS estimated a TFR of 5 in 2010, five years preceding the survey (TIPS = 5). The TFR estimate six years before the survey was 6, an unexpectedly large change for a single calendar year. The TFR estimate of 6 in 2009 has been overestimated due to births that have been ‘displaced’ over the five year threshold (left panel).

The TIPS fixed and random effects cannot be identified from a single survey because it is not possible to distinguish a ‘true’ large fertility change from reporting bias. We used overlapping survey periods to identify these effects (right panel, 2015 DHS shown in red). TIPS indices may span more than one calendar year (1 year preceding a survey conducted in July of a given year contains 6 months of one year, and six months of the preceding year). In 2009, when the 2015 survey was exposed to birth displacement bias (TIPS = 6), there were also estimates from the 2010 DHS (TIPS = 0 and 1), 2012 MIS (TIPS = 2 and 3), 2013 MICS (TIPS = 4 and 5), and the 2019 MICS (TIPS = 10 and 11). These surveys were not exposed to systematic birth displacement bias due to the 5-year reporing threshold, and therefore we are able to estimate the observation-level deviation of the 2015 DHS from the process-level observation of total fertility.

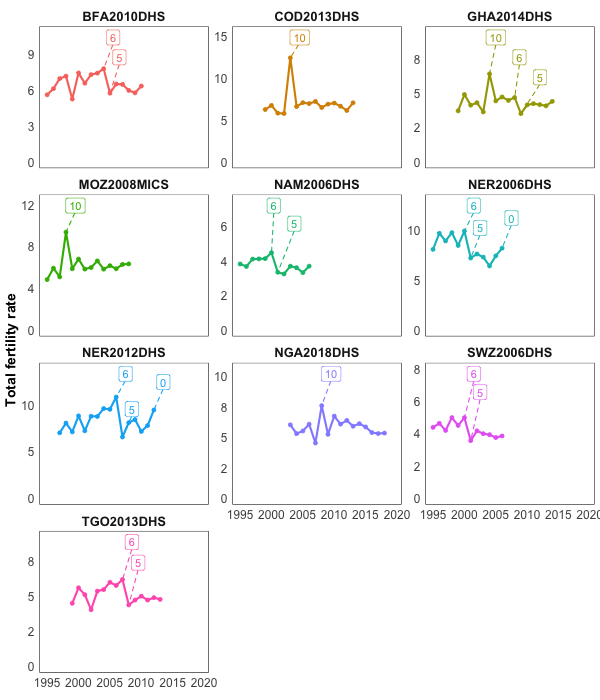

**Supplementary Figure S2: Bias and data heaping in household survey data.**

Time Preceding Survey (TIPS) labelled for 0 (year of survey), 5 and 6 (birth displacement) and 10 years (round year heaping).

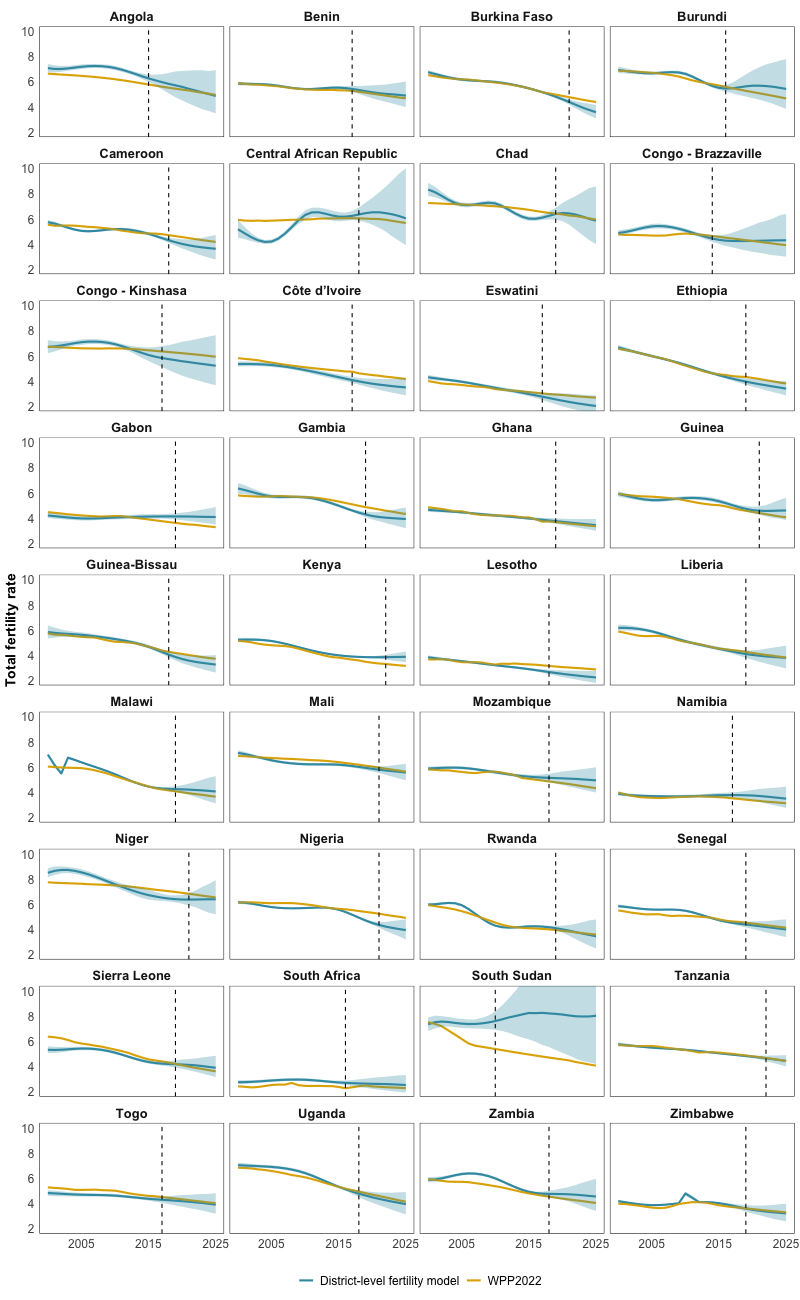

**Supplementary Figure S3: Comparison of national-level total fertility rate estimates between the district-level fertility model and UN Population Division World Population Prospects 2022.** Dotted line represents the year of most recent survey.
