## Supplementary File 2 for "Small-area estimation of district-level fertility in 36 countries in sub-Saharan Africa 2000-2025"

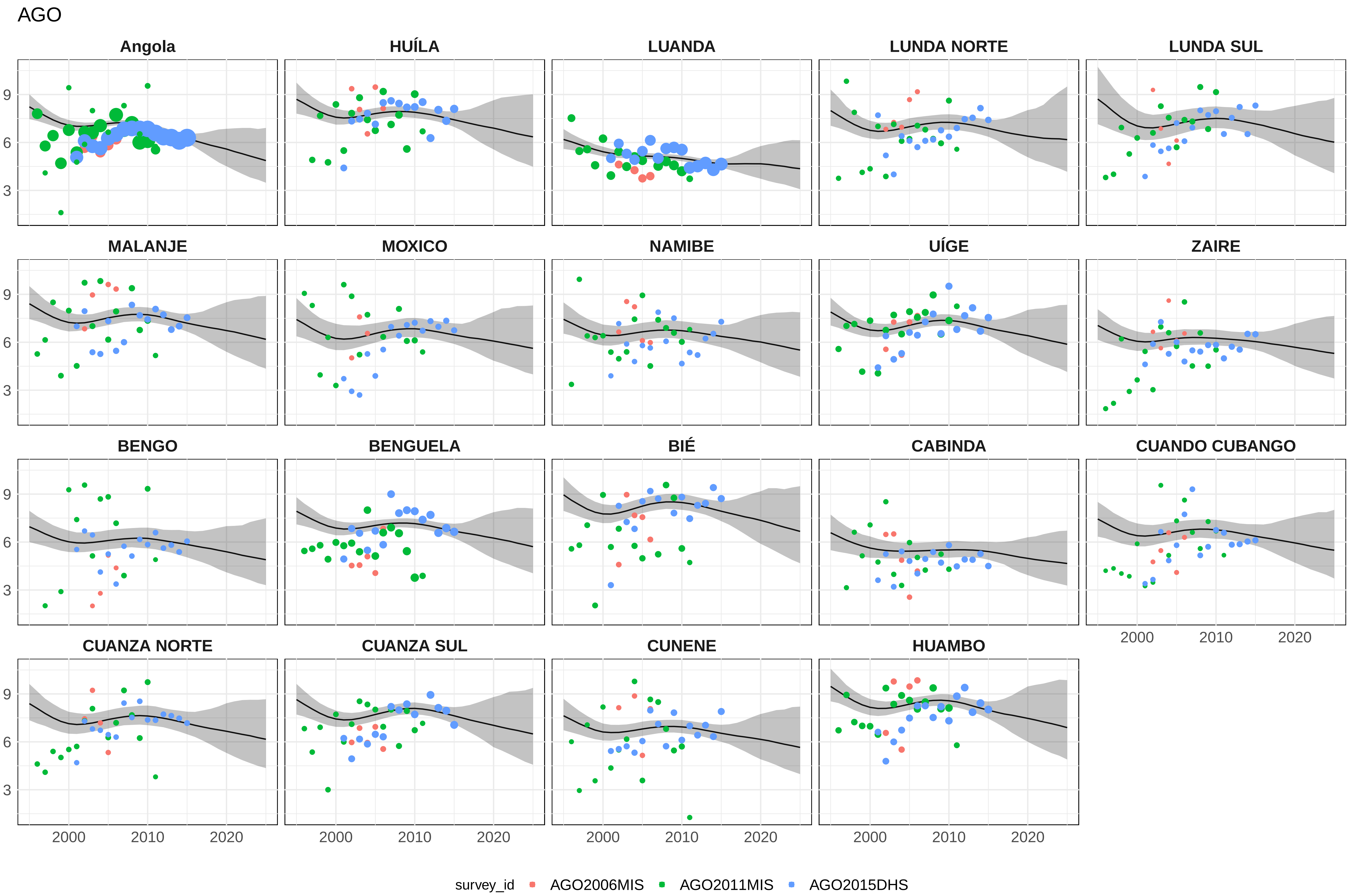

AGO
Angola
HUÍLA
LUANDA
LUNDA NORTE
LUNDA SUL
MALANJE
MOXICO
NAMIBE
UÍGE
ZAIRE
BENGO
BENGUELA
BIÉ
CABINDA
CUANDO CUBANGO
CUANZA NORTE
CUANZA SUL
CUNENE
HUAMBO
3
6
9
2000
2010
2020
survey_id
AGO2006MIS
AGO2011MIS
AGO2015DHS

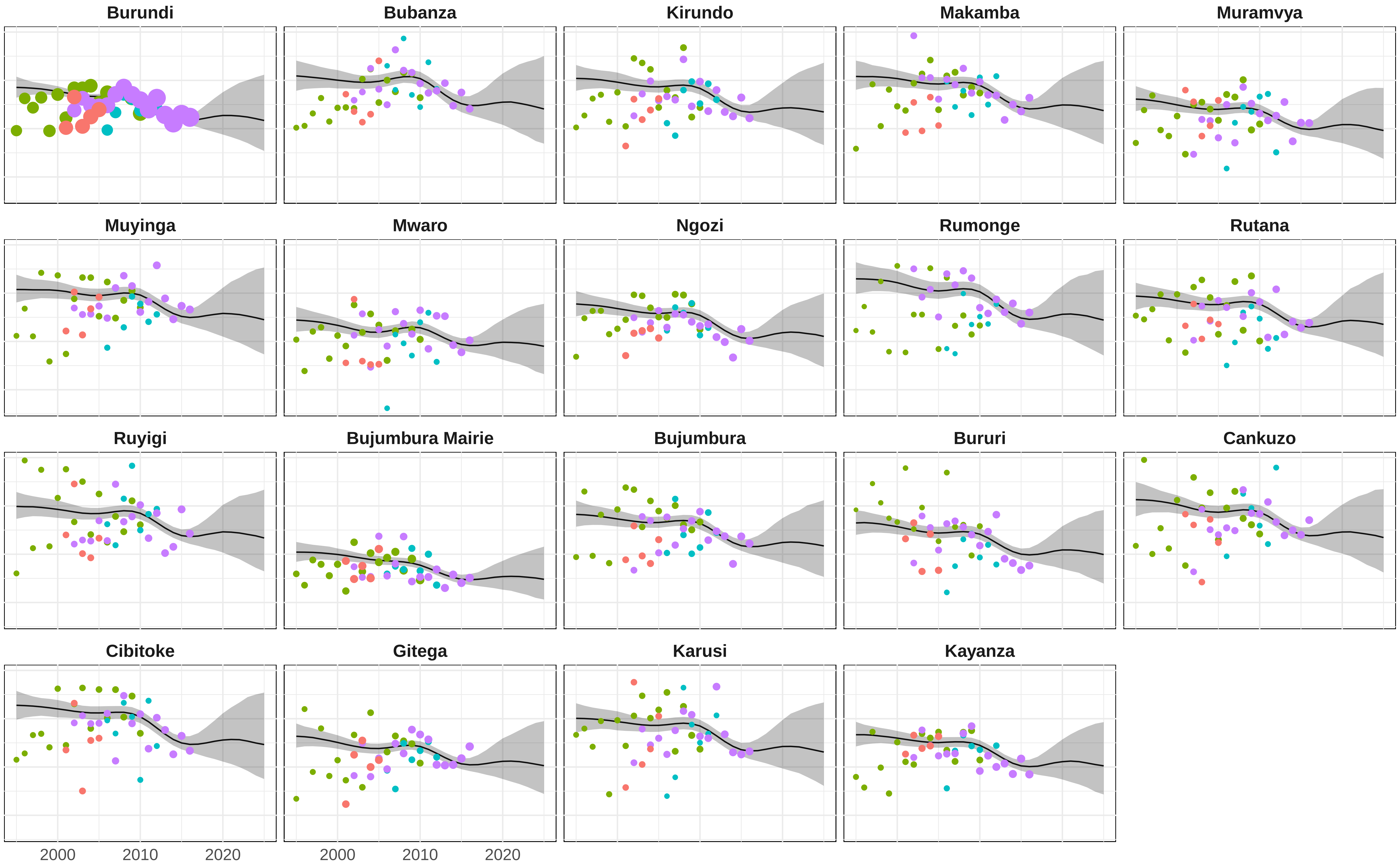
Burundi
Bubanza
Kirundo
Makamba
Muramvya
Muyinga
Mwaro
Ngozi
Rumonge
Rutana
Ruyigi
Bujumbura Mairie
Bujumbura
Bururi
Cankuzo
Cibitoke
Gitega
Karusi
Kayanza
2000
2010
2020
2000
2010
2020

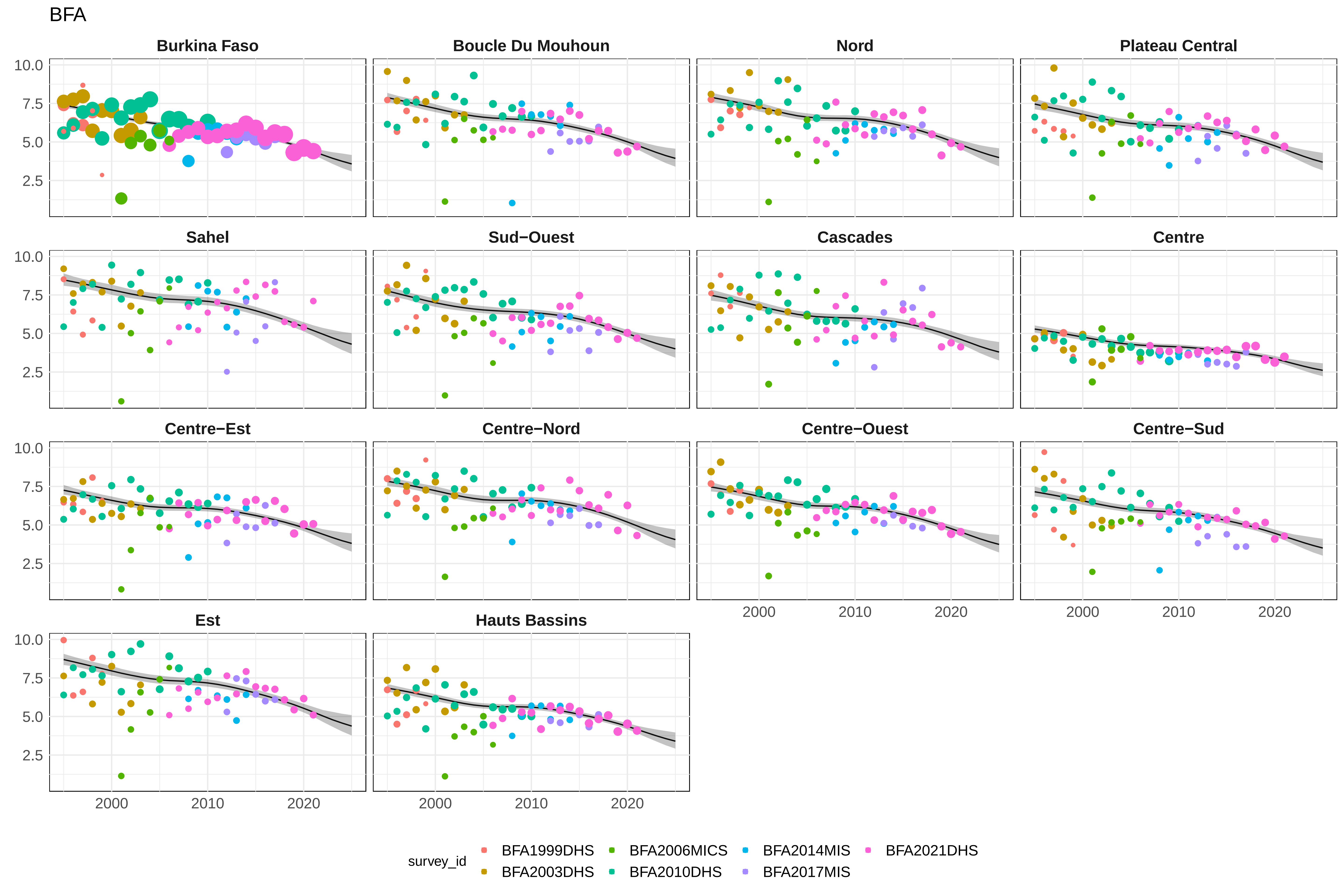

BFA
Burkina Faso
Boucle Du Mouhoun
Nord
Plateau Central
Sahel
Sud−Ouest
Cascades
Centre
Centre−Est
Centre−Nord
Centre−Ouest
Centre−Sud
Est
Hauts Bassins
10.0
7.5
5.0
2.5
2000
2010
2020
survey_id
BFA1999DHS
BFA2003DHS
BFA2006MICS
BFA2010DHS
BFA2014MIS
BFA2017MIS
BFA2021DHS

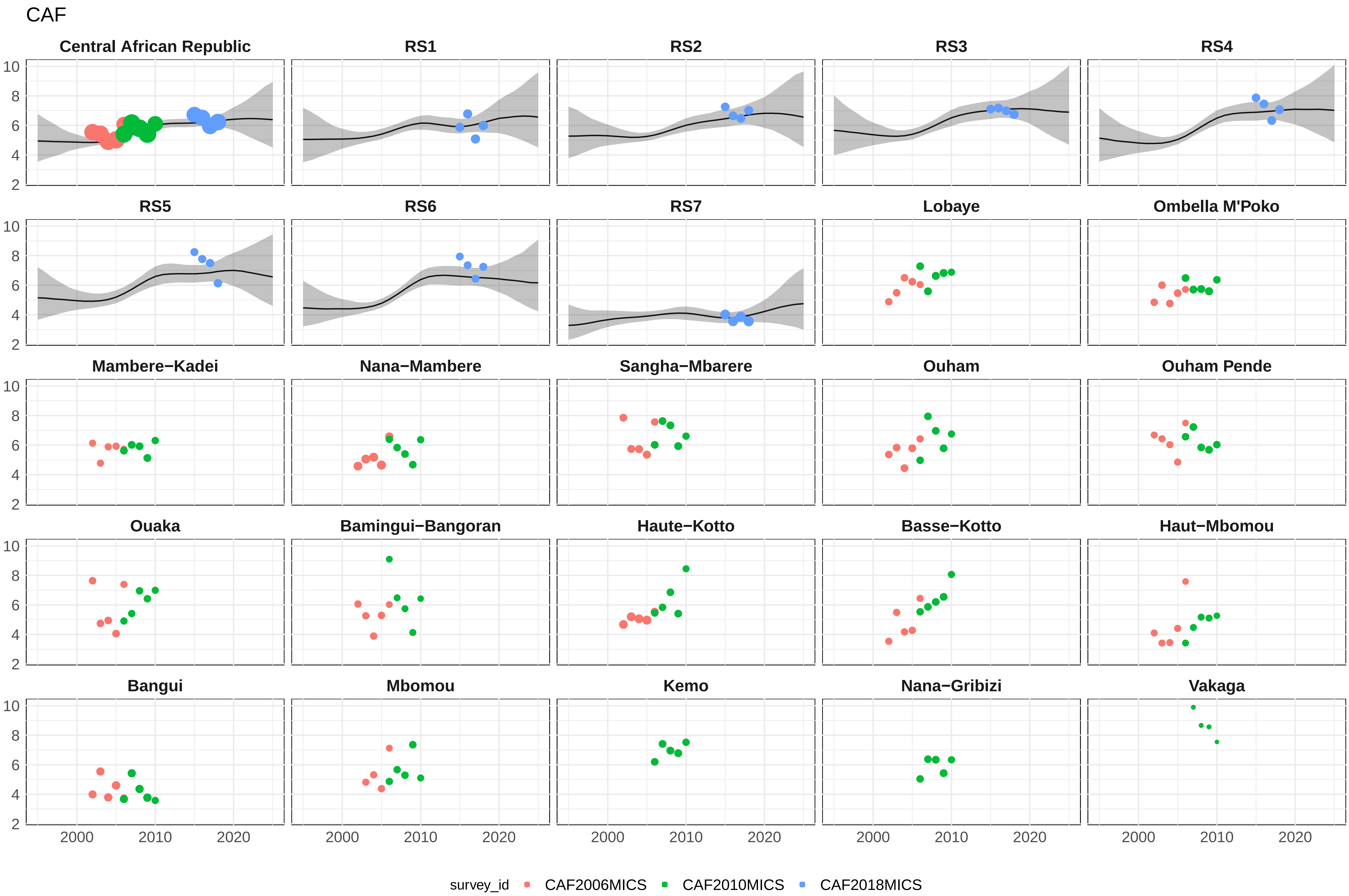

CAF
Central African Republic
RS1
RS2
RS3
RS4
RS5
RS6
RS7
Lobaye
Ombella M'Poko
Mambere−Kadei
Nana−Mambere
Sangha−Mbarere
Ouham
Ouham Pende
Ouaka
Bamingui−Bangoran
Haute−Kotto
Basse−Kotto
Haut−Mbomou
Bangui
Mbomou
Kemo
Nana−Gribizi
Vakaga
2
4
6
8
10
2000
2010
2020
survey_id
CAF2006MICS
CAF2010MICS
CAF2018MICS

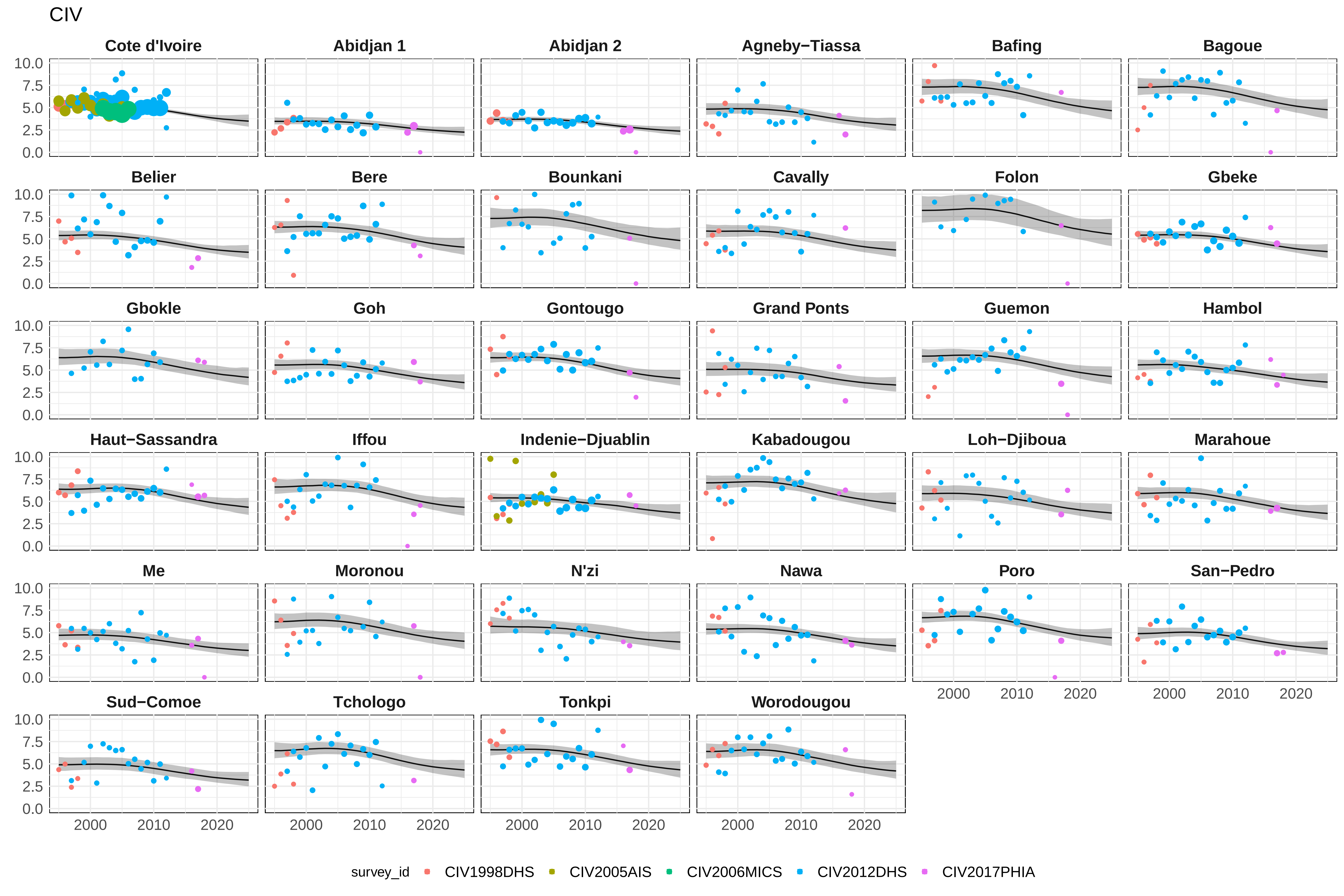
CIV
Cote d'Ivoire
Abidjan 1
Abidjan 2
Agneby−Tiassa
Bafing
Bagoue
Belier
Bere
Bounkani
Cavally
Folon
Gbeke
Gbokle
Goh
Gontougo
Grand Ponts
Guemon
Hambol
Haut−Sassandra
Iffou
Indenie−Djuablin
Kabadougou
Loh−Djiboua
Marahoue
Me
Moronou
N'zi
Nawa
Poro
San−Pedro
Sud−Comoe
Tchologo
Tonkpi
Worodougou
0.0
2.5
5.0
7.5
10.0
2000
2010
2020
survey_id
CIV1998DHS
CIV2005AIS
CIV2006MICS
CIV2012DHS
CIV2017PHIA

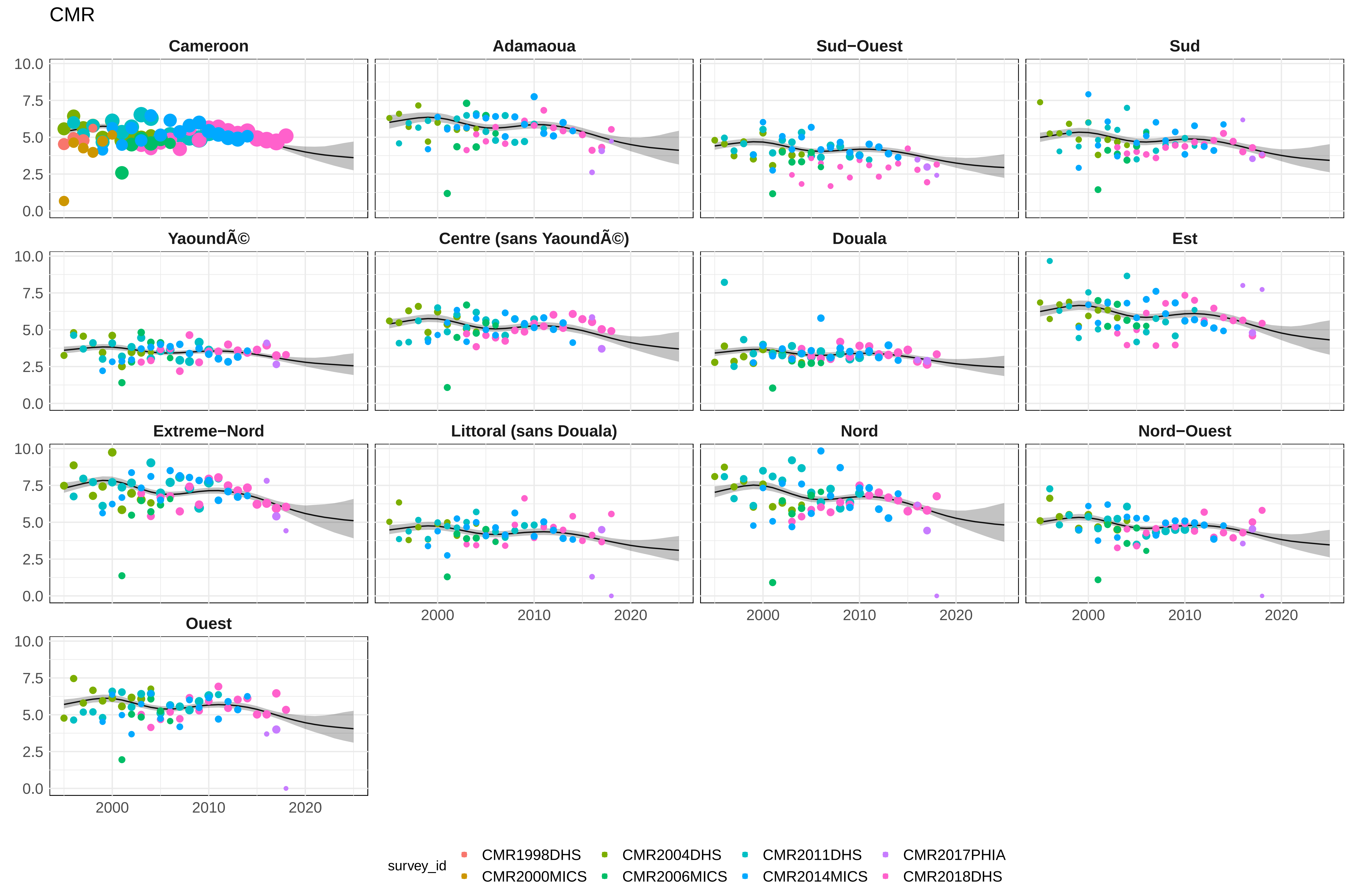

CMR
Cameroon
Adamaoua
Sud−Ouest
Sud
YaoundÃ©
Centre (sans YaoundÃ©)
Douala
Est
Extreme−Nord
Littoral (sans Douala)
Nord
Nord−Ouest
Ouest
10.0
7.5
5.0
2.5
0.0
2000
2010
2020
survey_id
CMR1998DHS
CMR2000MICS
CMR2004DHS
CMR2006MICS
CMR2011DHS
CMR2014MICS
CMR2017PHIA
CMR2018DHS

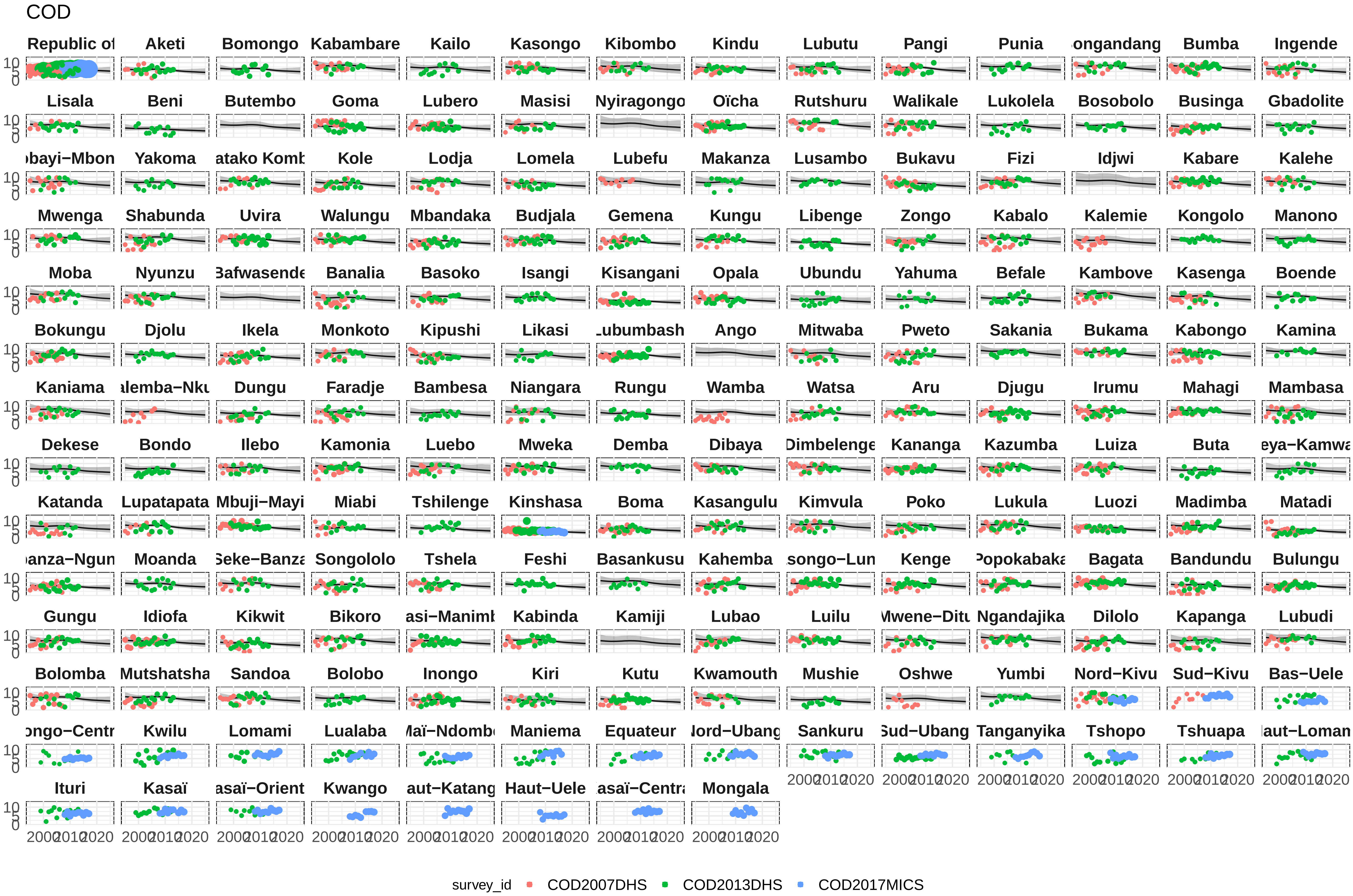
COD
Republic of
Aketi
Bomongo
Kabambare
Kailo
Kasongo
Kibombo
Kindu
Lubutu
Pangi
Punia
ongandang
Bumba
Ingende
Lisala
Beni
Butembo
Goma
Lubero
Masisi
Nyiragongo
Oïcha
Rutshuru
Walikale
Lukolela
Bosobolo
Businga
Gbadolite
bayi−Mbon
Yakoma
atako Komb
Kole
Lodja
Lomela
Lubefu
Makanza
Lusambo
Bukavu
Fizi
Idjwi
Kabare
Kalehe
Mwenga
Shabunda
Uvira
Walungu
Mbandaka
Budjala
Gemena
Kungu
Libenge
Zongo
Kabalo
Kalemie
Kongolo
Manono
Moba
Nyunzu
Bafwasende
Banalia
Basoko
Isangi
Kisangani
Opala
Ubundu
Yahuma
Befale
Kambove
Kasenga
Boende
Bokungu
Djolu
Ikela
Monkoto
Kipushi
Likasi
ubumbash
Ango
Mitwaba
Pweto
Sakania
Bukama
Kabongo
Kamina
Kaniama
alemba−Nku
Dungu
Faradje
Bambesa
Niangara
Rungu
Wamba
Watsa
Aru
Djugu
Irumu
Mahagi
Mambasa
Dekese
Bondo
Ilebo
Kamonia
Luebo
Mweka
Demba
Dibaya
Dimbelenge
Kananga
Kazumba
Luiza
Buta
eya−Kamwa
Katanda
Lupatapata
Mbuji−Mayi
Miabi
Tshilenge
Kinshasa
Boma
Kasangulu
Kimvula
Poko
Lukula
Luozi
Madimba
Matadi
anza−Ngun
Moanda
Seke−Banza
Songololo
Tshela
Feshi
Basankusu
Kahemba
songo−Lun
Kenge
Popokabaka
Bagata
Bandundu
Bulungu
Gungu
Idiofa
Kikwit
Bikoro
asi−Manimb
Kabinda
Kamiji
Lubao
Luilu
Mwene−Ditu
Ngandajika
Dilolo
Kapanga
Lubudi
Bolomba
Mutshatsha
Sandoa
Bolobo
Inongo
Kiri
Kutu
Kwamouth
Mushie
Oshwe
Yumbi
Nord−Kivu
Sud−Kivu
Bas−Uele
ongo−Centr
Kwilu
Lomami
Lualaba
Maï−Ndombe
Maniema
Equateur
Nord−Ubang
Sankuru
Sud−Ubang
Tanganyika
Tshopo
Tshuapa
Haut−Loman
Ituri
Kasaï
Kasaï−Orient
Kwango
Haut−Katang
Haut−Uele
Kasaï−Centr
Mongala
10
5
0
2000
2010
2020
survey_id
COD2007DHS
COD2013DHS
COD2017MICS

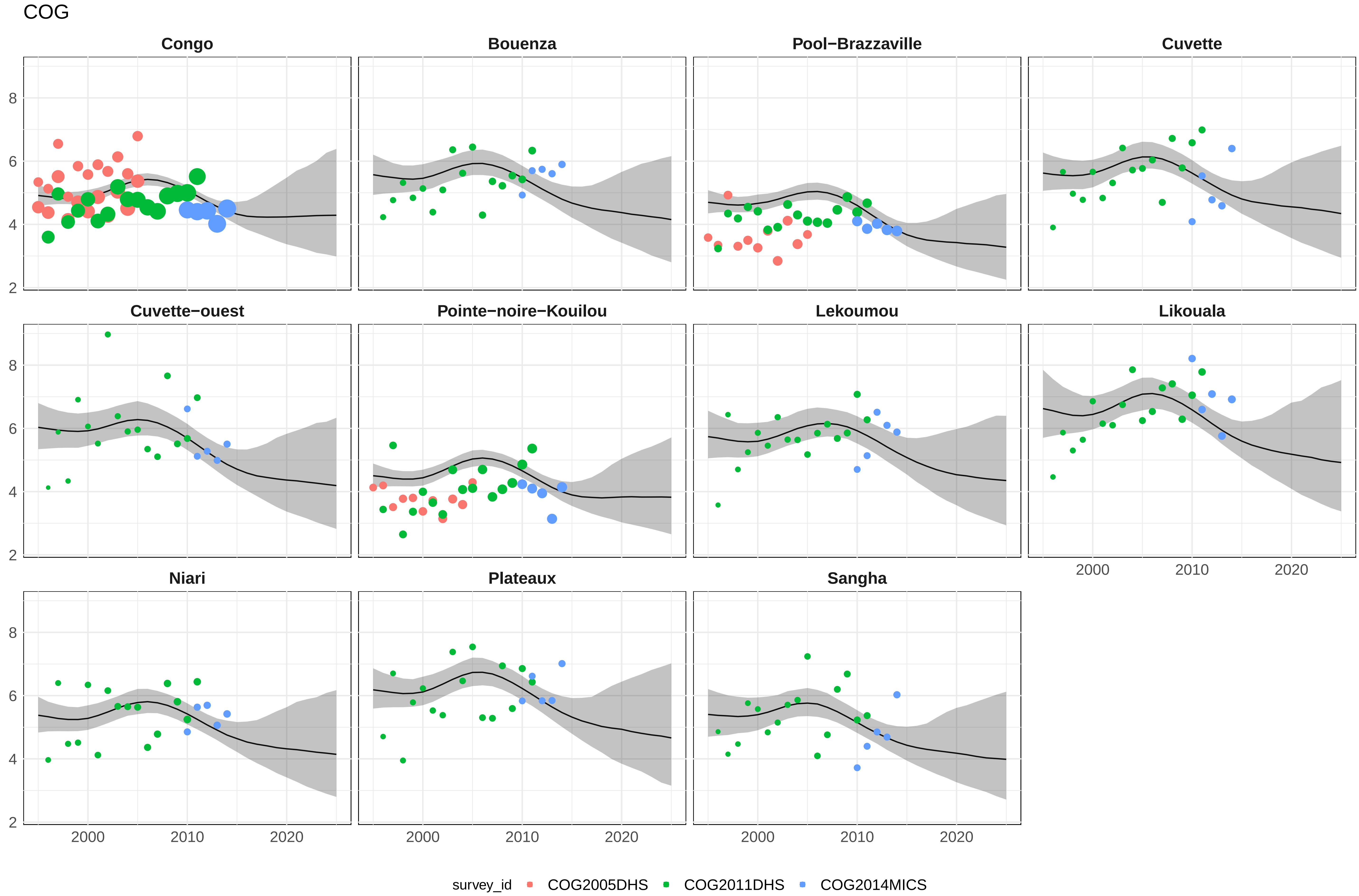
COG
Congo
Bouenza
Pool−Brazzaville
Cuvette
Cuvette−ouest
Pointe−noire−Kouilou
Lekoumou
Likouala
Niari
Plateaux
Sangha
2
4
6
8
2000
2010
2020
survey_id
COG2005DHS
COG2011DHS
COG2014MICS

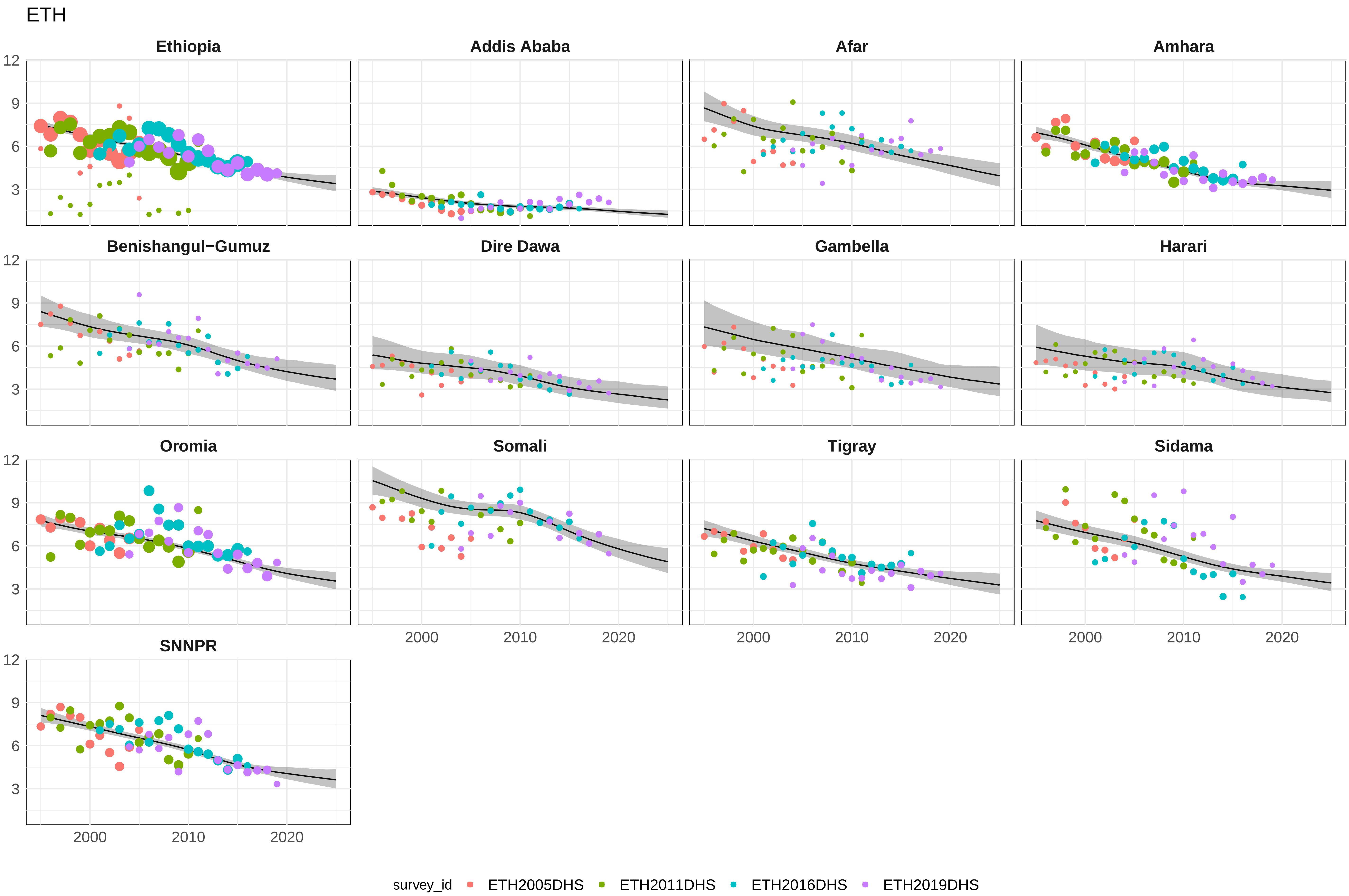
ETH
Ethiopia
Addis Ababa
Afar
Amhara
Benishangul−Gumuz
Dire Dawa
Gambella
Harari
Oromia
Somali
Tigray
Sidama
SNNPR
12
9
6
3
2000
2010
2020
survey_id
ETH2005DHS
ETH2011DHS
ETH2016DHS
ETH2019DHS

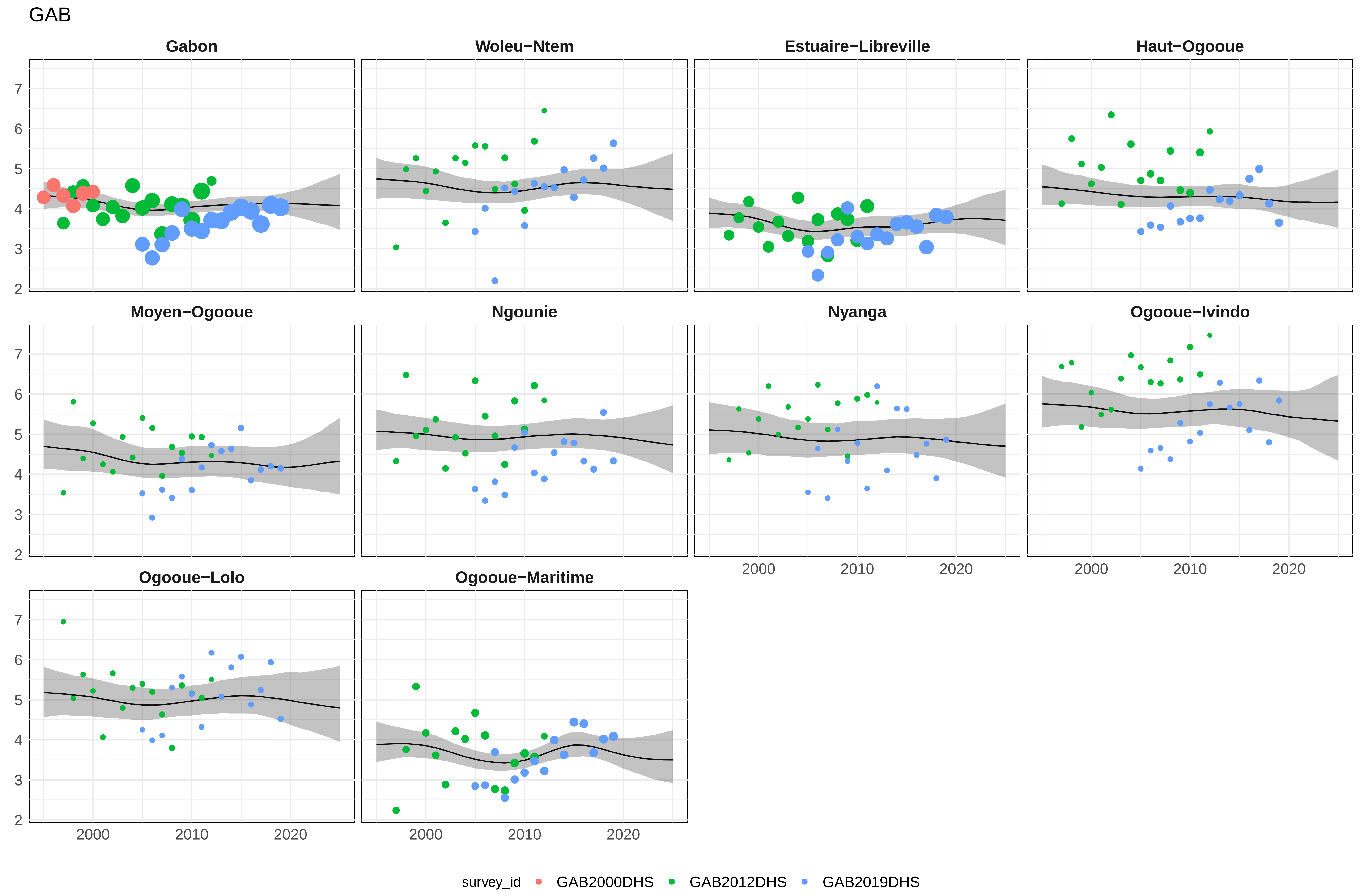

GAB
Gabon
Woleu−Ntem
Estuaire−Libreville
Haut−Ogooue
Moyen−Ogooue
Ngounie
Nyanga
Ogooue−Ivindo
Ogooue−Lolo
Ogooue−Maritime
2
3
4
5
6
7
2000
2010
2020
survey_id
GAB2000DHS
GAB2012DHS
GAB2019DHS

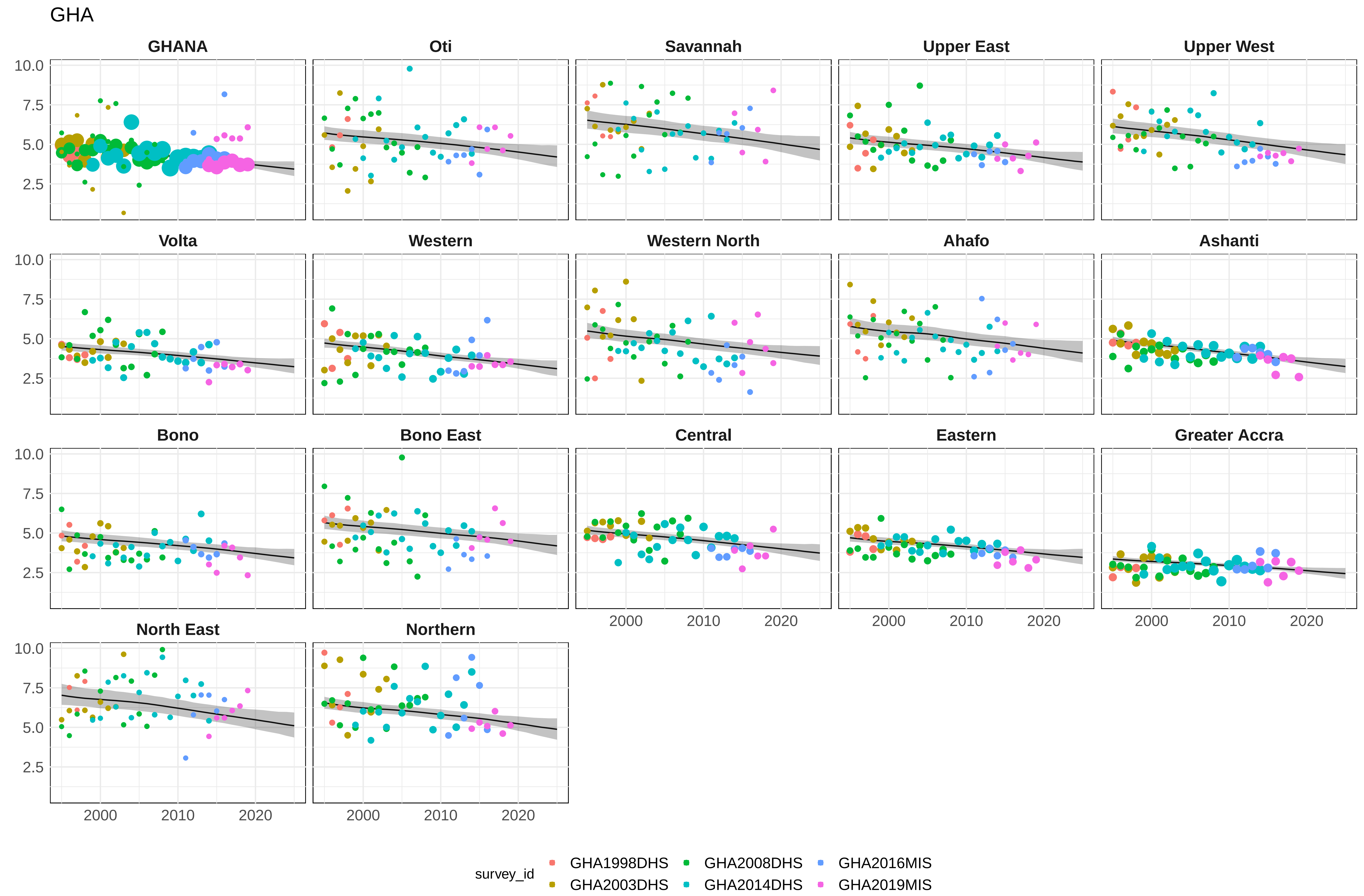
GHA
GHANA
Oti
Savannah
Upper East
Upper West
Volta
Western
Western North
Ahafo
Ashanti
Bono
Bono East
Central
Eastern
Greater Accra
North East
Northern
10.0
7.5
5.0
2.5
2000
2010
2020
survey_id
GHA1998DHS
GHA2003DHS
GHA2008DHS
GHA2014DHS
GHA2016MIS
GHA2019MIS

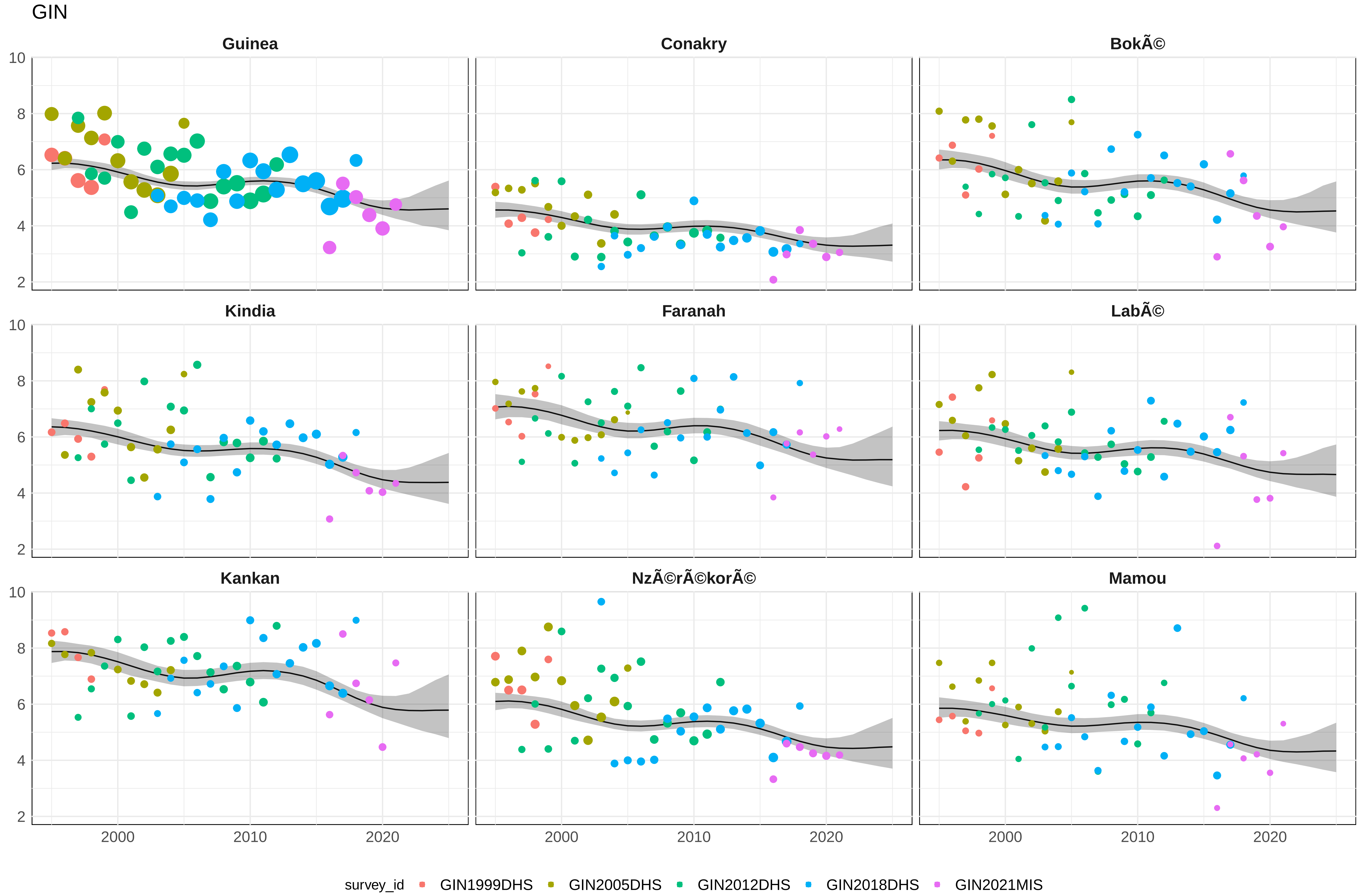

GIN
Guinea
Conakry
BokÃ©
Kindia
Faranah
LabÃ©
Kankan
NzÃ©rÃ©korÃ©
Mamou
10
8
6
4
2
2000
2010
2020
survey_id
GIN1999DHS
GIN2005DHS
GIN2012DHS
GIN2018DHS
GIN2021MIS

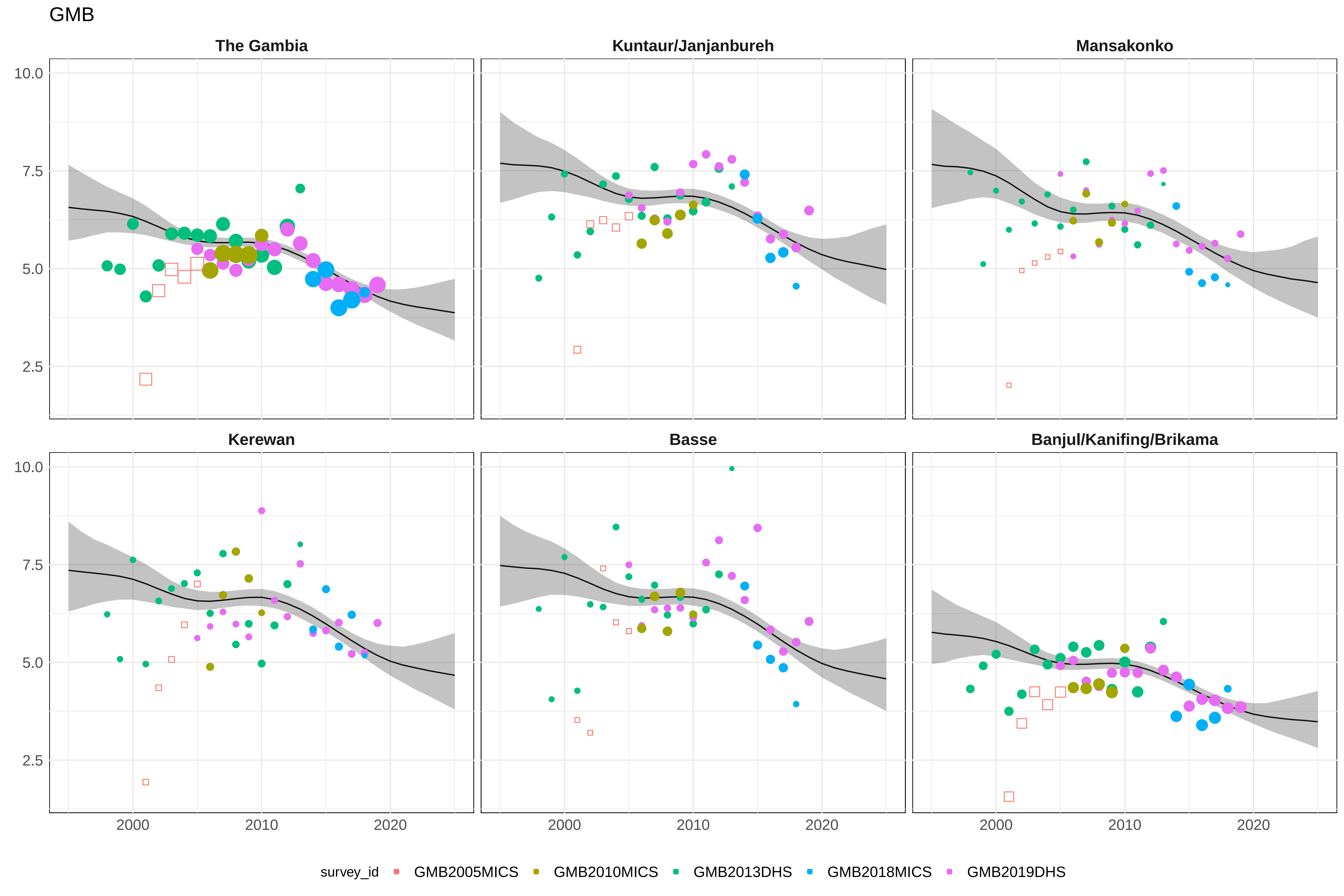
GMB
The Gambia
Kuntaur/Janjanbureh
Mansakonko
Kerewan
Basse
Banjul/Kanifing/Brikama
10.0
7.5
5.0
2.5
2000
2010
2020
survey_id
GMB2005MICS
GMB2010MICS
GMB2013DHS
GMB2018MICS
GMB2019DHS

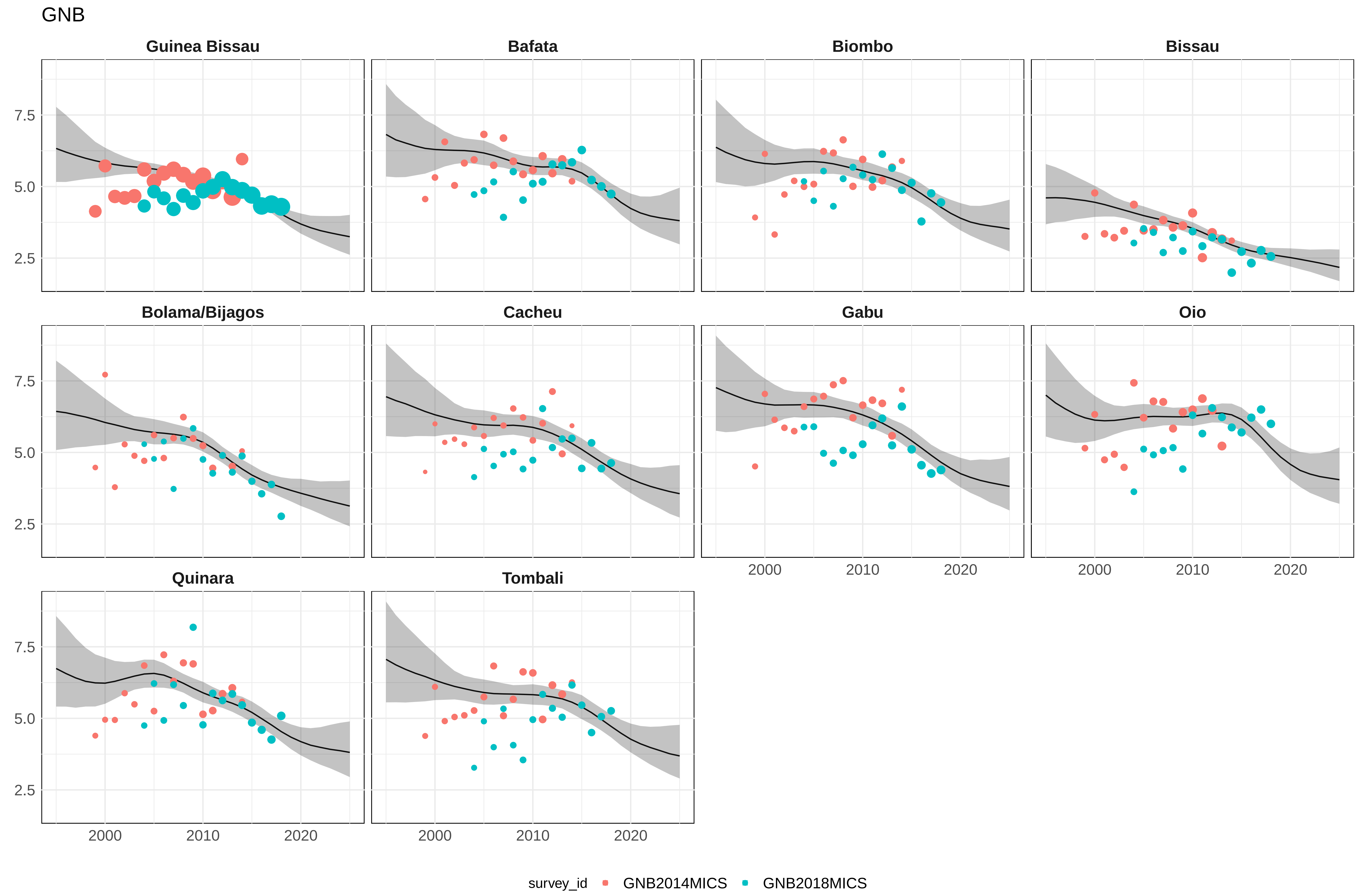

GNB
Guinea Bissau
Bafata
Biombo
Bissau
Bolama/Bijagos
Cacheu
Gabu
Oio
Quinara
Tombali
7.5
5.0
2.5
2000
2010
2020
survey_id
GNB2014MICS
GNB2018MICS

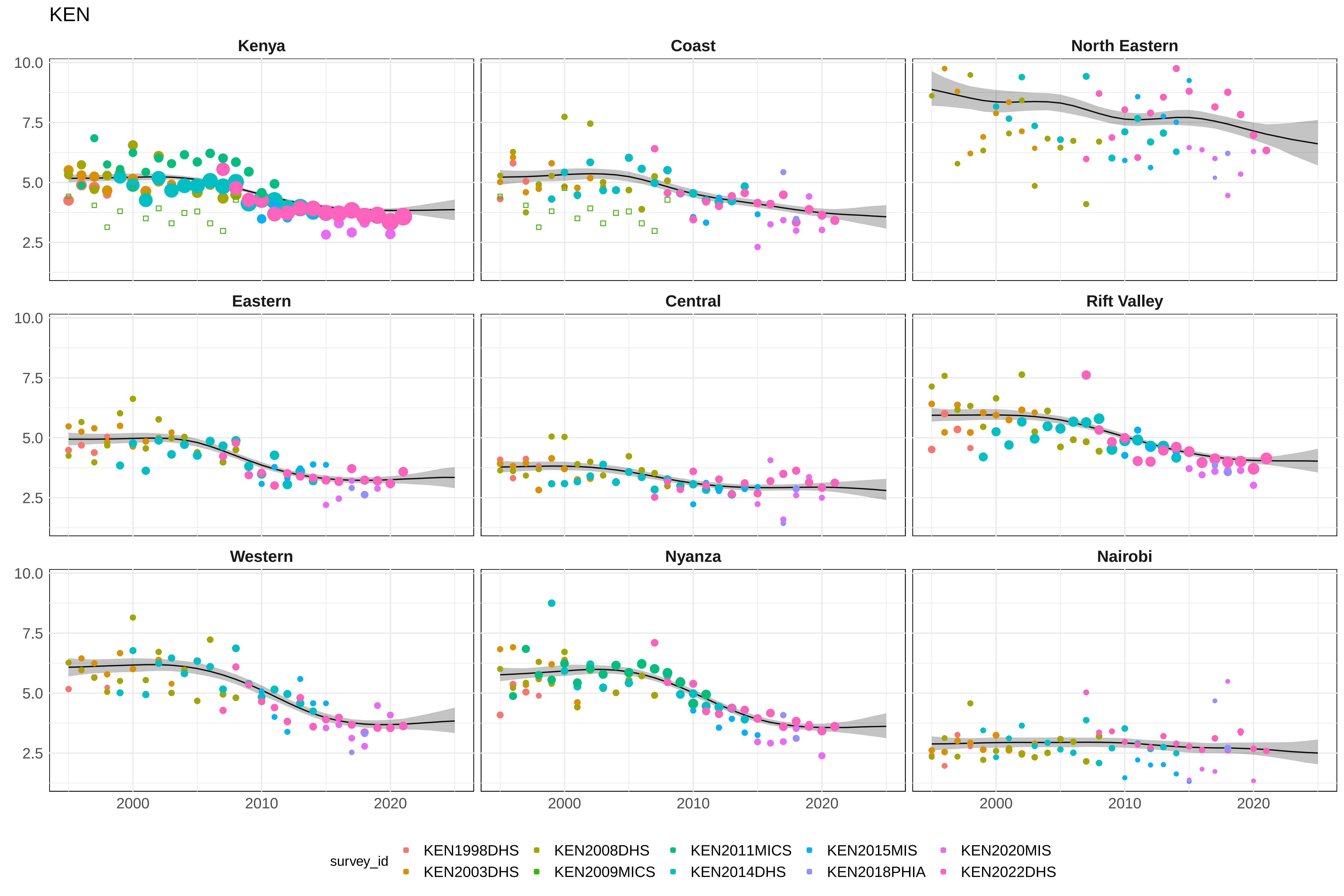
KEN
Kenya
Coast
North Eastern
Eastern
Central
Rift Valley
Western
Nyanza
Nairobi
10.0
7.5
5.0
2.5
2000
2010
2020
survey_id
KEN1998DHS
KEN2003DHS
KEN2008DHS
KEN2009MICS
KEN2011MICS
KEN2014DHS
KEN2015MIS
KEN2018PHIA
KEN2020MIS
KEN2022DHS

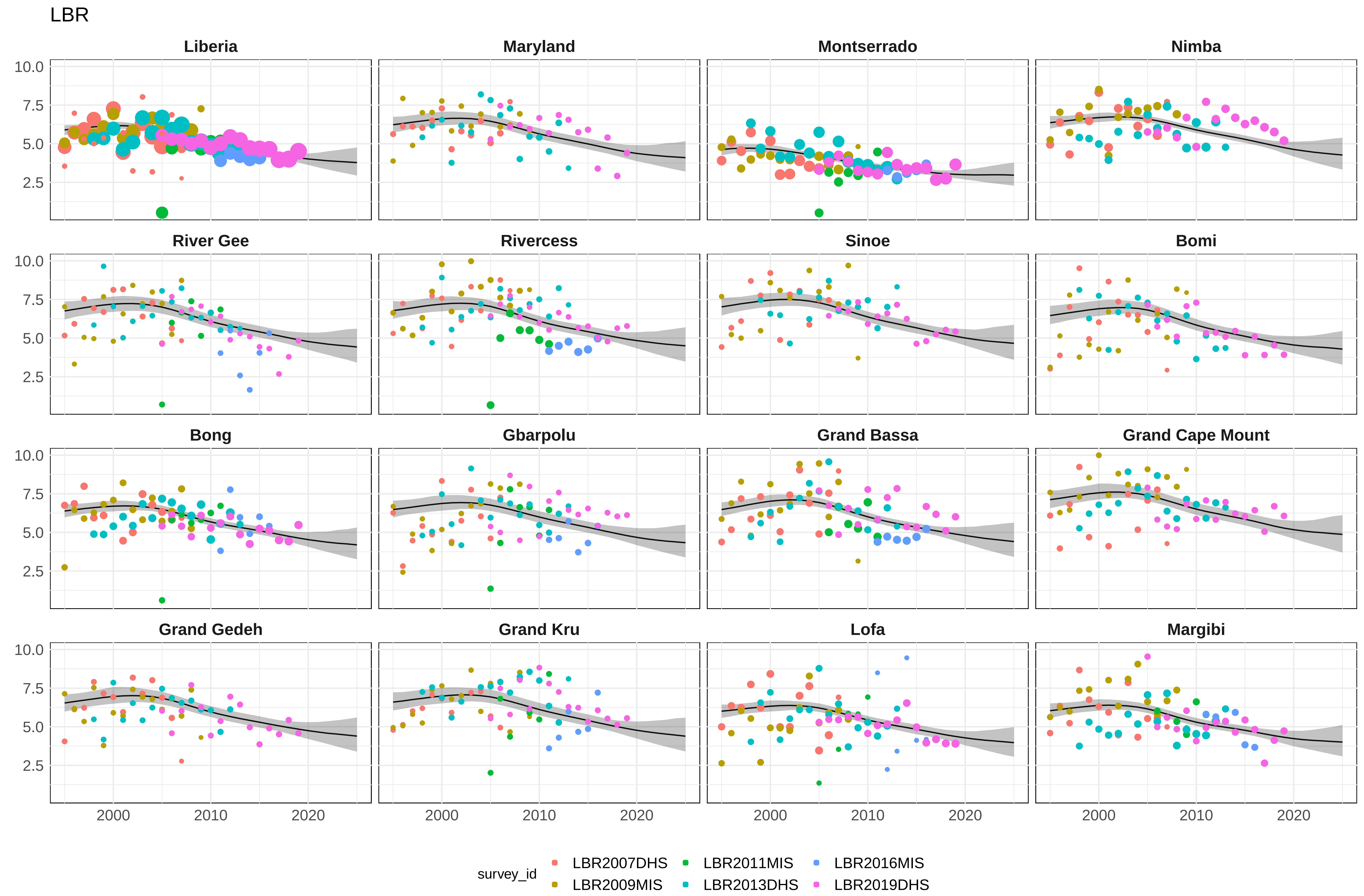

LBR
Liberia
Maryland
Montserrado
Nimba
River Gee
Rivercess
Sinoe
Bomi
Bong
Gbarpolu
Grand Bassa
Grand Cape Mount
Grand Gedeh
Grand Kru
Lofa
Margibi
10.0
7.5
5.0
2.5
2000
2010
2020
survey_id
LBR2007DHS
LBR2009MIS
LBR2011MIS
LBR2013DHS
LBR2016MIS
LBR2019DHS

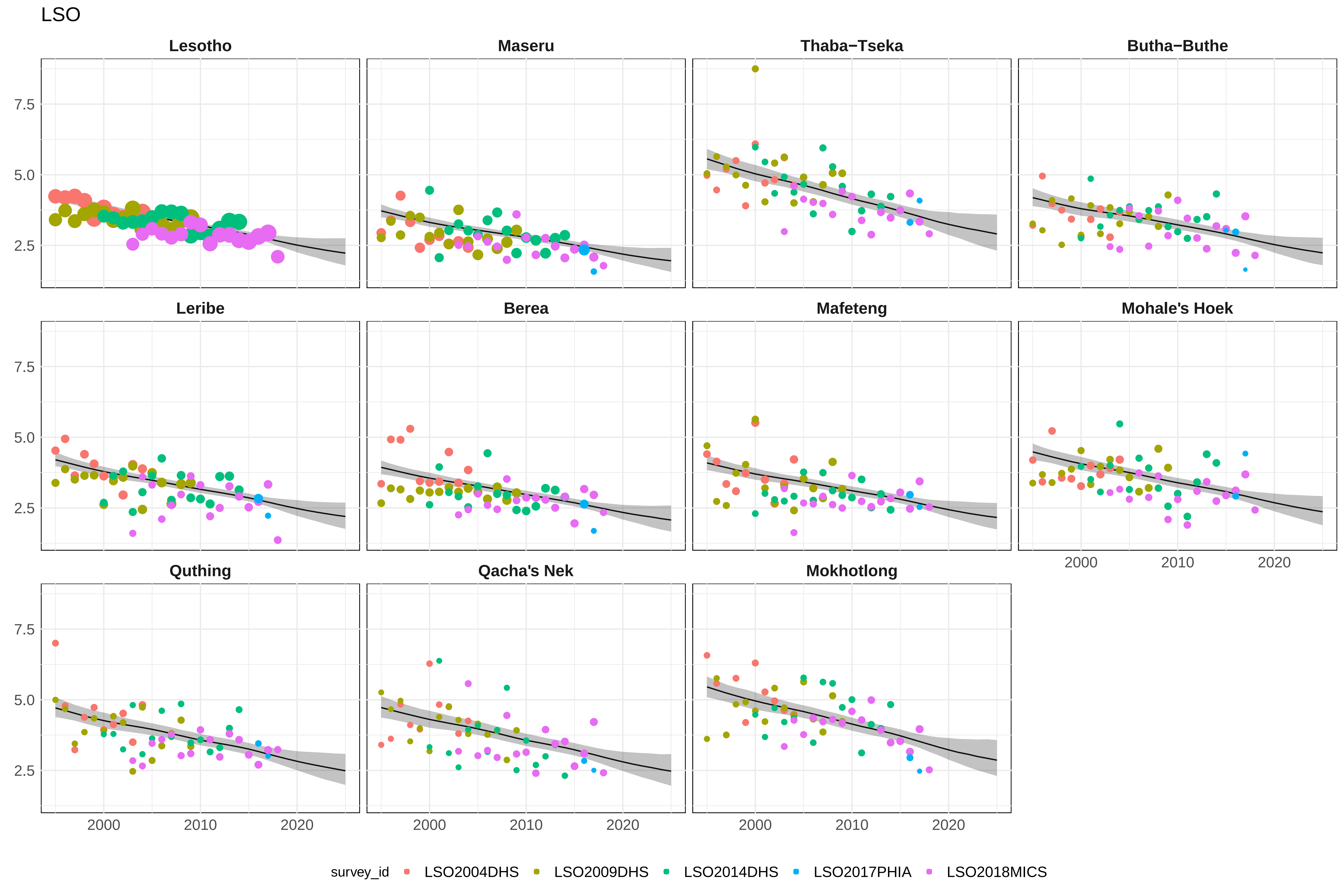

LSO
Lesotho
Maseru
Thaba−Tseka
Butha−Buthe
Leribe
Berea
Mafeteng
Mohale's Hoek
Quthing
Qacha's Nek
Mokhotlong
7.5
5.0
2.5
2000
2010
2020
survey_id
LSO2004DHS
LSO2009DHS
LSO2014DHS
LSO2017PHIA
LSO2018MICS

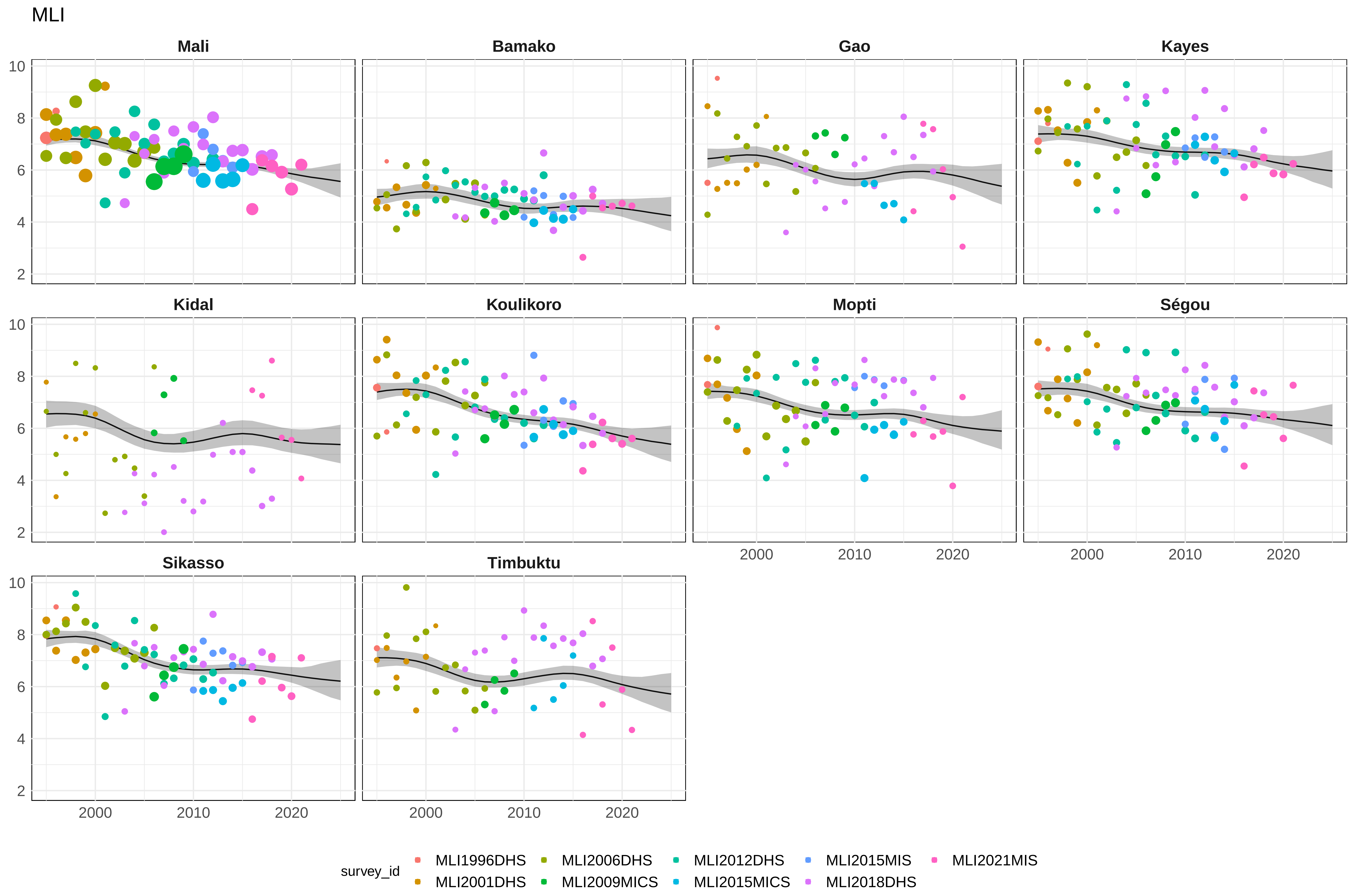

MLI
Mali
Bamako
Gao
Kayes
Kidal
Koulikoro
Mopti
Ségou
Sikasso
Timbuktu
10
8
6
4
2
2000
2010
2020
survey_id
MLI1996DHS
MLI2001DHS
MLI2006DHS
MLI2009MICS
MLI2012DHS
MLI2015MICS
MLI2015MIS
MLI2018DHS
MLI2021MIS

MOZ

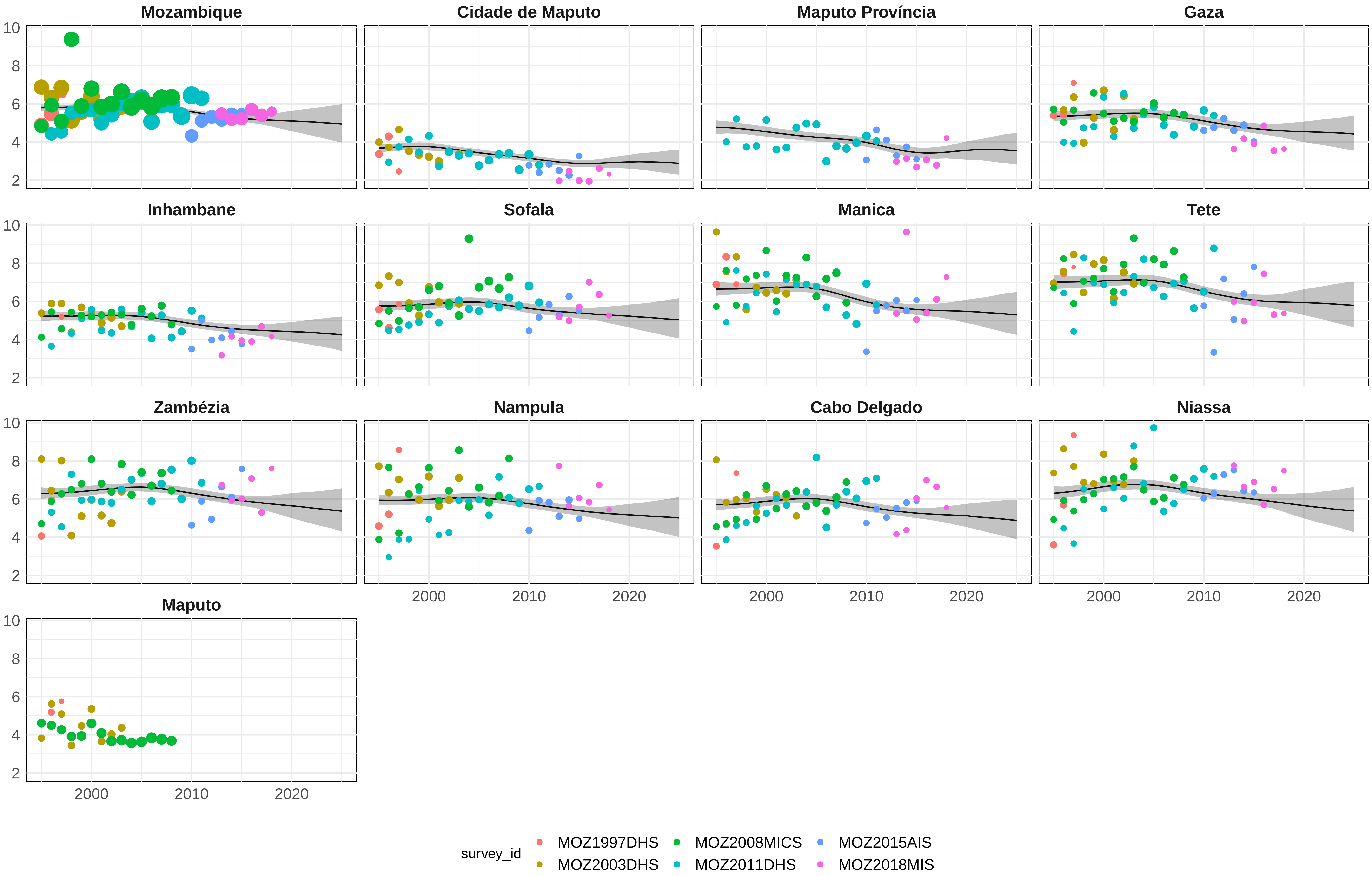

Mozambique
Cidade de Maputo
Maputo Província
Gaza
Inhambane
Sofala
Manica
Tete
Zambézia
Nampula
Cabo Delgado
Niassa
Maputo
10
8
6
4
2
2000
2010
2020
survey_id
MOZ1997DHS
MOZ2003DHS
MOZ2008MICS
MOZ2011DHS
MOZ2015AIS
MOZ2018MIS

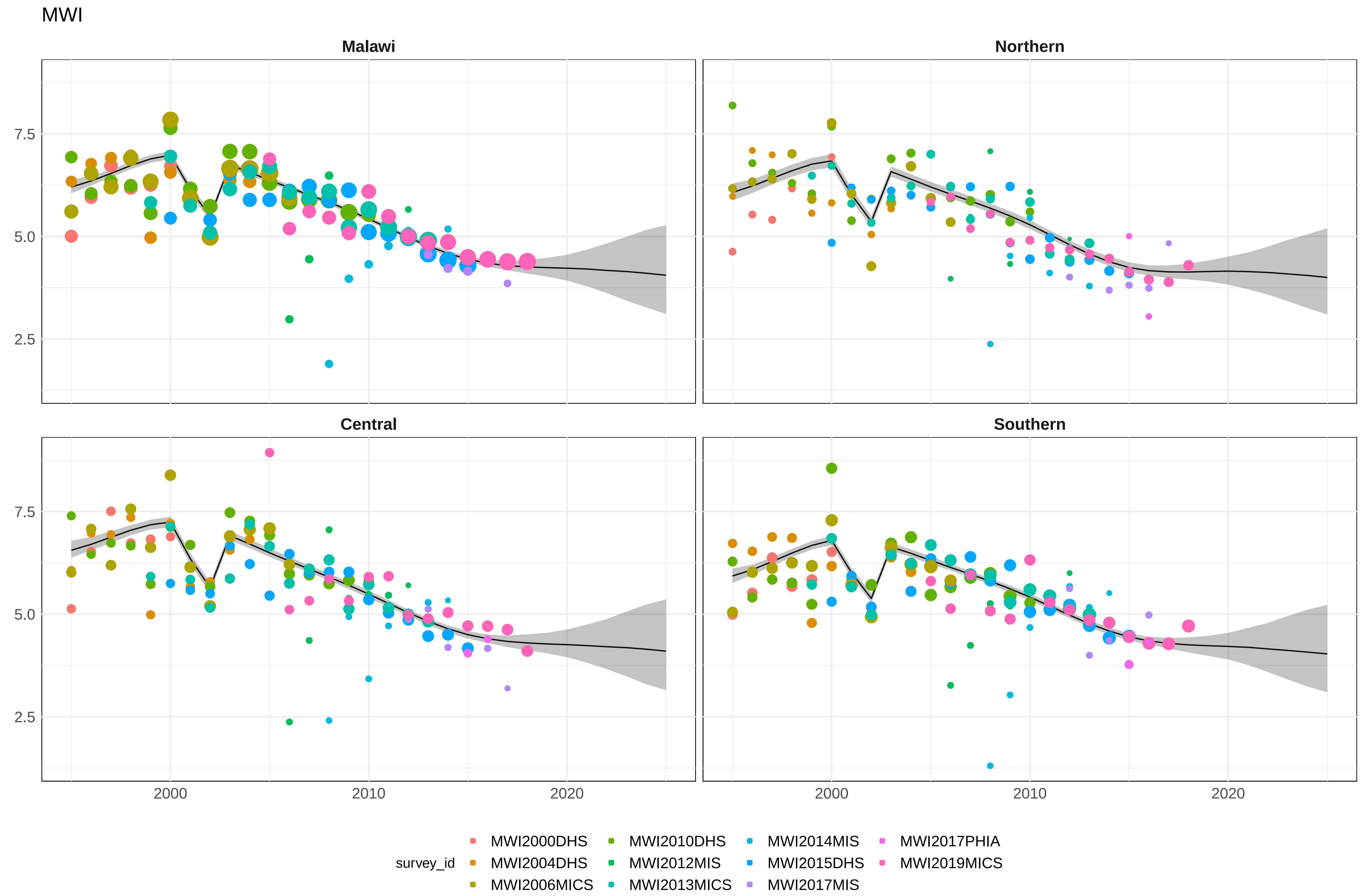

MWI
Malawi
Northern
Central
Southern
7.5
5.0
2.5
2000
2010
2020
survey_id
MWI2000DHS
MWI2004DHS
MWI2006MICS
MWI2010DHS
MWI2012MIS
MWI2013MICS
MWI2014MIS
MWI2015DHS
MWI2017MIS
MWI2017PHIA
MWI2019MICS

NAM

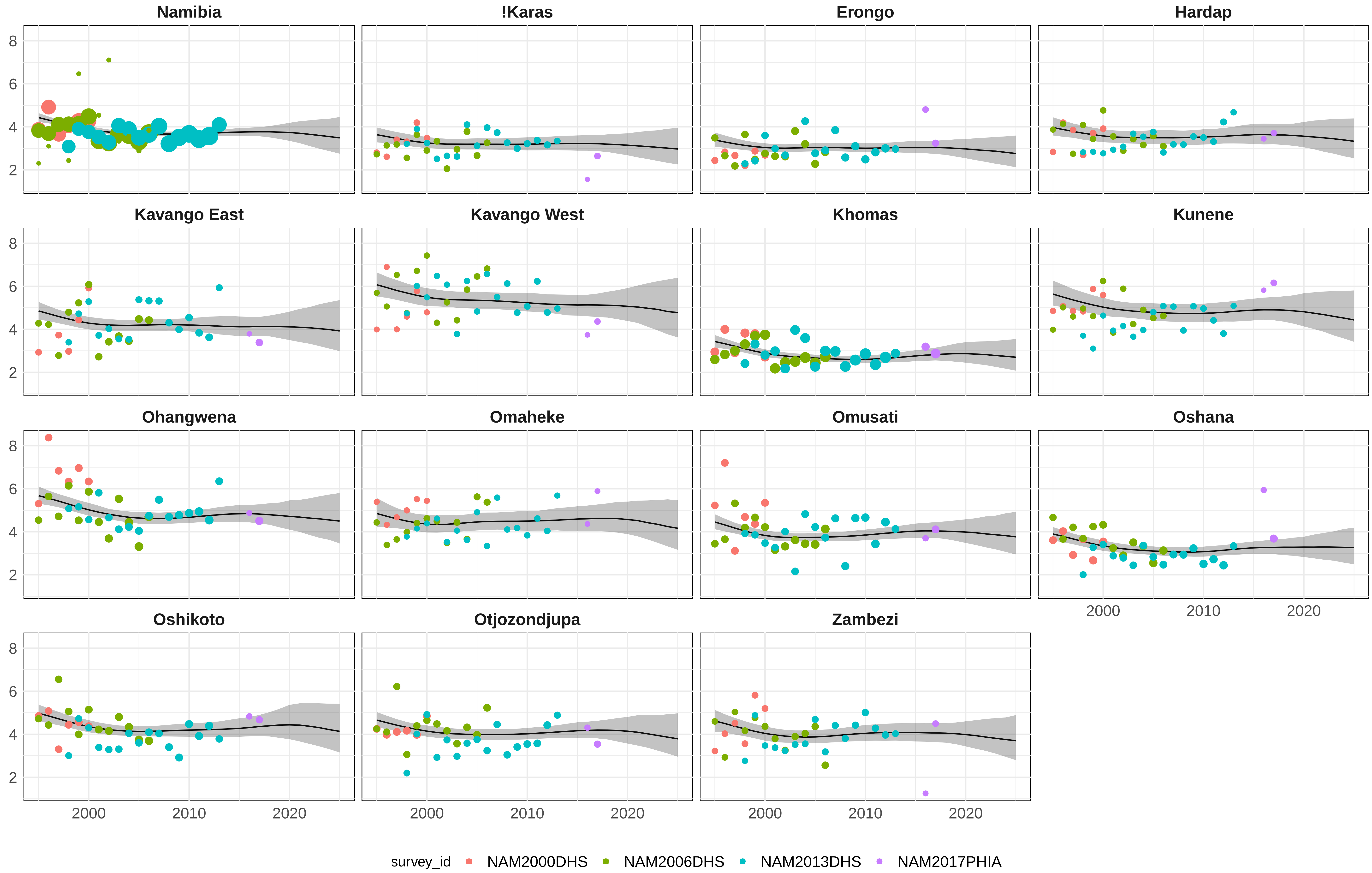

Namibia
!Karas
Erongo
Hardap
Kavango East
Kavango West
Khomas
Kunene
Ohangwena
Omaheke
Omusati
Oshana
Oshikoto
Otjozondjupa
Zambezi
2
4
6
8
2000
2010
2020
survey_id
NAM2000DHS
NAM2006DHS
NAM2013DHS
NAM2017PHIA

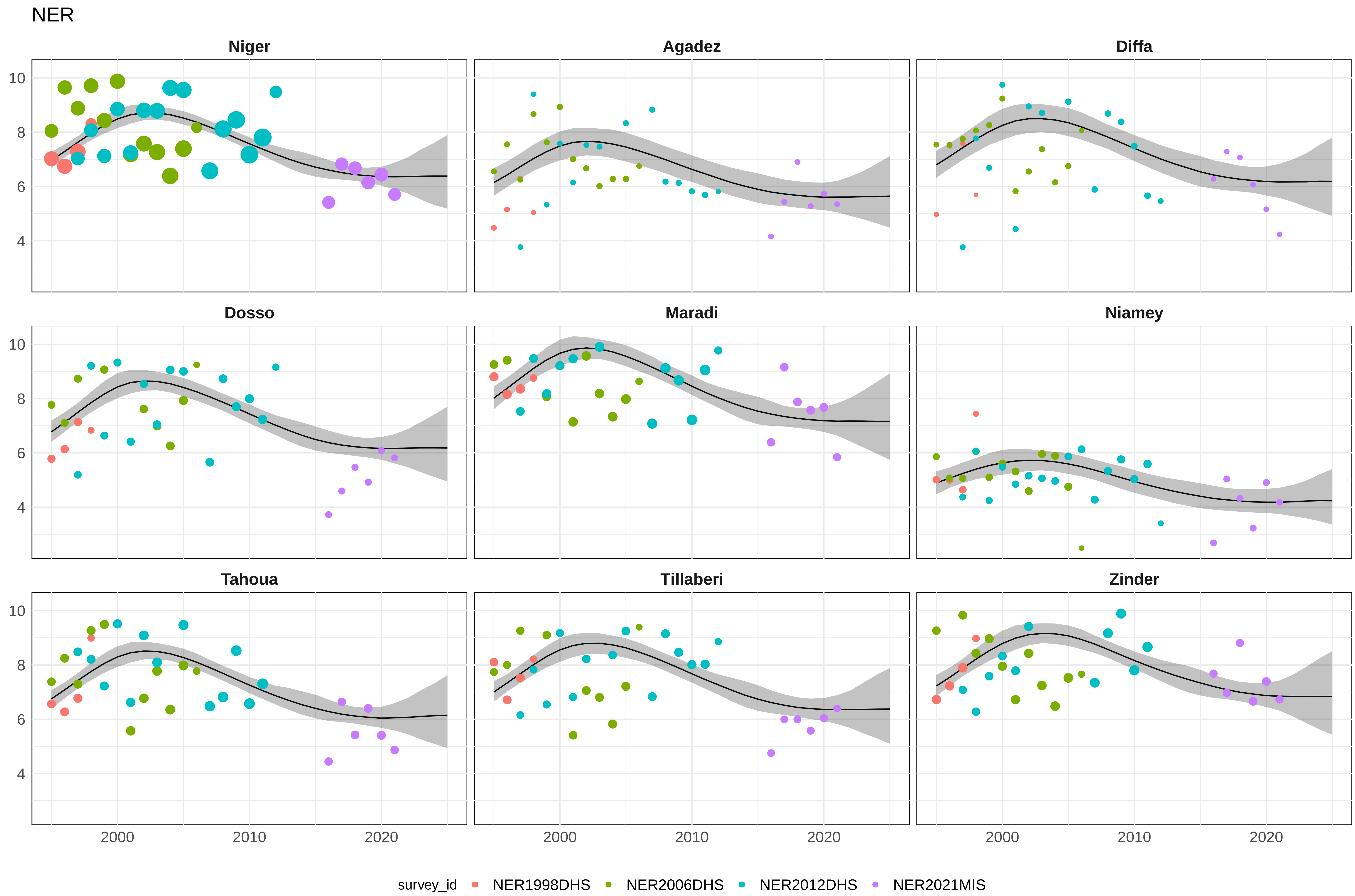

NER
Niger
Agadez
Diffa
Dosso
Maradi
Niamey
Tahoua
Tillaberi
Zinder
10
8
6
4
2000
2010
2020
survey_id
NER1998DHS
NER2006DHS
NER2012DHS
NER2021MIS

NGA

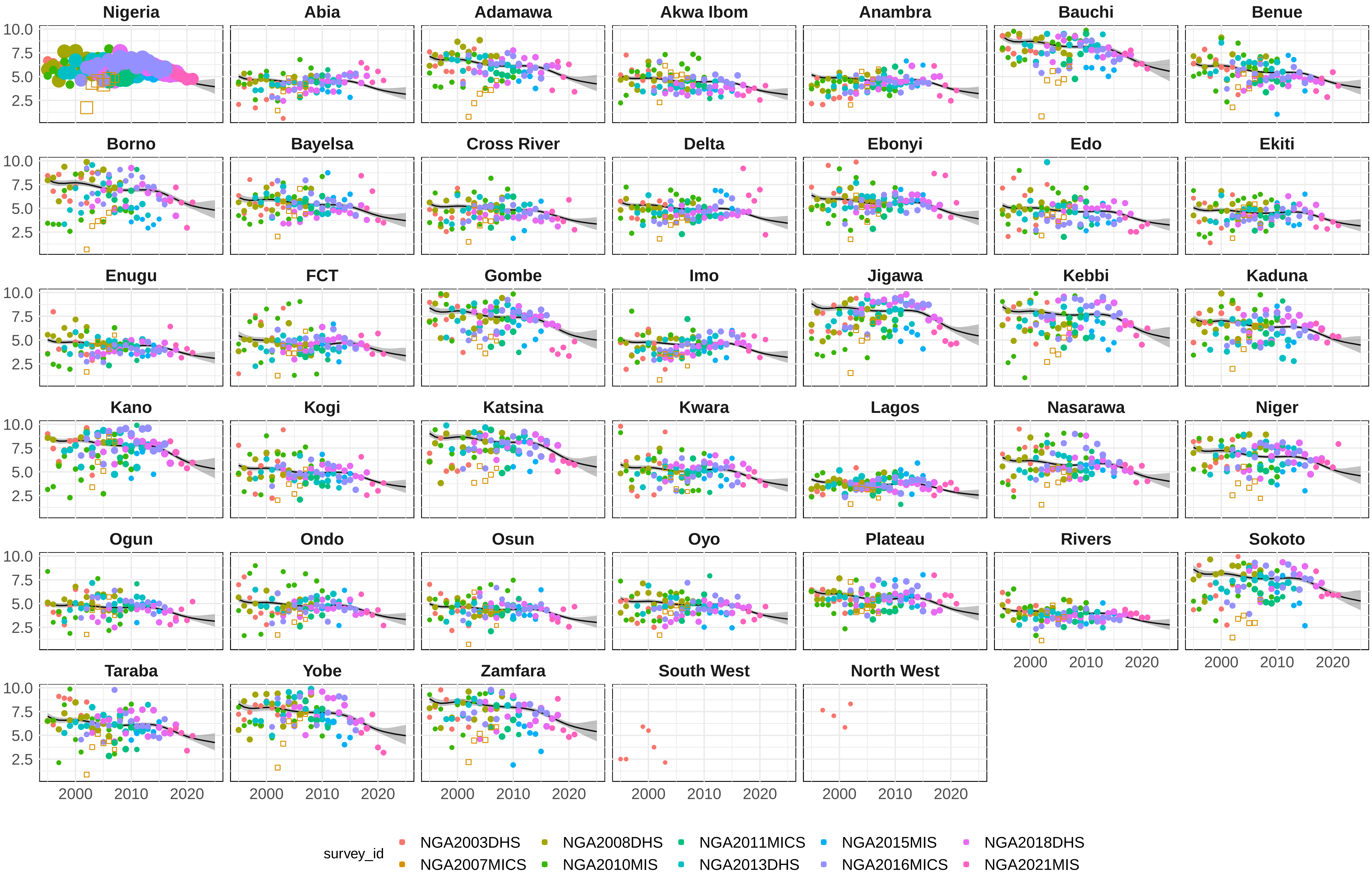
Nigeria
Abia
Adamawa
Akwa Ibom
Anambra
Bauchi
Benue
Borno
Bayelsa
Cross River
Delta
Ebonyi
Edo
Ekiti
Enugu
FCT
Gombe
Imo
Jigawa
Kebbi
Kaduna
Kano
Kogi
Katsina
Kwara
Lagos
Nasarawa
Niger
Ogun
Ondo
Osun
Oyo
Plateau
Rivers
Sokoto
Taraba
Yobe
Zamfara
South West
North West
10.0
7.5
5.0
2.5
2000
2010
2020
survey_id
NGA2003DHS
NGA2007MICS
NGA2008DHS
NGA2010MIS
NGA2011MICS
NGA2013DHS
NGA2015MIS
NGA2016MICS
NGA2018DHS
NGA2021MIS

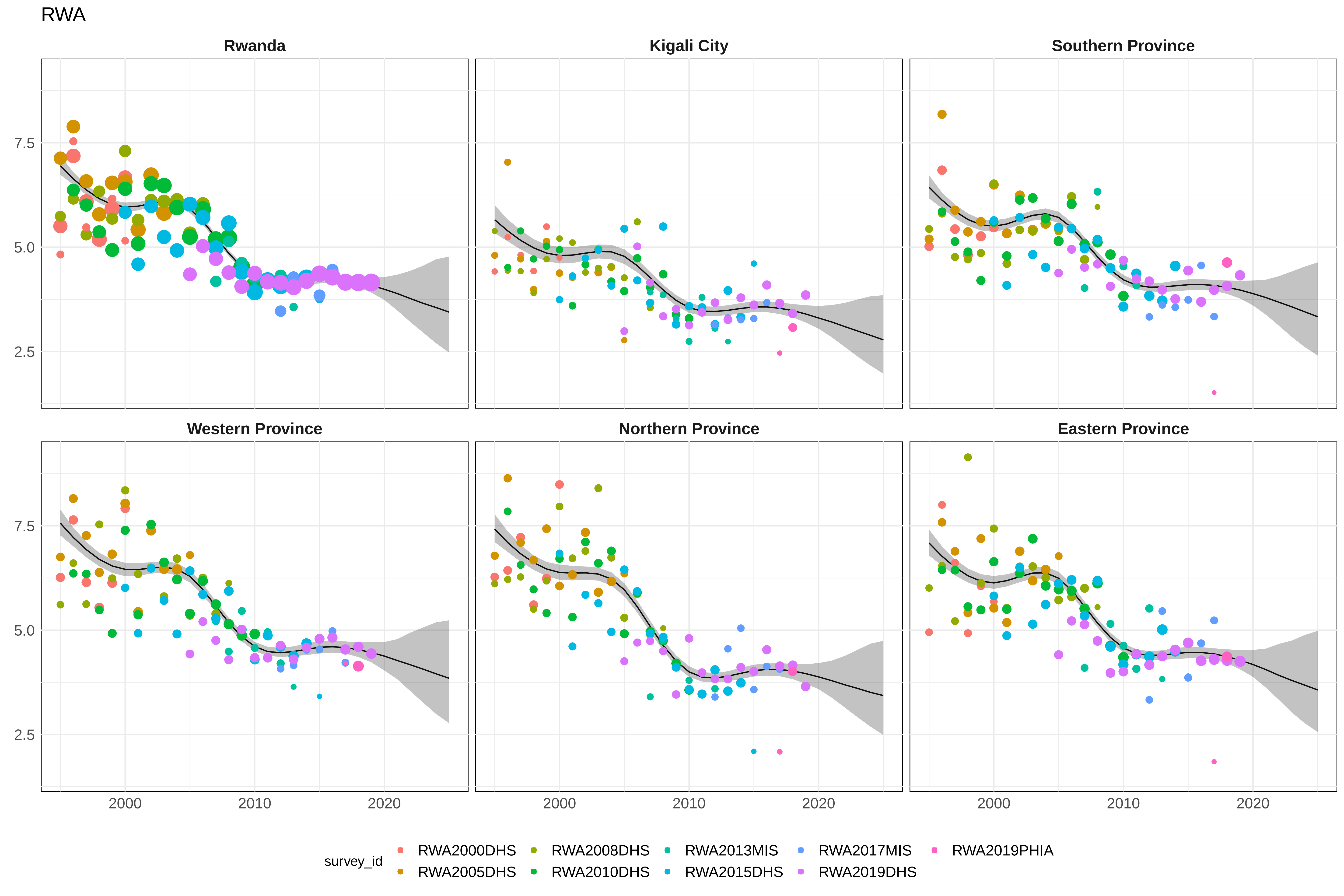
RWA
Rwanda
Kigali City
Southern Province
Western Province
Northern Province
Eastern Province
7.5
5.0
2.5
2000
2010
2020
survey_id
RWA2000DHS
RWA2005DHS
RWA2008DHS
RWA2010DHS
RWA2013MIS
RWA2015DHS
RWA2017MIS
RWA2019DHS
RWA2019PHIA

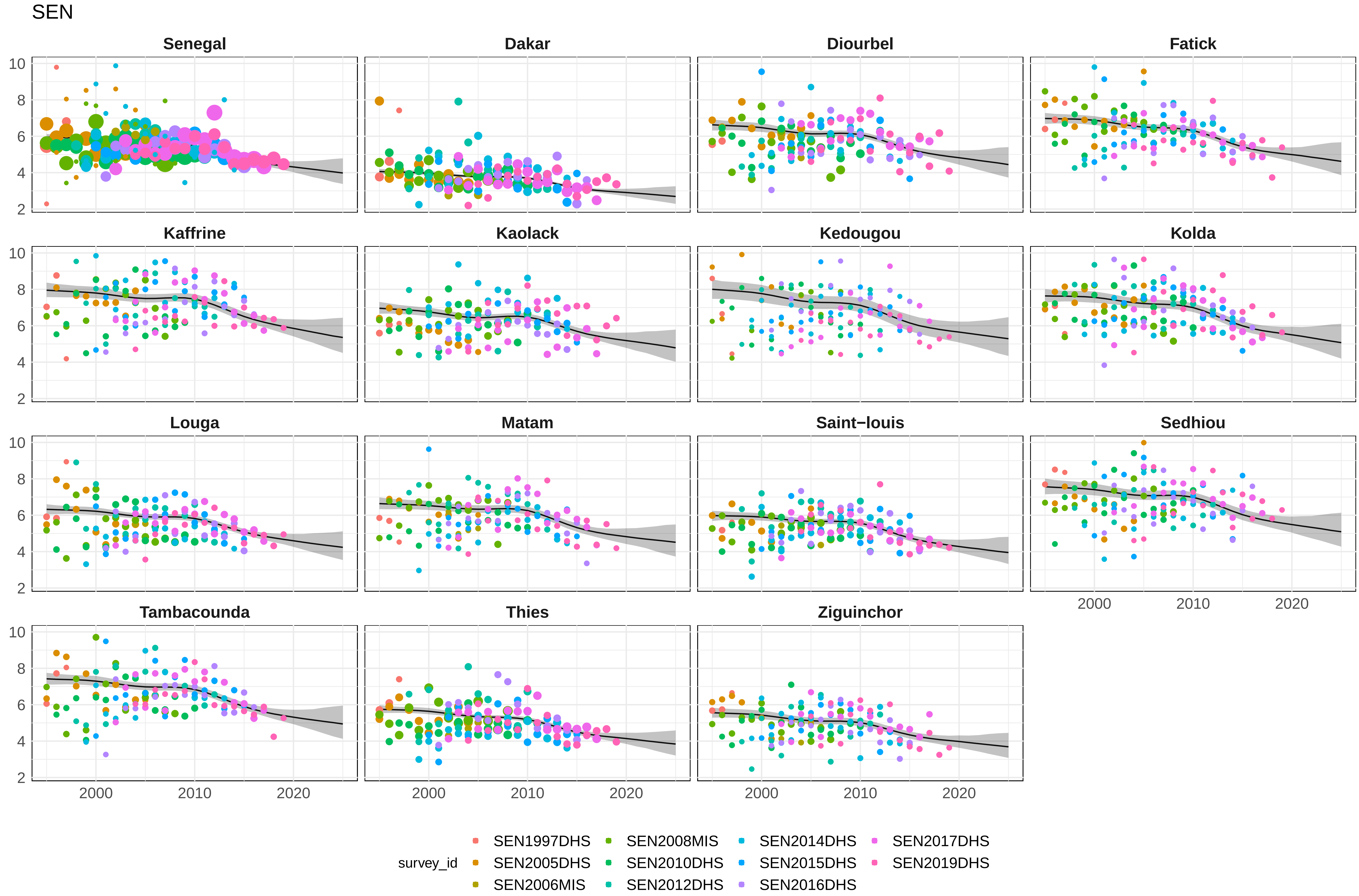
SEN
Senegal
Dakar
Diourbel
Fatick
Kaffrine
Kaolack
Kedougou
Kolda
Louga
Matam
Saint−louis
Sedhiou
Tambacounda
Thies
Ziguinchor
10
8
6
4
2
2000
2010
2020
survey_id
SEN1997DHS
SEN2005DHS
SEN2006MIS
SEN2008MIS
SEN2010DHS
SEN2012DHS
SEN2014DHS
SEN2015DHS
SEN2016DHS
SEN2017DHS
SEN2019DHS

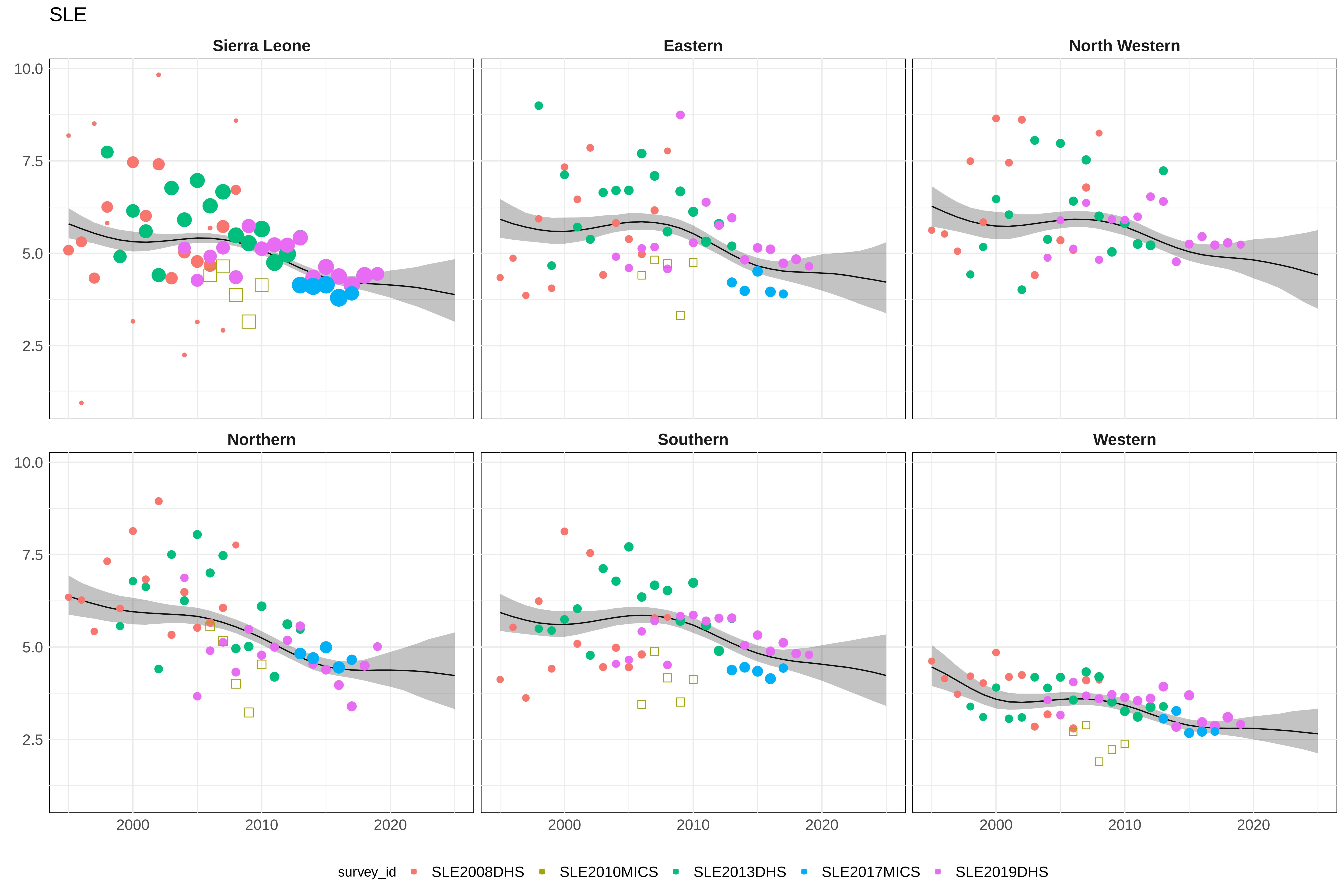
SLE
Sierra Leone
Eastern
North Western
Northern
Southern
Western
10.0
7.5
5.0
2.5
2000
2010
2020
survey_id
SLE2008DHS
SLE2010MICS
SLE2013DHS
SLE2017MICS
SLE2019DHS

SWZ

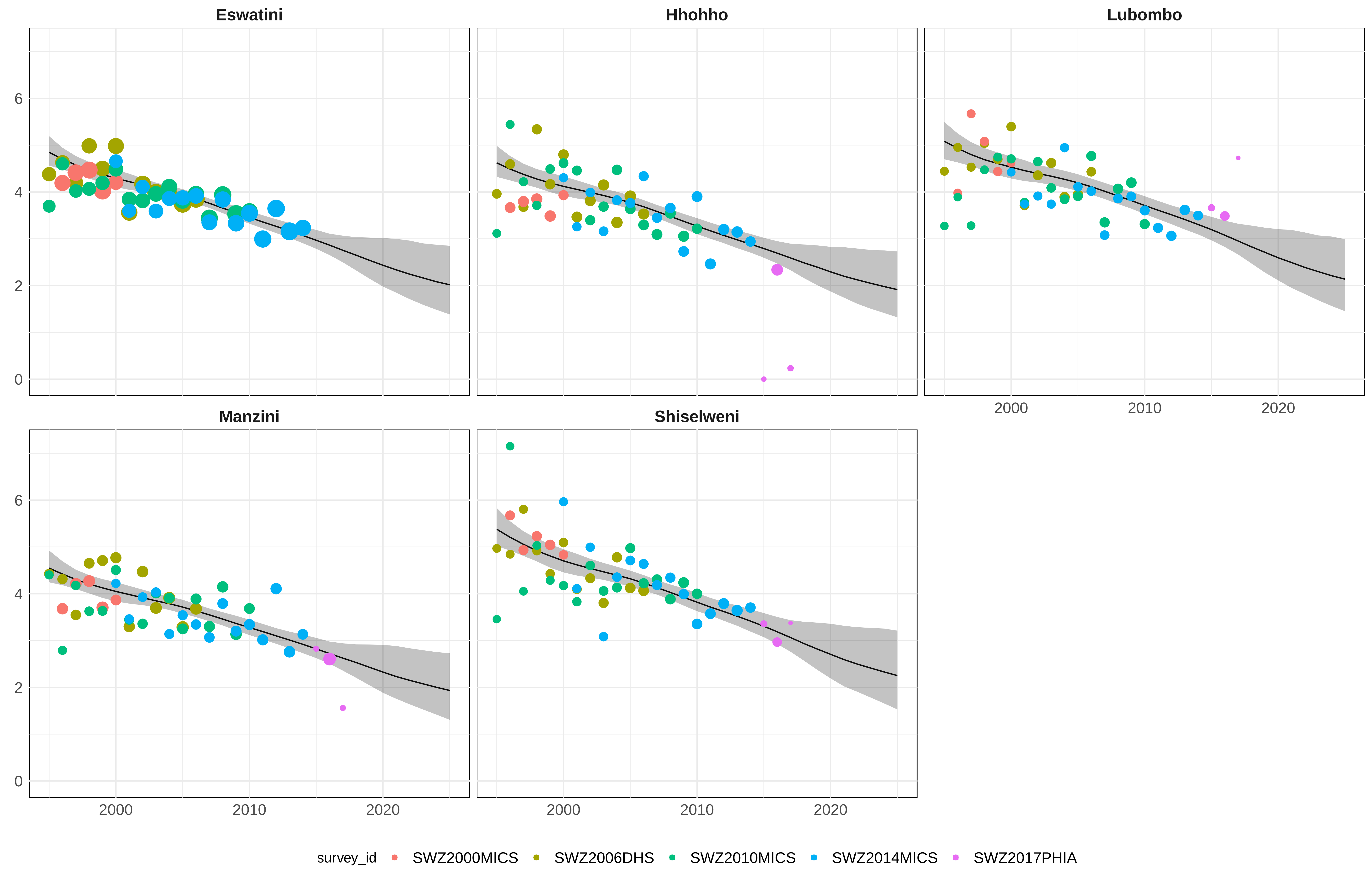

Eswatini
Hhohho
Lubombo
Manzini
Shiselweni
0
2
4
6
2000
2010
2020
survey_id
SWZ2000MICS
SWZ2006DHS
SWZ2010MICS
SWZ2014MICS
SWZ2017PHIA

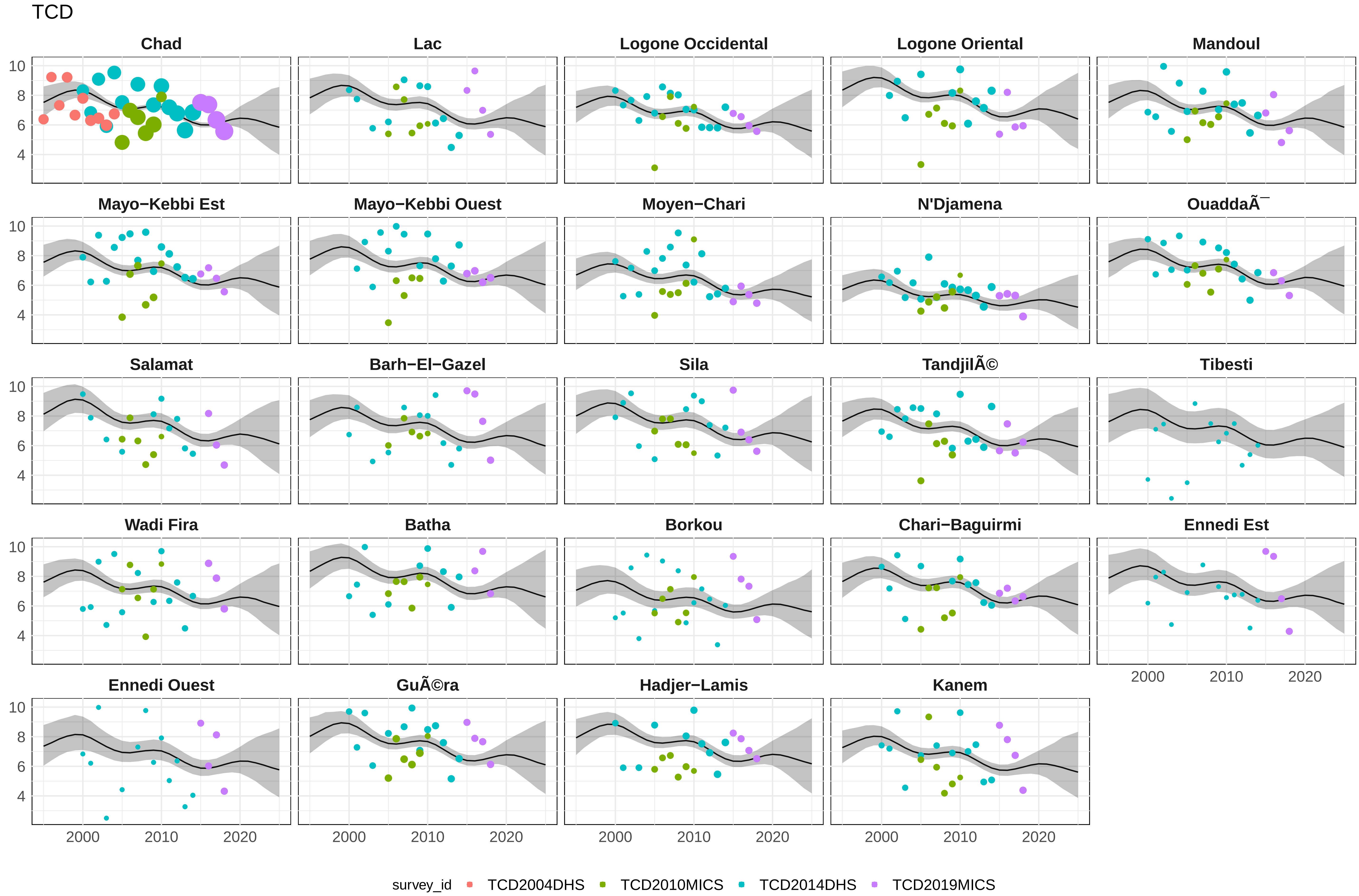

TCD
Chad
Lac
Logone Occidental
Logone Oriental
Mandoul
Mayo−Kebbi Est
Mayo−Kebbi Ouest
Moyen−Chari
N'Djamena
OuaddaÃ¯
Salamat
Barh−El−Gazel
Sila
TandjilÃ©
Tibesti
Wadi Fira
Batha
Borkou
Chari−Baguirmi
Ennedi Est
Ennedi Ouest
GuÃ©ra
Hadjer−Lamis
Kanem
10
8
6
4
2000
2010
2020
survey_id
TCD2004DHS
TCD2010MICS
TCD2014DHS
TCD2019MICS

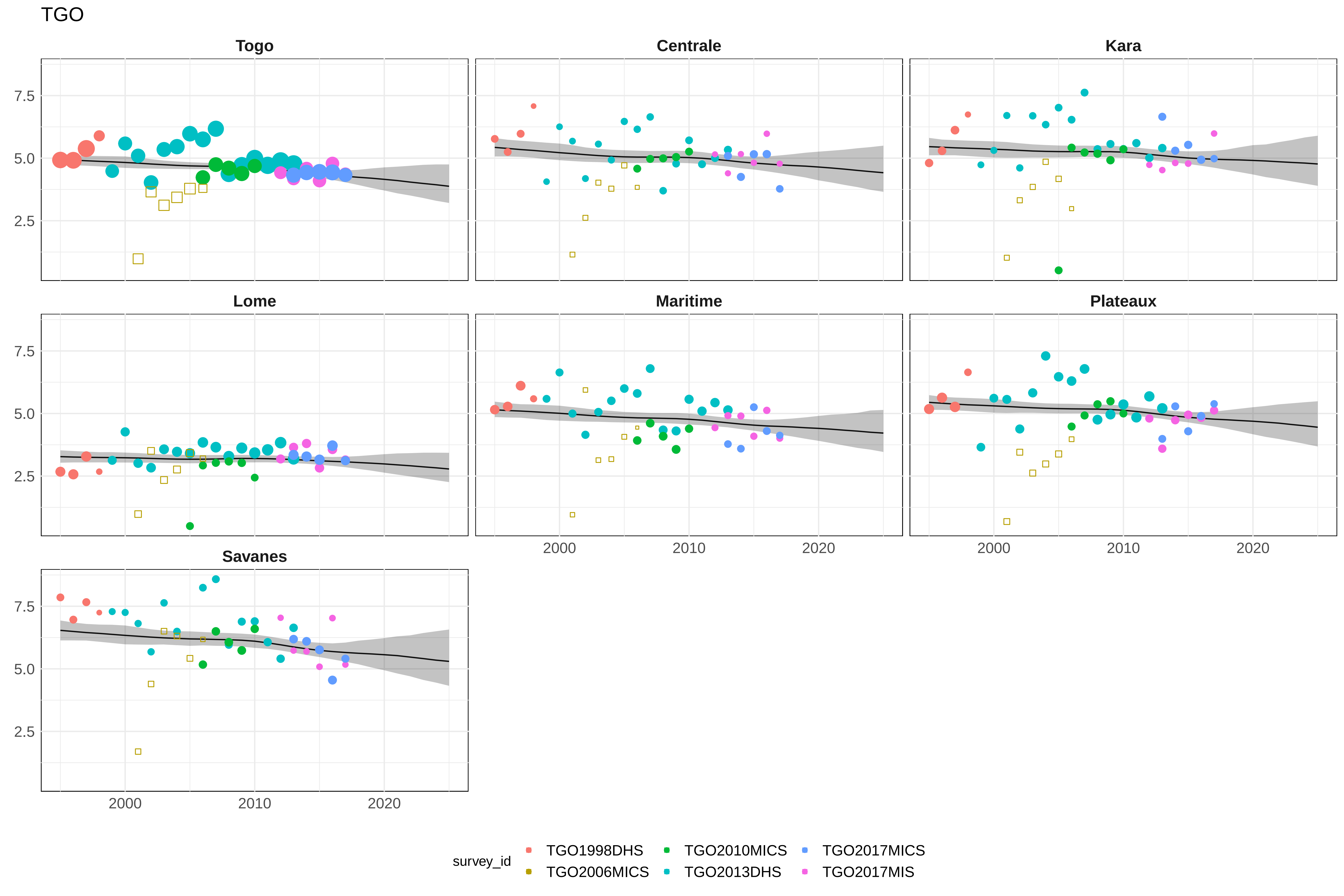

TGO
Togo
Centrale
Kara
Lome
Maritime
Plateaux
Savanes
7.5
5.0
2.5
2000
2010
2020
survey_id
TGO1998DHS
TGO2006MICS
TGO2010MICS
TGO2013DHS
TGO2017MICS
TGO2017MIS

TZA

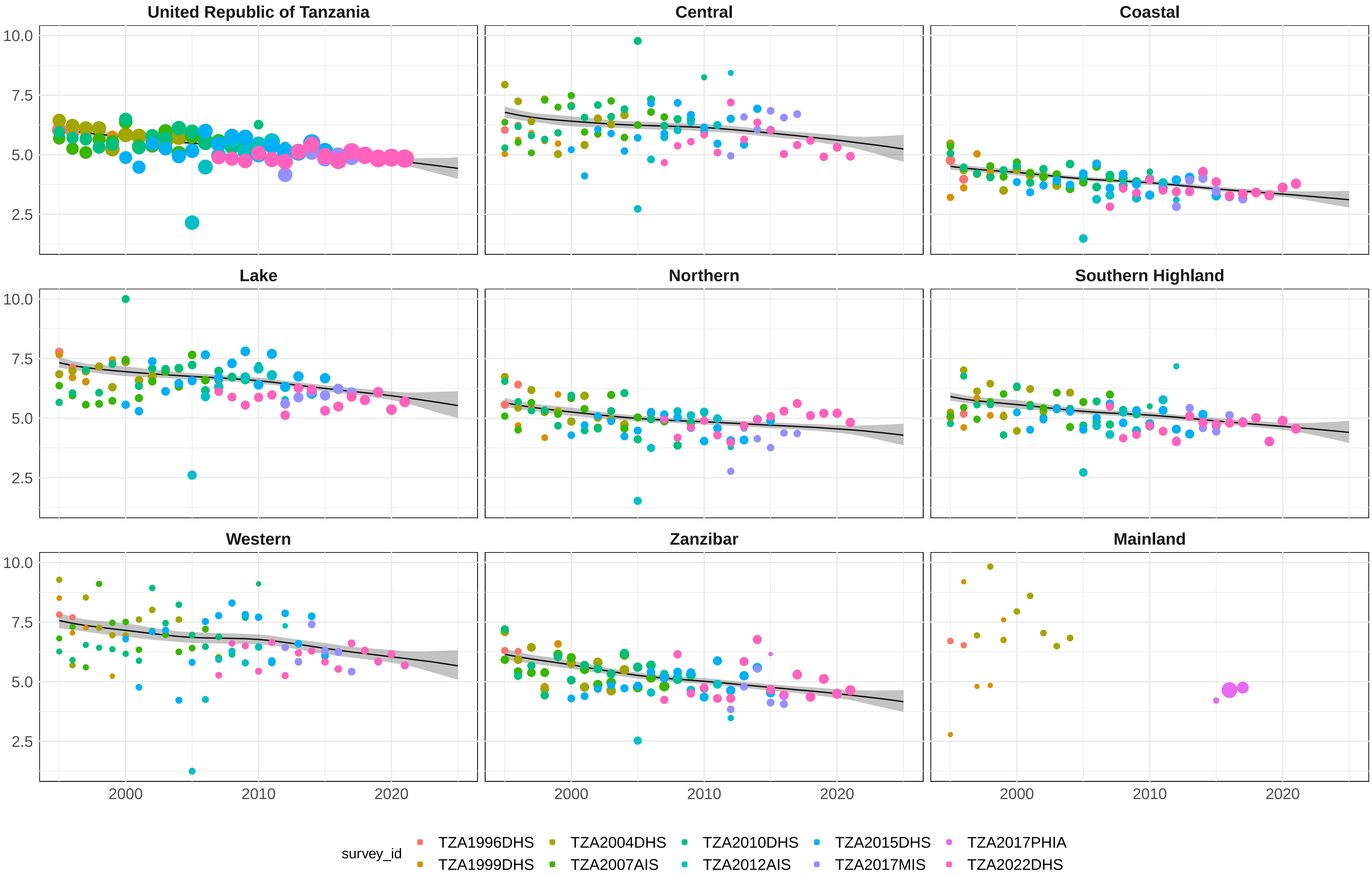

United Republic of Tanzania
Central
Coastal
Lake
Northern
Southern Highland
Western
Zanzibar
Mainland
10.0
7.5
5.0
2.5
2000
2010
2020
survey_id
TZA1996DHS
TZA1999DHS
TZA2004DHS
TZA2007AIS
TZA2010DHS
TZA2012AIS
TZA2015DHS
TZA2017MIS
TZA2017PHIA
TZA2022DHS

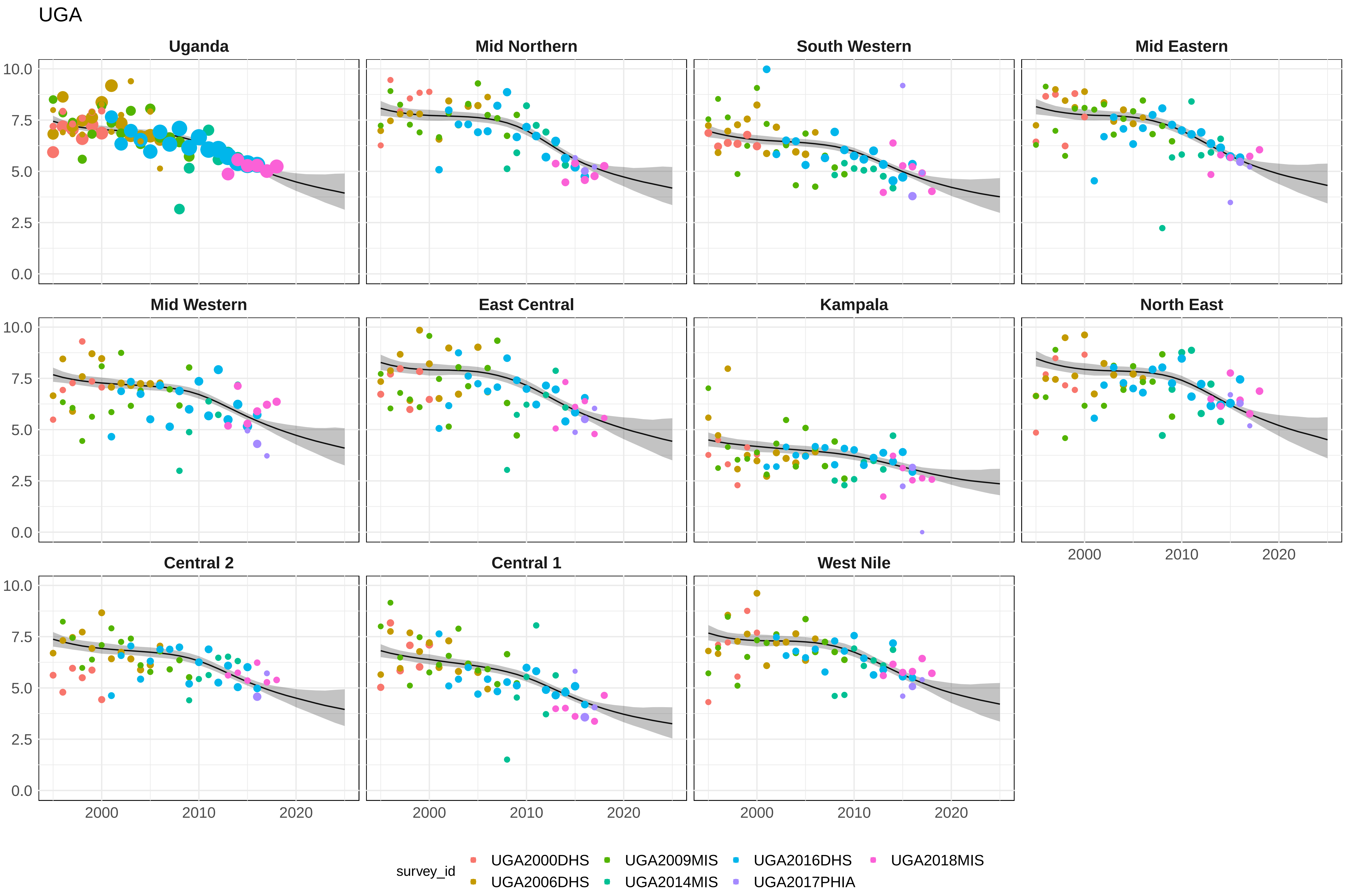

UGA
Uganda
Mid Northern
South Western
Mid Eastern
Mid Western
East Central
Kampala
North East
Central 2
Central 1
West Nile
0.0
2.5
5.0
7.5
10.0
2000
2010
2020
survey_id
UGA2000DHS
UGA2006DHS
UGA2009MIS
UGA2014MIS
UGA2016DHS
UGA2017PHIA
UGA2018MIS

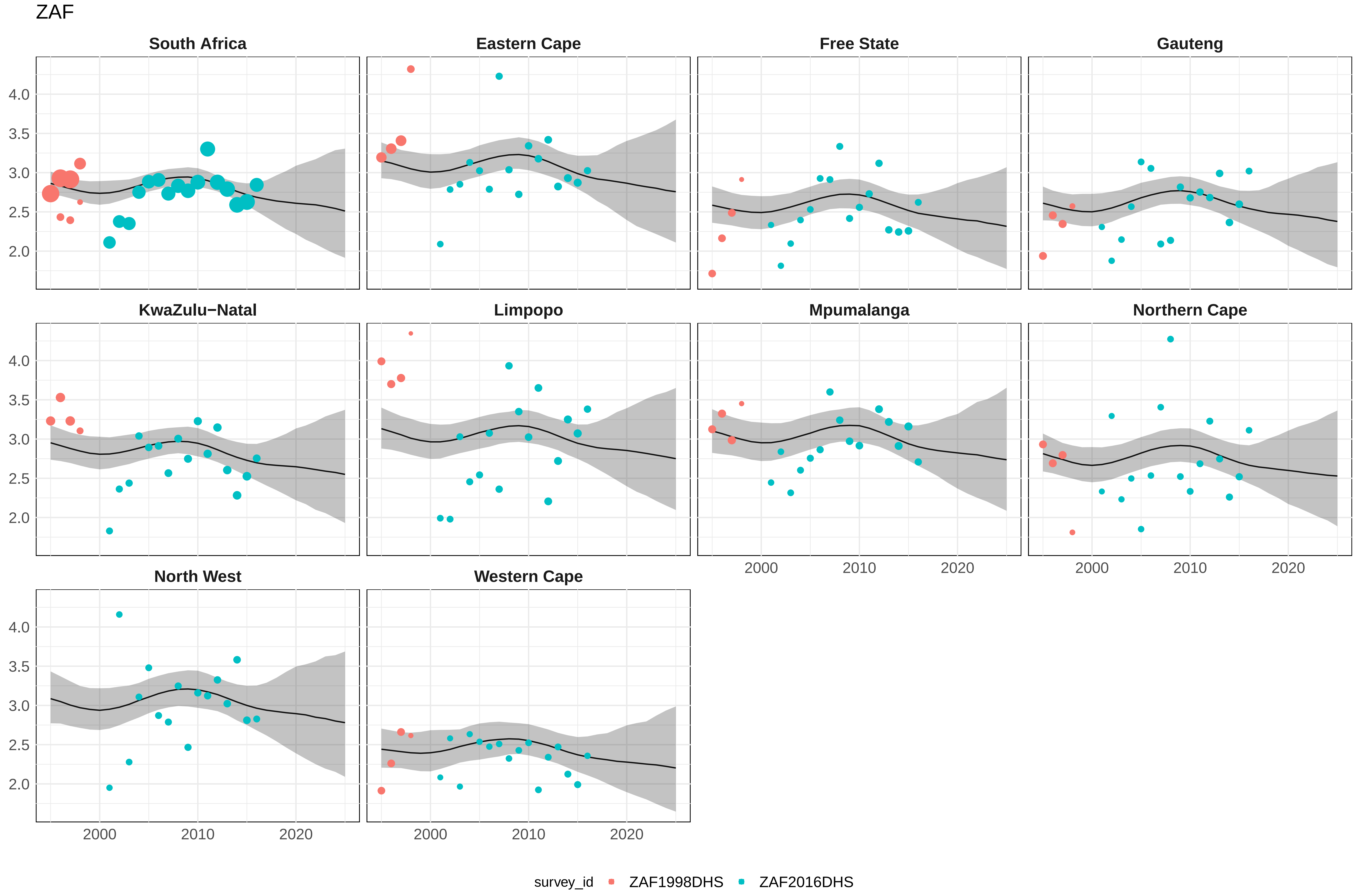

ZAF
South Africa
Eastern Cape
Free State
Gauteng
KwaZulu−Natal
Limpopo
Mpumalanga
Northern Cape
North West
Western Cape
4.0
3.5
3.0
2.5
2.0
2000
2010
2020
survey_id
ZAF1998DHS
ZAF2016DHS

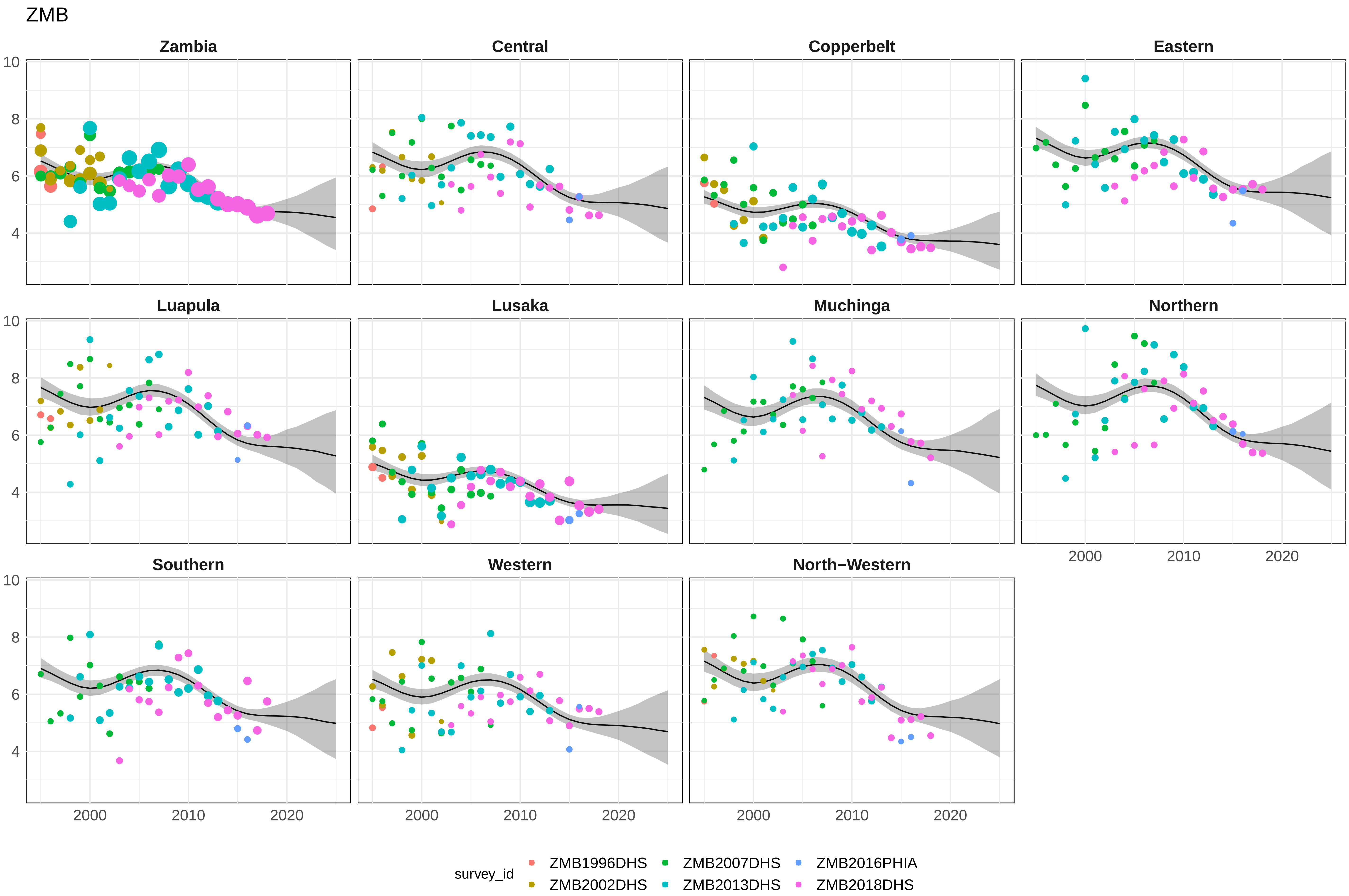
ZMB
Zambia
Central
Copperbelt
Eastern
Luapula
Lusaka
Muchinga
Northern
Southern
Western
North−Western
10
8
6
4
2000
2010
2020
survey_id
ZMB1996DHS
ZMB2002DHS
ZMB2007DHS
ZMB2013DHS
ZMB2016PHIA
ZMB2018DHS

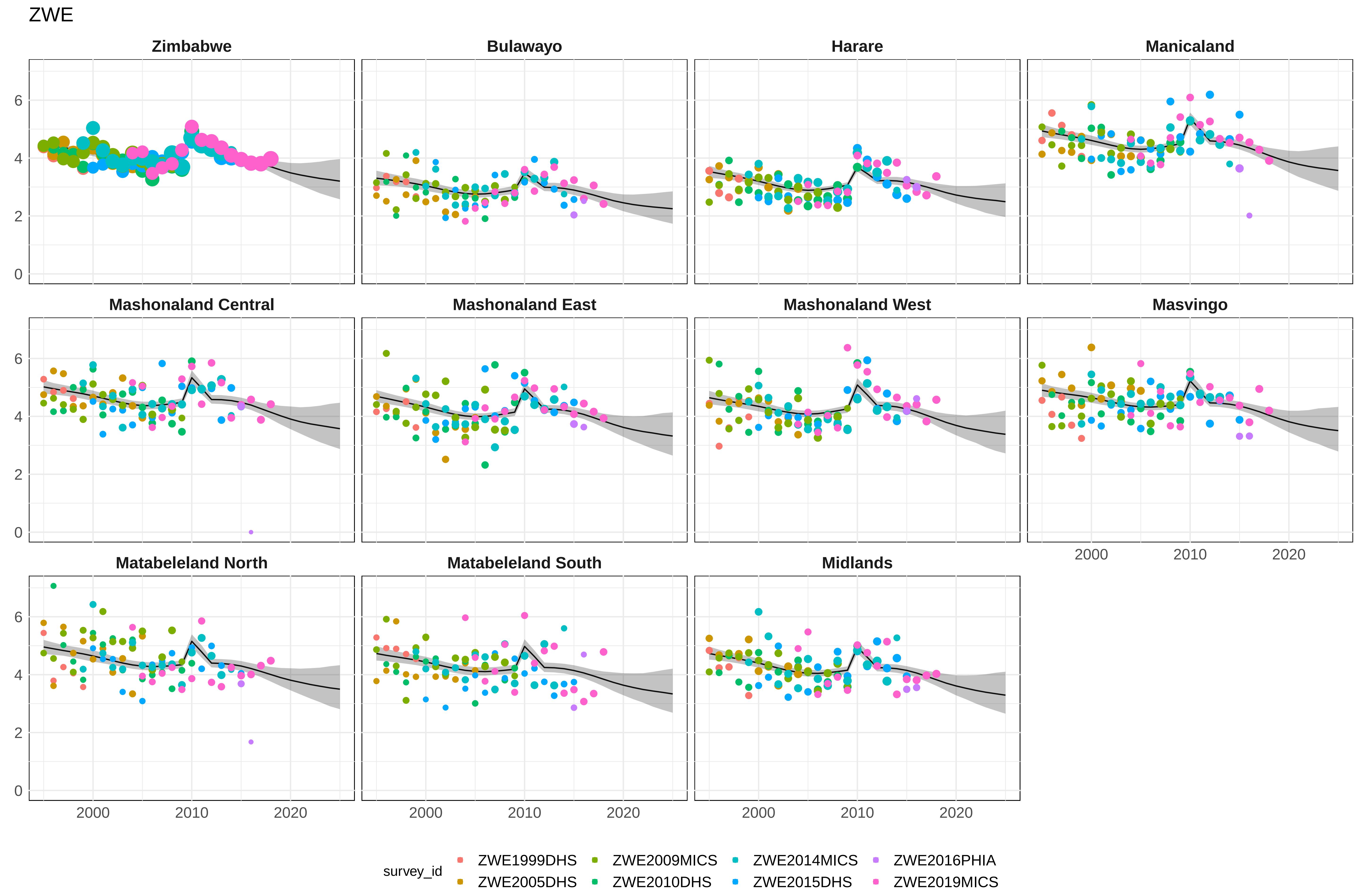

ZWE
Zimbabwe
Bulawayo
Harare
Manicaland
Mashonaland Central
Mashonaland East
Mashonaland West
Masvingo
Matabeleland North
Matabeleland South
Midlands
0
2
4
6
2000
2010
2020
survey_id
ZWE1999DHS
ZWE2005DHS
ZWE2009MICS
ZWE2010DHS
ZWE2014MICS
ZWE2015DHS
ZWE2016PHIA
ZWE2019MICS
